# Characteristics of women diagnosed with endometriosis in England: 2011-2021

**DOI:** 10.1101/2024.12.11.24318835

**Authors:** Hannah Bunk, Emma Campbell, Gemma C Sharp, Vahé Nafilyan, Daniel Ayoubkhani, Isobel L Thompson-Ward

## Abstract

**Objectives:** Endometriosis is a chronic disease and the second most common gynaecological condition in the UK. It is characterised by the growth of endometrial tissue outside the uterus, causing varying symptoms and having far reaching physical, psychological, social, and economic impacts; however, the characteristics of women with a diagnosis in England are not known.

**Methods:** Using a retrospective cohort design, we linked Hospital Episode Statistics (HES) to sociodemographic information from Census 2011, providing individual-level self-reported characteristics. Our main outcome of interest was a primary diagnosis of endometriosis in an NHS hospital in England. Our exposures were a range of socio-economic and geographical factors self-reported from Census such as age group, and Index of Multiple Deprivation (IMD). We calculated crude and age-standardised rates per 100,000 people, and odds of receiving a diagnosis in an NHS hospital between 27 March 2011 and 31 December 2021 using logistic regression models adjusted for age and health.

**Results:** We estimated the underlying rate of diagnosed endometriosis to be approximately 2% of reproductive age women, with an average age at first diagnosis of 35 years (IQR: 27-43 years). Compared with the White ethnic group, all other ethnic groups had significantly lower odds of diagnosis (Black/African/Caribbean/Black British: OR=0.59; 95%CI=0.57-0.62, Other ethnic group: OR=0.71; 95%CI=0.67-0.76, Asian/Asian British: OR=0.73; 95%CI=0.72-0.75, Mixed/Multiple ethnic groups: OR=0.90, 95%CI=0.87-0.94). Women living in the most and least deprived areas were least likely to have an endometriosis diagnosis in hospital, possibly reflecting lower access to healthcare services in the most deprived group and more use of private healthcare in the least deprived group.

**Conclusions:** Our results demonstrate significant sociodemographic differences between groups of women receiving an endometriosis diagnosis in an NHS hospital in England which should be inform healthcare policies to better support groups of women most affected by endometriosis; subsequent work should explore presentations in primary care, as well as the broader socioeconomic ramifications of endometriosis.

**Research in context:** *What is already known on this topic:* Endometriosis is a common gynaecological condition which has debilitating impacts across many domains, including physical, psychological, social and economic. It is estimated to affect one in ten reproductive age women in England, however evidence on the differences in endometriosis diagnosis by sociodemographic characteristics is lacking.

*What this study adds:* Our study utilises population-level Census and HES data for England to estimate crude and age-standardised rates of diagnosed endometriosis per 100,000 people, and odds of being diagnosed with endometriosis in an NHS hospital in England between 27 March 2011 and 31 December 2021 by a range of sociodemographic characteristics. We estimate the underlying rate of diagnosed endometriosis in an NHS hospital to be approximately 2% of reproductive age women in our linked population, with an average age at first diagnosis of 35 years. Women living in the most and least deprived areas were least likely to have an endometriosis diagnosis; this possibly reflects less access to healthcare services in the most deprived group and more use of private healthcare in the least deprived group. Women in the White ethnic group had significantly higher odds of diagnosis compared with all other ethnic groups. Women self-reporting to be in bad health, or limited in their day-to-day activities, were more likely to have been diagnosed with endometriosis in an NHS hospital compared with those in very good health or non-disabled women, respectively. This study is the most comprehensive analysis of the characteristics of women with an endometriosis diagnosis in England to date.

*How this study might affect research, practice or policy:* This research provides important information to gynaecologists, clinicians and other allied health professionals, as well as policy makers, to illustrate the underlying rate of diagnosed endometriosis in NHS hospitals, the groups most affected by endometriosis, and barriers to receiving a diagnosis. In the 2022 Women’s Health Strategy for England, menstrual health and gynaecological conditions were identified as one of the priority areas, with a call for evidence and investment in women’s health research.

## Introduction

Endometriosis is a chronic disease and the second most common gynaecological condition in the UK, estimated to impact approximately 1.5 million women (1). Endometriosis is a condition where endometrial tissue, similar to the lining of the uterus, grows in other places, such as the ovaries and fallopian tubes. Common symptoms include chronic pelvic pain, fatigue, heavy menstrual bleeding, pain during or after sex, painful urination and bowel movements, and reduced fertility (2,3). Symptoms can vary across women, and severity of symptoms often does not reflect the stage of the disease (4). Endometriosis usually affects women during their reproductive years (between menarche and menopause) but can affect women of any age (5). While the exact cause of endometriosis is unknown, several factors have been implicated in its development, including immune, endocrine, genetic, and environmental influences (3,6).

Despite the profound impacts endometriosis poses on women’s lives, there has been no population study in England assessing the characteristics of women living with endometriosis. Given the disease is known to have negative impacts across many domains, such as psychological, social, and economic, understanding which groups of women are most affected is imperative to further quantify differences in the progression and burden of this disease, and inform targeted work to reduce these inequalities. This has ramifications clinically, for understanding which groups are most affected, as well as from a policy perspective to provide evidence-based change to targeted populations.

Diagnosing endometriosis can be challenging due to the non-specific nature of its symptoms, the overlap of symptoms with other conditions such as pelvic inflammatory disease or irritable bowel syndrome, biases within the healthcare system (7), and the lack of a definitive non-invasive diagnostic test. In England, it takes on average eight years from onset of symptoms to receiving a diagnosis (8). The gold standard for diagnosis is in secondary care after laparoscopic visualisation and biopsy of the lesions (9). According to the National Institute for Health and Care Excellence (NICE) guidelines, a diagnosis of endometriosis can only be made definitively following a laparoscopy. Women may have previously presented with symptoms of endometriosis in primary care, however a General Practitioner (GP) cannot diagnose endometriosis for certain and should refer women to a specialist service in an NHS hospital (9,10). In the current study, we utilise individual population-level hospital inpatient data from Hospital Episode Statistics (HES) (11) for England to define a cohort of women who received a diagnosis of endometriosis in a National Health Service (NHS) hospital over a ten-year period. Using Census 2011 data we estimated differences in odds of diagnosis by sociodemographic characteristics including ethnic group, socioeconomic position, education level, country of birth, health, disability and region. This is of important public health significance due to the prevalence of this condition, its debilitating symptoms, and the broader socioeconomic burden of the disease.

## Methods

### Study design and data sources

We conducted a population-level retrospective cohort study using the Public Health Data Asset 2011 Cohort (12), which combines Census 2011 data, death registrations, and 2009-2021 Hospital Episode Statistics (HES) Admitted Patient Care (APC) data (13). Census 2011 has been linked to the 2011–2013 NHS Patient Registers to obtain NHS numbers, with a linkage rate of 94.6% (12). The decennial Census for England and Wales captured population and household characteristics of 56 million people in 2011 with respondents reporting detailed demographic information. The HES APC data capture records of patients admitted for treatment at NHS hospitals in England. Each HES record includes up to 20 diagnosis values, with the first value (primary diagnosis) recording the main condition being treated or investigated, and the other values recording any relevant secondary/subsidiary diagnoses (14).

### Study population

The study population includes all women enumerated in the 2011 Census who were usual residents in England on Census Day (27 March 2011) and could be linked to an NHS number [Supplementary Table 1]. We defined two cohorts of women with endometriosis. For our main analysis, we estimated incidence of endometriosis diagnosis in hospital by identifying a group of women with a primary diagnosis of endometriosis in an NHS hospital during the study period between 27 March 2011 and 31 December 2021 and used two years of HES data (1 April 2009 to 26 March 2011) prior to the study start to exclude instances where the hospital diagnosis of endometriosis during the study period was not the first. The control group of women for the main analysis were enumerated in Census 2011 and had no primary or secondary diagnosis of endometriosis in hospital during the study period, or in the two years prior. For our secondary analysis, we estimated the underlying rate of diagnosed endometriosis in hospital by identifying women with any diagnosis of endometriosis (primary or secondary) in an NHS hospital during the study period and a control group of women enumerated in Census 2011 with no evidence of a primary or secondary diagnosis of endometriosis in hospital during the study period. All groups were filtered to restrict our cohort to people who self-reported as female in the 2011 Census [Supplementary Table 1].

**Table 1:**
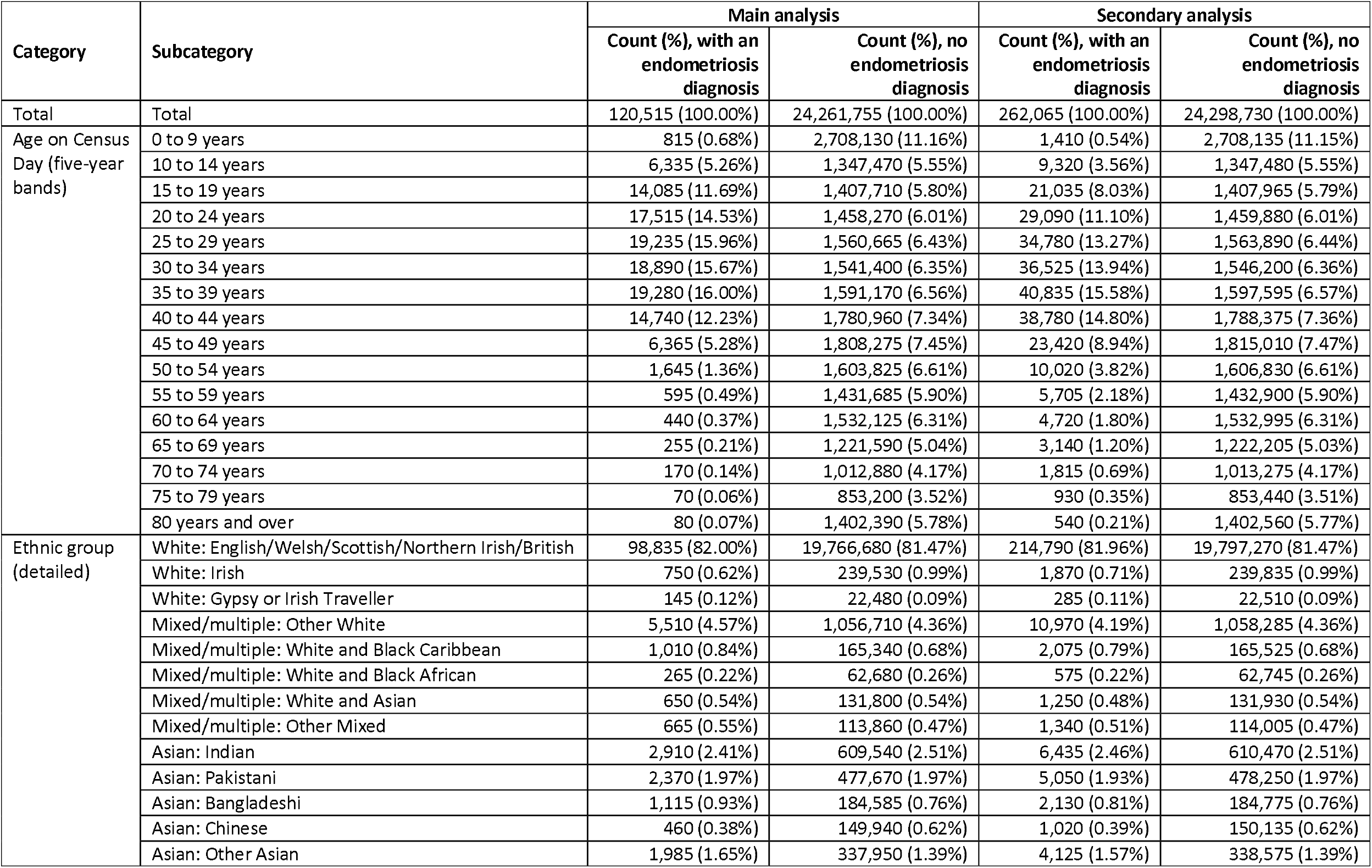
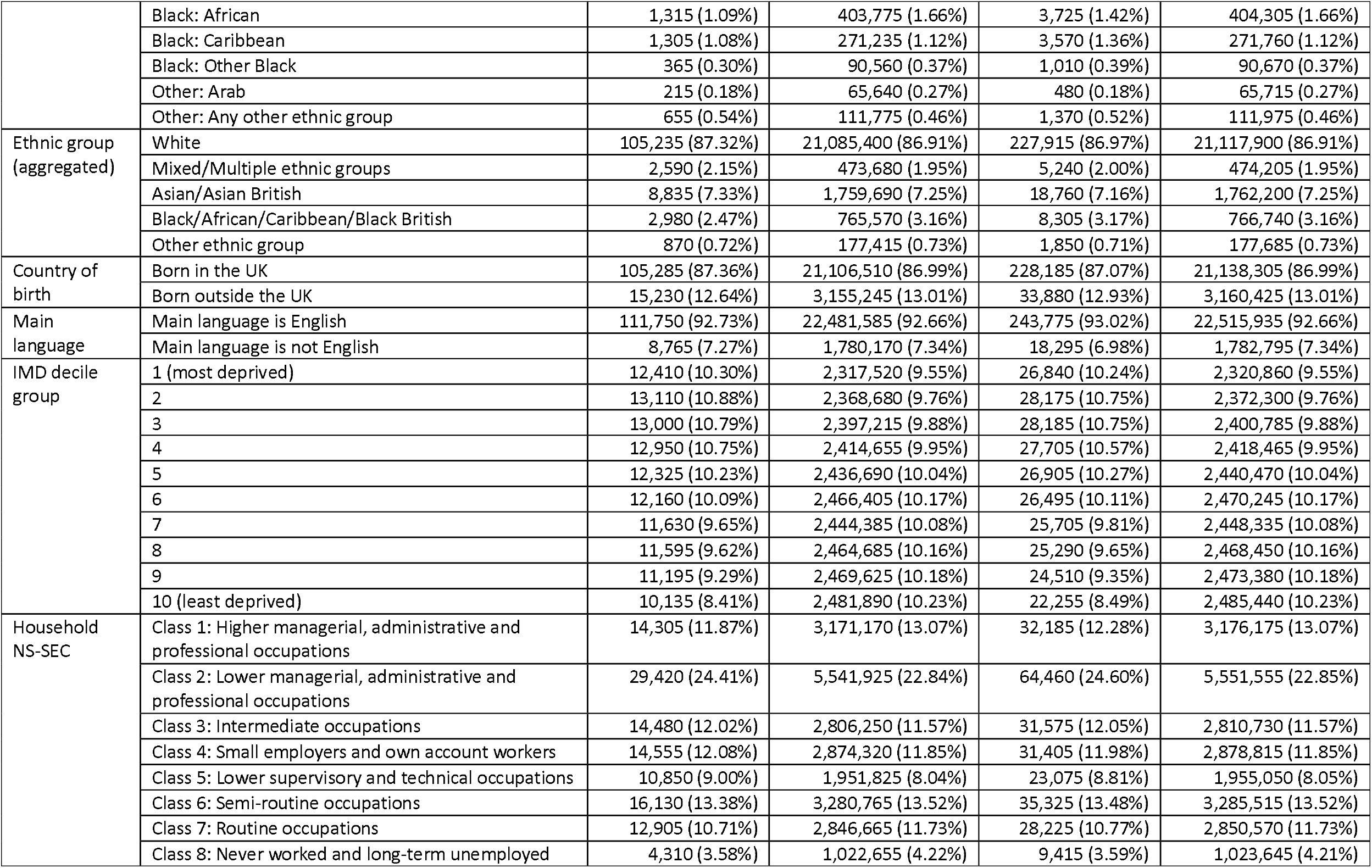
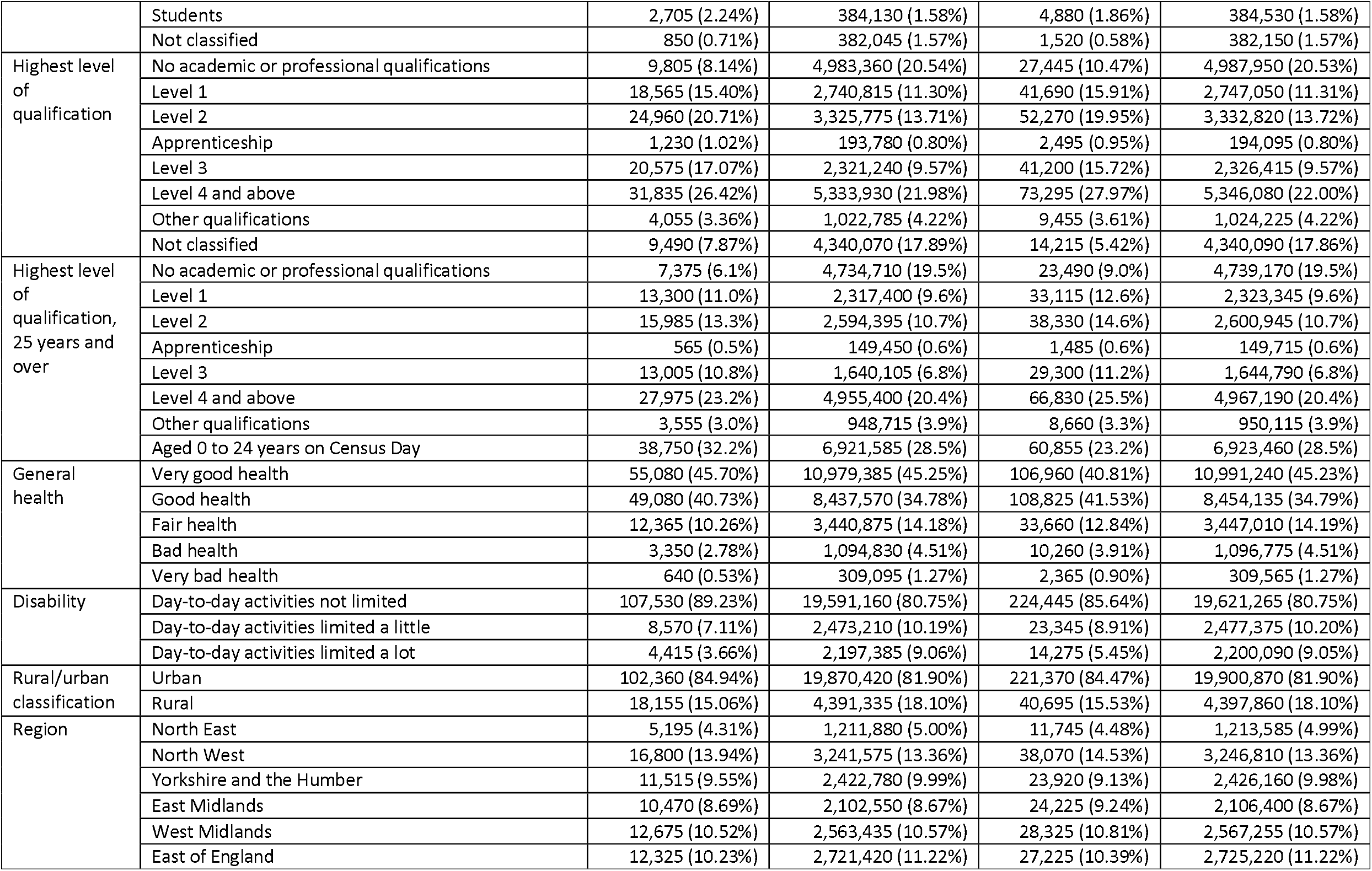
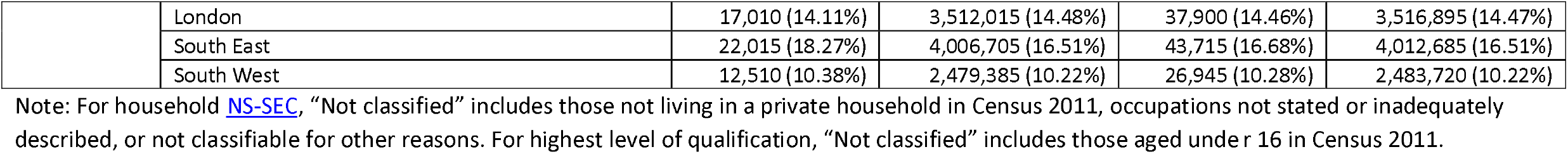
Characteristics of the study population.

| Category | Subcategory | Main analysis |  | Secondary analysis |  |
| --- | --- | --- | --- | --- | --- |
|  |  | Count (%), with an endometriosis diagnosis | Count (%), no endometriosis diagnosis | Count (%), with an endometriosis diagnosis | Count (%), no endometriosis diagnosis |
| Total | Total | 120,515 (100.00%) | 24,261,755 (100.00%) | 262,065 (100.00%) | 24,298,730 (100.00%) |
| Age on Census Day (five-year bands) | 0 to 9 years | 815 (0.68%) | 2,708,130 (11.16%) | 1,410 (0.54%) | 2,708,135 (11.15%) |
|  | 10 to 14 years | 6,335 (5.26%) | 1,347,470 (5.55%) | 9,320 (3.56%) | 1,347,480 (5.55%) |
|  | 15 to 19 years | 14,085 (11.69%) | 1,407,710 (5.80%) | 21,035 (8.03%) | 1,407,965 (5.79%) |
|  | 20 to 24 years | 17,515 (14.53%) | 1,458,270 (6.01%) | 29,090 (11.10%) | 1,459,880 (6.01%) |
|  | 25 to 29 years | 19,235 (15.96%) | 1,560,665 (6.43%) | 34,780 (13.27%) | 1,563,890 (6.44%) |
|  | 30 to 34 years | 18,890 (15.67%) | 1,541,400 (6.35%) | 36,525 (13.94%) | 1,546,200 (6.36%) |
|  | 35 to 39 years | 19,280 (16.00%) | 1,591,170 (6.56%) | 40,835 (15.58%) | 1,597,595 (6.57%) |
|  | 40 to 44 years | 14,740 (12.23%) | 1,780,960 (7.34%) | 38,780 (14.80%) | 1,788,375 (7.36%) |
|  | 45 to 49 years | 6,365 (5.28%) | 1,808,275 (7.45%) | 23,420 (8.94%) | 1,815,010 (7.47%) |
|  | 50 to 54 years | 1,645 (1.36%) | 1,603,825 (6.61%) | 10,020 (3.82%) | 1,606,830 (6.61%) |
|  | 55 to 59 years | 595 (0.49%) | 1,431,685 (5.90%) | 5,705 (2.18%) | 1,432,900 (5.90%) |
|  | 60 to 64 years | 440 (0.37%) | 1,532,125 (6.31%) | 4,720 (1.80%) | 1,532,995 (6.31%) |
|  | 65 to 69 years | 255 (0.21%) | 1,221,590 (5.04%) | 3,140 (1.20%) | 1,222,205 (5.03%) |
|  | 70 to 74 years | 170 (0.14%) | 1,012,880 (4.17%) | 1,815 (0.69%) | 1,013,275 (4.17%) |
|  | 75 to 79 years | 70 (0.06%) | 853,200 (3.52%) | 930 (0.35%) | 853,440 (3.51%) |
|  | 80 years and over | 80 (0.07%) | 1,402,390 (5.78%) | 540 (0.21%) | 1,402,560 (5.77%) |
| Ethnic group (detailed) | White: English/Welsh/Scottish/Northern Irish/British | 98,835 (82.00%) | 19,766,680 (81.47%) | 214,790 (81.96%) | 19,797,270 (81.47%) |
|  | White: Irish | 750 (0.62%) | 239,530 (0.99%) | 1,870 (0.71%) | 239,835 (0.99%) |
|  | White: Gypsy or Irish Traveller | 145 (0.12%) | 22,480 (0.09%) | 285 (0.11%) | 22,510 (0.09%) |
|  | Mixed/multiple: Other White | 5,510 (4.57%) | 1,056,710 (4.36%) | 10,970 (4.19%) | 1,058,285 (4.36%) |
|  | Mixed/multiple: White and Black Caribbean | 1,010 (0.84%) | 165,340 (0.68%) | 2,075 (0.79%) | 165,525 (0.68%) |
|  | Mixed/multiple: White and Black African | 265 (0.22%) | 62,680 (0.26%) | 575 (0.22%) | 62,745 (0.26%) |
|  | Mixed/multiple: White and Asian | 650 (0.54%) | 131,800 (0.54%) | 1,250 (0.48%) | 131,930 (0.54%) |
|  | Mixed/multiple: Other Mixed | 665 (0.55%) | 113,860 (0.47%) | 1,340 (0.51%) | 114,005 (0.47%) |
|  | Asian: Indian | 2,910 (2.41%) | 609,540 (2.51%) | 6,435 (2.46%) | 610,470 (2.51%) |
|  | Asian: Pakistani | 2,370 (1.97%) | 477,670 (1.97%) | 5,050 (1.93%) | 478,250 (1.97%) |
|  | Asian: Bangladeshi | 1,115 (0.93%) | 184,585 (0.76%) | 2,130 (0.81%) | 184,775 (0.76%) |
|  | Asian: Chinese | 460 (0.38%) | 149,940 (0.62%) | 1,020 (0.39%) | 150,135 (0.62%) |
|  | Asian: Other Asian | 1,985 (1.65%) | 337,950 (1.39%) | 4,125 (1.57%) | 338,575 (1.39%) |
|  | Black: African | 1,315 (1.09%) | 403,775 (1.66%) | 3,725 (1.42%) | 404,305 (1.66%) |
|  | Black: Caribbean | 1,305 (1.08%) | 271,235 (1.12%) | 3,570 (1.36%) | 271,760 (1.12%) |
|  | Black: Other Black | 365 (0.30%) | 90,560 (0.37%) | 1,010 (0.39%) | 90,670 (0.37%) |
|  | Other: Arab | 215 (0.18%) | 65,640 (0.27%) | 480 (0.18%) | 65,715 (0.27%) |
|  | Other: Any other ethnic group | 655 (0.54%) | 111,775 (0.46%) | 1,370 (0.52%) | 111,975 (0.46%) |
| Ethnic group<br>(aggregated) | White | 105,235 (87.32%) | 21,085,400 (86.91%) | 227,915 (86.97%) | 21,117,900 (86.91%) |
|  | Mixed/Multiple ethnic groups | 2,590 (2.15%) | 473,680 (1.95%) | 5,240 (2.00%) | 474,205 (1.95%) |
|  | Asian/Asian British | 8,835 (7.33%) | 1,759,690 (7.25%) | 18,760 (7.16%) | 1,762,200 (7.25%) |
|  | Black/African/Caribbean/Black British | 2,980 (2.47%) | 765,570 (3.16%) | 8,305 (3.17%) | 766,740 (3.16%) |
|  | Other ethnic group | 870 (0.72%) | 177,415 (0.73%) | 1,850 (0.71%) | 177,685 (0.73%) |
| Country of<br>birth | Born in the UK | 105,285 (87.36%) | 21,106,510 (86.99%) | 228,185 (87.07%) | 21,138,305 (86.99%) |
|  | Born outside the UK | 15,230 (12.64%) | 3,155,245 (13.01%) | 33,880 (12.93%) | 3,160,425 (13.01%) |
| Main<br>language | Main language is English | 111,750 (92.73%) | 22,481,585 (92.66%) | 243,775 (93.02%) | 22,515,935 (92.66%) |
|  | Main language is not English | 8,765 (7.27%) | 1,780,170 (7.34%) | 18,295 (6.98%) | 1,782,795 (7.34%) |
| IMD decile<br>group | 1 (most deprived) | 12,410 (10.30%) | 2,317,520 (9.55%) | 26,840 (10.24%) | 2,320,860 (9.55%) |
|  | 2 | 13,110 (10.88%) | 2,368,680 (9.76%) | 28,175 (10.75%) | 2,372,300 (9.76%) |
|  | 3 | 13,000 (10.79%) | 2,397,215 (9.88%) | 28,185 (10.75%) | 2,400,785 (9.88%) |
|  | 4 | 12,950 (10.75%) | 2,414,655 (9.95%) | 27,705 (10.57%) | 2,418,465 (9.95%) |
|  | 5 | 12,325 (10.23%) | 2,436,690 (10.04%) | 26,905 (10.27%) | 2,440,470 (10.04%) |
|  | 6 | 12,160 (10.09%) | 2,466,405 (10.17%) | 26,495 (10.11%) | 2,470,245 (10.17%) |
|  | 7 | 11,630 (9.65%) | 2,444,385 (10.08%) | 25,705 (9.81%) | 2,448,335 (10.08%) |
|  | 8 | 11,595 (9.62%) | 2,464,685 (10.16%) | 25,290 (9.65%) | 2,468,450 (10.16%) |
|  | 9 | 11,195 (9.29%) | 2,469,625 (10.18%) | 24,510 (9.35%) | 2,473,380 (10.18%) |
|  | 10 (least deprived) | 10,135 (8.41%) | 2,481,890 (10.23%) | 22,255 (8.49%) | 2,485,440 (10.23%) |
| Household<br>NS-SEC | Class 1: Higher managerial, administrative and professional occupations | 14,305 (11.87%) | 3,171,170 (13.07%) | 32,185 (12.28%) | 3,176,175 (13.07%) |
|  | Class 2: Lower managerial, administrative and professional occupations | 29,420 (24.41%) | 5,541,925 (22.84%) | 64,460 (24.60%) | 5,551,555 (22.85%) |
|  | Class 3: Intermediate occupations | 14,480 (12.02%) | 2,806,250 (11.57%) | 31,575 (12.05%) | 2,810,730 (11.57%) |
|  | Class 4: Small employers and own account workers | 14,555 (12.08%) | 2,874,320 (11.85%) | 31,405 (11.98%) | 2,878,815 (11.85%) |
|  | Class 5: Lower supervisory and technical occupations | 10,850 (9.00%) | 1,951,825 (8.04%) | 23,075 (8.81%) | 1,955,050 (8.05%) |
|  | Class 6: Semi-routine occupations | 16,130 (13.38%) | 3,280,765 (13.52%) | 35,325 (13.48%) | 3,285,515 (13.52%) |
|  | Class 7: Routine occupations | 12,905 (10.71%) | 2,846,665 (11.73%) | 28,225 (10.77%) | 2,850,570 (11.73%) |
|  | Class 8: Never worked and long-term unemployed | 4,310 (3.58%) | 1,022,655 (4.22%) | 9,415 (3.59%) | 1,023,645 (4.21%) |
|  | Students | 2,705 (2.24%) | 384,130 (1.58%) | 4,880 (1.86%) | 384,530 (1.58%) |
|  | Not classified | 850 (0.71%) | 382,045 (1.57%) | 1,520 (0.58%) | 382,150 (1.57%) |
| Highest level of qualification | No academic or professional qualifications | 9,805 (8.14%) | 4,983,360 (20.54%) | 27,445 (10.47%) | 4,987,950 (20.53%) |
|  | Level 1 | 18,565 (15.40%) | 2,740,815 (11.30%) | 41,690 (15.91%) | 2,747,050 (11.31%) |
|  | Level 2 | 24,960 (20.71%) | 3,325,775 (13.71%) | 52,270 (19.95%) | 3,332,820 (13.72%) |
|  | Apprenticeship | 1,230 (1.02%) | 193,780 (0.80%) | 2,495 (0.95%) | 194,095 (0.80%) |
|  | Level 3 | 20,575 (17.07%) | 2,321,240 (9.57%) | 41,200 (15.72%) | 2,326,415 (9.57%) |
|  | Level 4 and above | 31,835 (26.42%) | 5,333,930 (21.98%) | 73,295 (27.97%) | 5,346,080 (22.00%) |
|  | Other qualifications | 4,055 (3.36%) | 1,022,785 (4.22%) | 9,455 (3.61%) | 1,024,225 (4.22%) |
|  | Not classified | 9,490 (7.87%) | 4,340,070 (17.89%) | 14,215 (5.42%) | 4,340,090 (17.86%) |
| Highest level of qualification, 25 years and over | No academic or professional qualifications | 7,375 (6.1%) | 4,734,710 (19.5%) | 23,490 (9.0%) | 4,739,170 (19.5%) |
|  | Level 1 | 13,300 (11.0%) | 2,317,400 (9.6%) | 33,115 (12.6%) | 2,323,345 (9.6%) |
|  | Level 2 | 15,985 (13.3%) | 2,594,395 (10.7%) | 38,330 (14.6%) | 2,600,945 (10.7%) |
|  | Apprenticeship | 565 (0.5%) | 149,450 (0.6%) | 1,485 (0.6%) | 149,715 (0.6%) |
|  | Level 3 | 13,005 (10.8%) | 1,640,105 (6.8%) | 29,300 (11.2%) | 1,644,790 (6.8%) |
|  | Level 4 and above | 27,975 (23.2%) | 4,955,400 (20.4%) | 66,830 (25.5%) | 4,967,190 (20.4%) |
|  | Other qualifications | 3,555 (3.0%) | 948,715 (3.9%) | 8,660 (3.3%) | 950,115 (3.9%) |
|  | Aged 0 to 24 years on Census Day | 38,750 (32.2%) | 6,921,585 (28.5%) | 60,855 (23.2%) | 6,923,460 (28.5%) |
| General health | Very good health | 55,080 (45.70%) | 10,979,385 (45.25%) | 106,960 (40.81%) | 10,991,240 (45.23%) |
|  | Good health | 49,080 (40.73%) | 8,437,570 (34.78%) | 108,825 (41.53%) | 8,454,135 (34.79%) |
|  | Fair health | 12,365 (10.26%) | 3,440,875 (14.18%) | 33,660 (12.84%) | 3,447,010 (14.19%) |
|  | Bad health | 3,350 (2.78%) | 1,094,830 (4.51%) | 10,260 (3.91%) | 1,096,775 (4.51%) |
|  | Very bad health | 640 (0.53%) | 309,095 (1.27%) | 2,365 (0.90%) | 309,565 (1.27%) |
| Disability | Day-to-day activities not limited | 107,530 (89.23%) | 19,591,160 (80.75%) | 224,445 (85.64%) | 19,621,265 (80.75%) |
|  | Day-to-day activities limited a little | 8,570 (7.11%) | 2,473,210 (10.19%) | 23,345 (8.91%) | 2,477,375 (10.20%) |
|  | Day-to-day activities limited a lot | 4,415 (3.66%) | 2,197,385 (9.06%) | 14,275 (5.45%) | 2,200,090 (9.05%) |
| Rural/urban classification | Urban | 102,360 (84.94%) | 19,870,420 (81.90%) | 221,370 (84.47%) | 19,900,870 (81.90%) |
|  | Rural | 18,155 (15.06%) | 4,391,335 (18.10%) | 40,695 (15.53%) | 4,397,860 (18.10%) |
| Region | North East | 5,195 (4.31%) | 1,211,880 (5.00%) | 11,745 (4.48%) | 1,213,585 (4.99%) |
|  | North West | 16,800 (13.94%) | 3,241,575 (13.36%) | 38,070 (14.53%) | 3,246,810 (13.36%) |
|  | Yorkshire and the Humber | 11,515 (9.55%) | 2,422,780 (9.99%) | 23,920 (9.13%) | 2,426,160 (9.98%) |
|  | East Midlands | 10,470 (8.69%) | 2,102,550 (8.67%) | 24,225 (9.24%) | 2,106,400 (8.67%) |
|  | West Midlands | 12,675 (10.52%) | 2,563,435 (10.57%) | 28,325 (10.81%) | 2,567,255 (10.57%) |
|  | East of England | 12,325 (10.23%) | 2,721,420 (11.22%) | 27,225 (10.39%) | 2,725,220 (11.22%) |
|  | London | 17,010 (14.11%) | 3,512,015 (14.48%) | 37,900 (14.46%) | 3,516,895 (14.47%) |
|  | South East | 22,015 (18.27%) | 4,006,705 (16.51%) | 43,715 (16.68%) | 4,012,685 (16.51%) |
|  | South West | 12,510 (10.38%) | 2,479,385 (10.22%) | 26,945 (10.28%) | 2,483,720 (10.22%) |

Note: For household NS-SEC, “Not classified” includes those not living in a private household in Census 2011, occupations not stated or inadequately described, or not classifiable for other reasons. For highest level of qualification, “Not classified” includes those aged under 16 in Census 2011.

### Outcome

Diagnosis of endometriosis was defined from the HES Admitted Patient Care (APC) data using International Classification of Diseases, Tenth Revision (ICD-10) codes N80.0-N80.9 [Supplementary Table 2]. For our main analysis, an outcome was identified if at least one diagnosis of endometriosis, recorded as a primary diagnosis, was recorded during the study period. For the secondary analysis, an outcome was identified if at least one diagnosis of endometriosis, recorded as either a primary or secondary diagnosis, was recorded during the study period. An episode must have started and ended within our study period for an outcome to be identified. Outcomes were coded as binary.

**Table 2:**
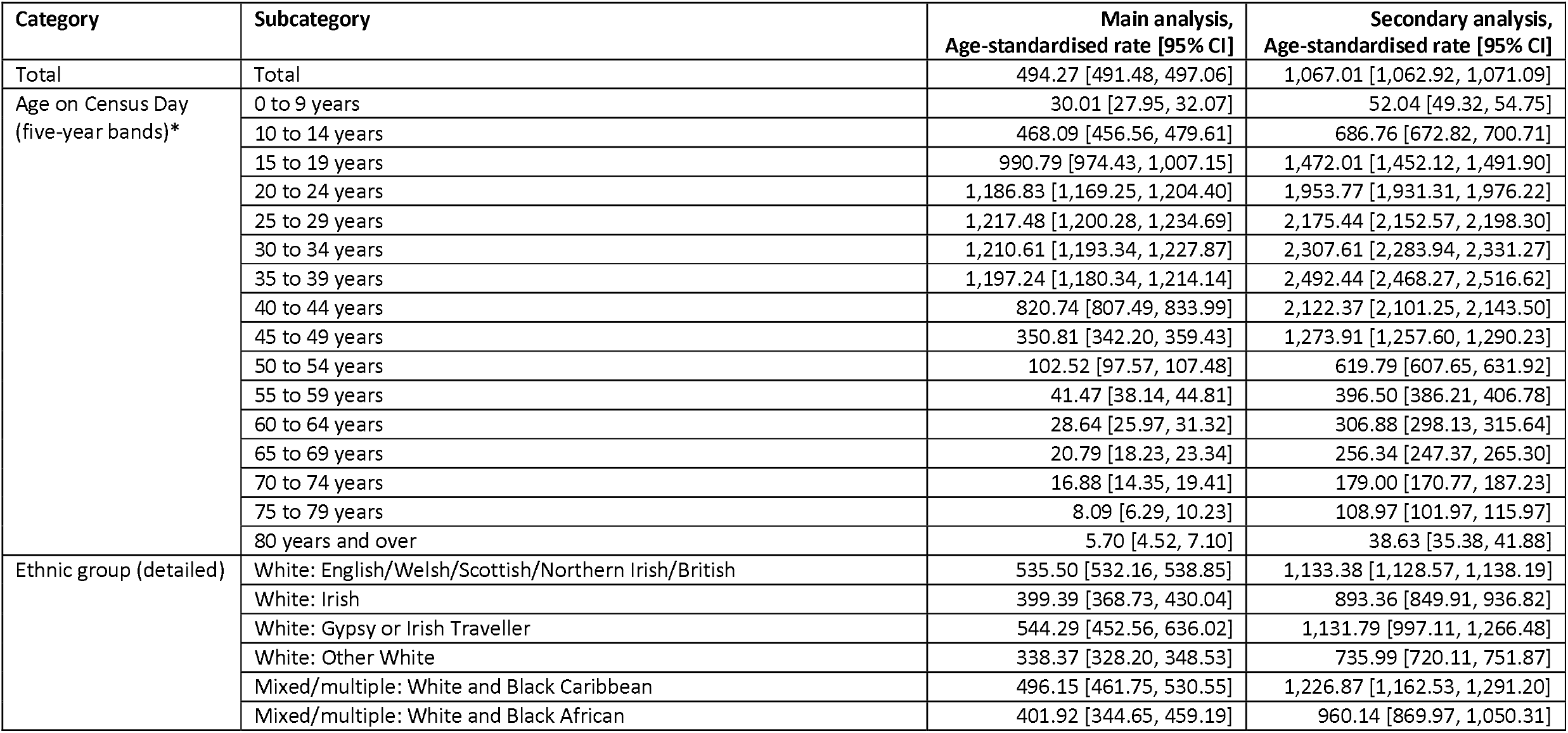
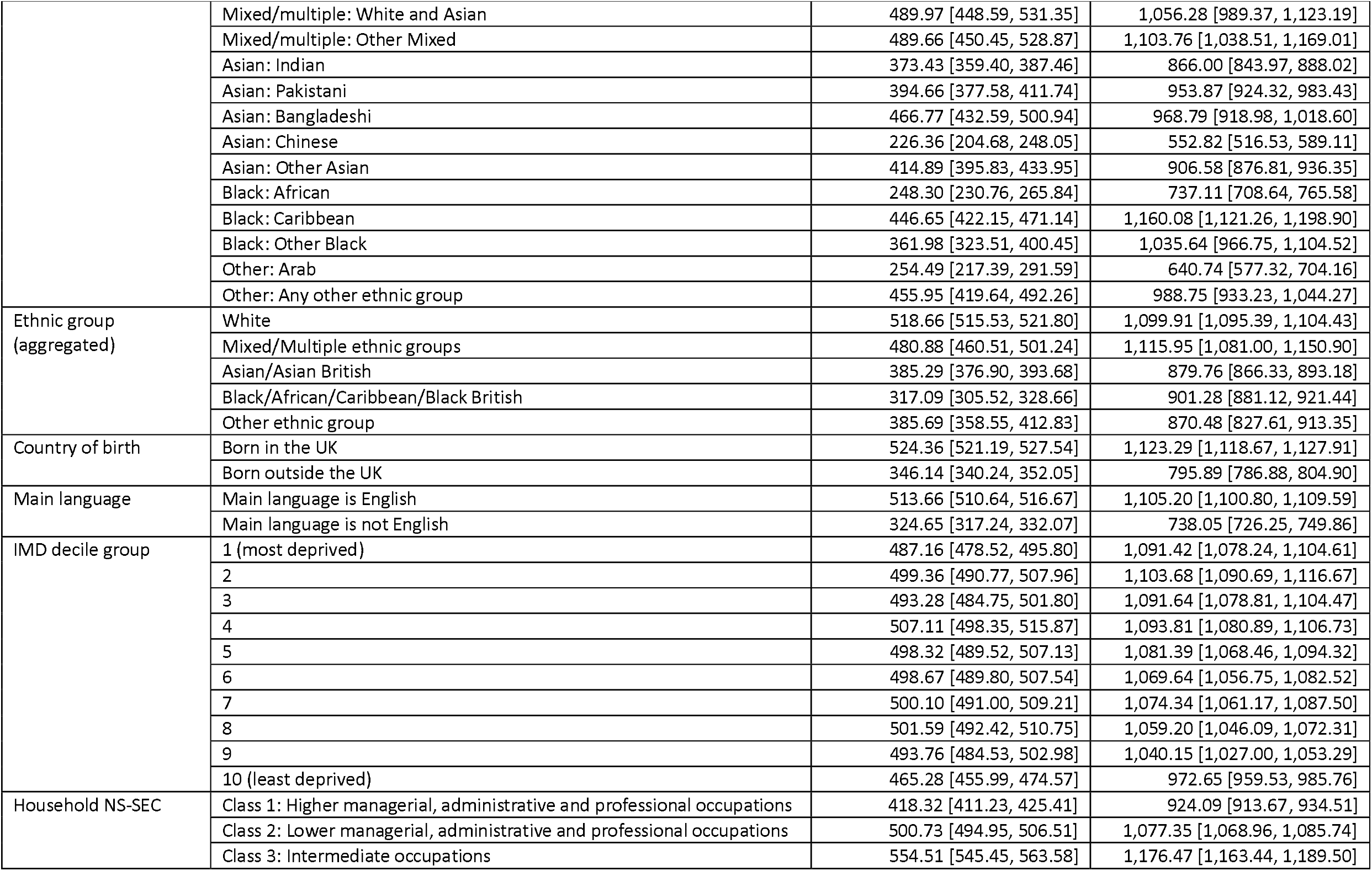
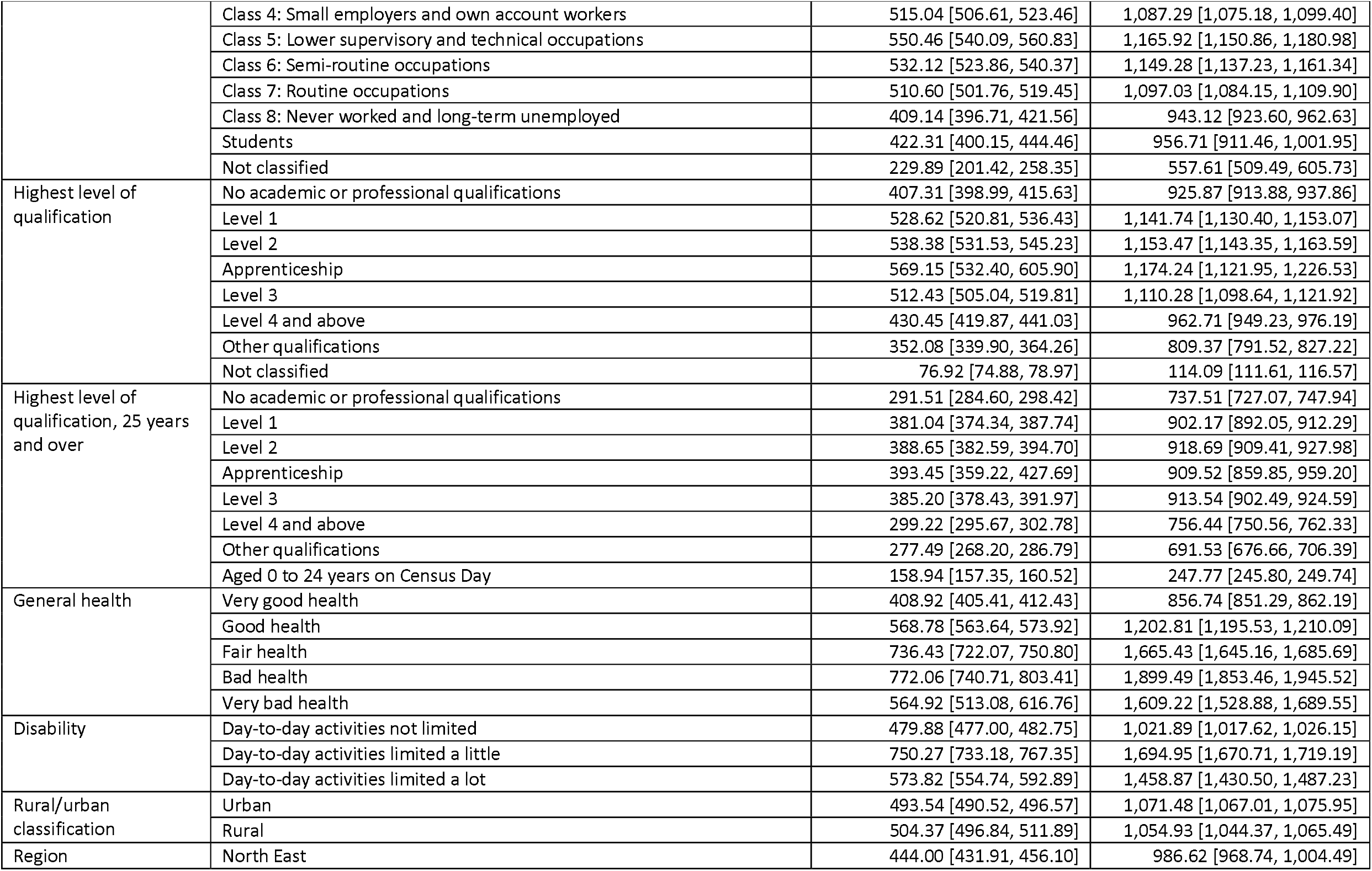
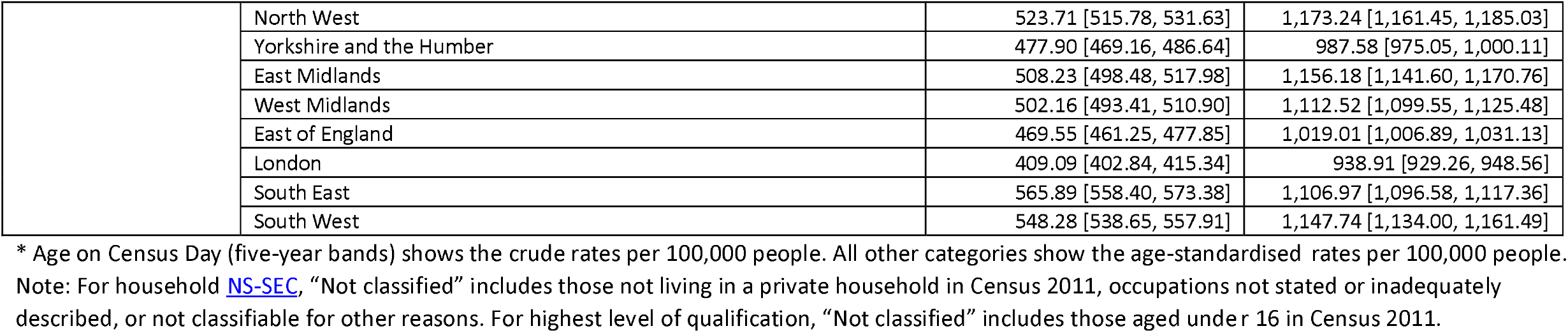
Age-standardised rates of endometriosis diagnosis per 100,000 people.

### Covariates

All sociodemographic variables included in the analyses were self-reported in the 2011 Census. Exposures included were age on Census Day (five-year age bands), ethnic group (detailed and aggregated), country of birth, main language, Index of Multiple Deprivation (IMD) decile, National Statistics Socioeconomic Classification (NS-SEC) of the Household Reference Person (HRP), highest level of educational qualification, general health, disability, rural/urban, region and upper tier local authority (UTLA). Groupings of all exposures can be seen in Supplementary Table 3. Where Census data was not applicable for a particular group, a “Not classified” group was included. Missing Census 2011 items were imputed using nearest neighbour donor imputation, the standard method used by the Office for National Statistics (ONS) to impute missing values (15). Importantly, at point of completion of 2011 Census all questions, except religious affiliation, were mandatory.

**Table 3:**
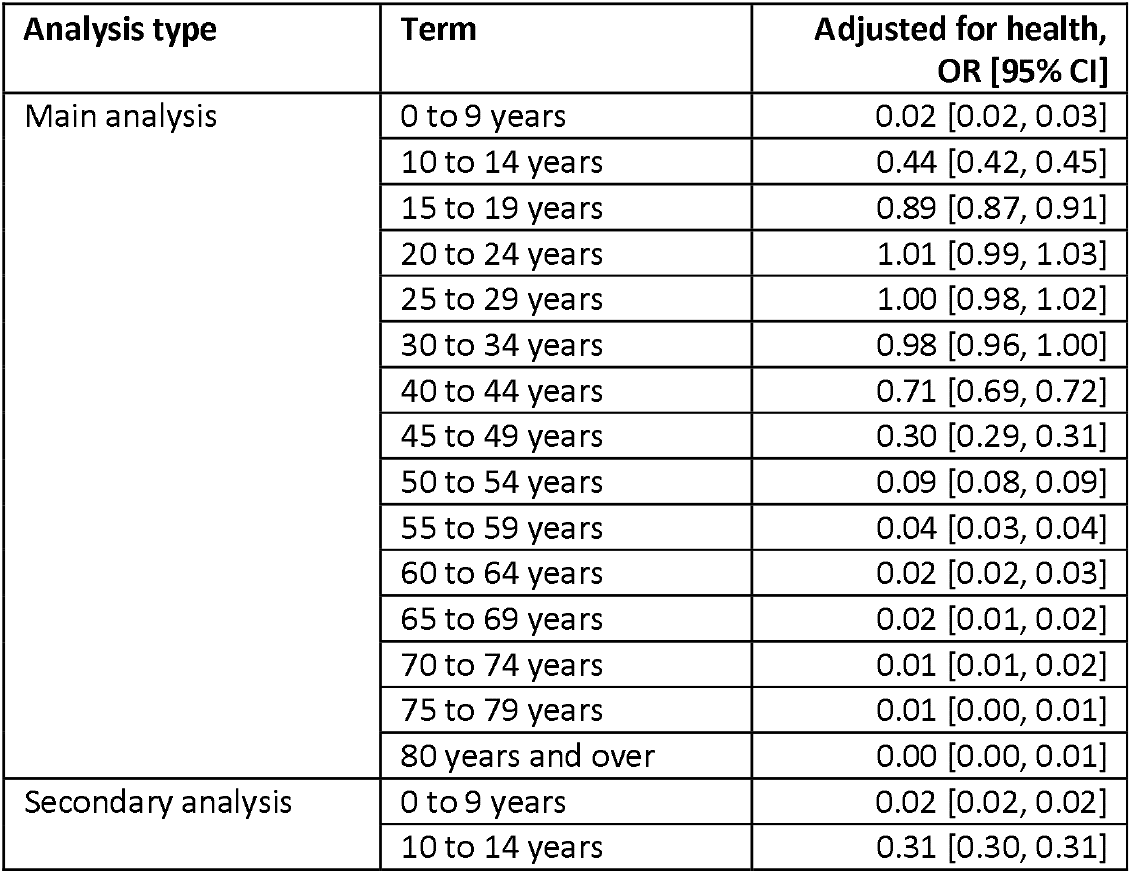
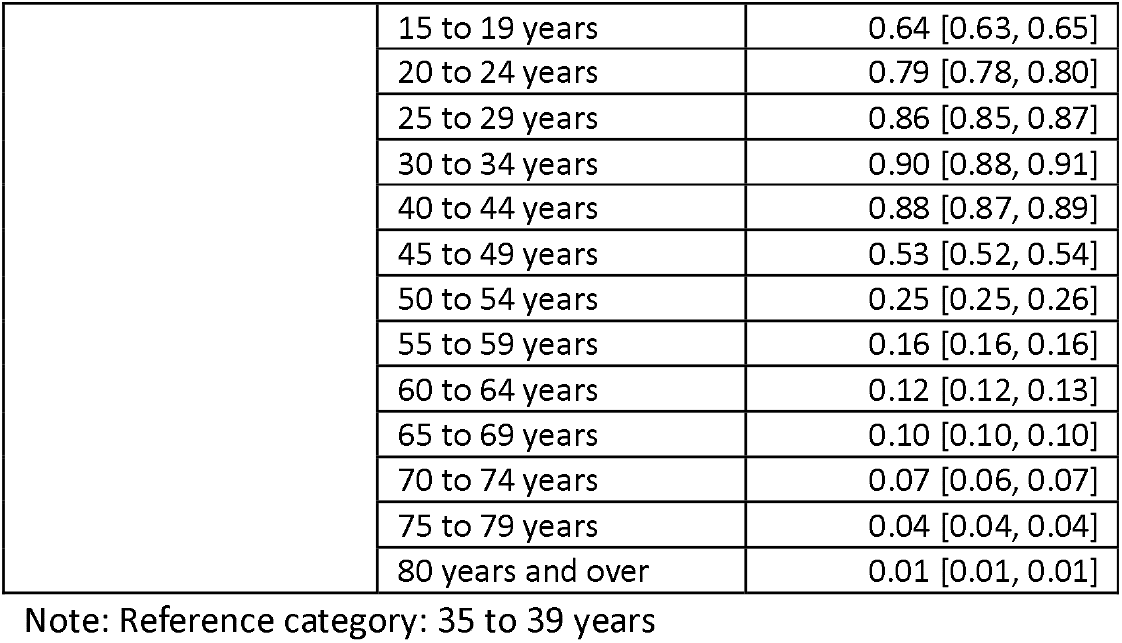
Odds ratios for endometriosis diagnosis and 95% confidence intervals by age on Census Day (five-year bands)

An indicator of health was determined using number of HES APC admissions in the two years prior to our study start date (1 April 2009 to 26 March 2011) for any reason, excluding mentions of endometriosis. Number of HES APC admissions were grouped into 0 episodes, 1 to 3 episodes, 4 to 6 episodes, 7 to 9 episodes, 10 to 14 episodes, and 15 or more episodes. We did not include self-reported health or self-reported disability status from 2011 Census as health adjustments in the models, as these variables could capture effects from symptoms related to endometriosis.

In our descriptive analysis, we investigated age at endometriosis diagnosis and method of hospital admission. Age at endometriosis diagnosis refers to age at first diagnosis during the study period and was obtained from the HES APC data, but if age was missing or zero (0.01%) in the administrative data it was calculated using the date difference between the date of birth self-reported in Census 2011 and the hospital episode start date. Method of hospital admission (emergency or non-emergency) was sourced from the HES APC data. Emergency admissions are defined in HES as “unpredictable and at short notice because of clinical need” (14), and were classified according to the codes listed in Supplementary Table 4.

**Table 4:** Odds ratios for endometriosis diagnosis and 95% confidence intervals by ethnic group.

| Analysis type | Exposure | Term | Adjusted for age,<br>OR [95% CI] | Adjusted for age<br>and health,<br>OR [95% CI] | Adjusted for age,<br>health and<br>country of birth,<br>OR [95% CI] | Adjusted for age,<br>health and main<br>language,<br>OR [95% CI] | Adjusted for age,<br>health, country<br>of birth and<br>main language,<br>OR [95% CI] |
| --- | --- | --- | --- | --- | --- | --- | --- |
| Main analysis | Ethnic group<br>(detailed)<br>(Reference<br>category: White:<br>English/Welsh/Sc<br>ottish/Northern<br>Irish/British) | White: Irish | 0.77 [0.72, 0.83] | 0.79 [0.74, 0.85] | 0.84 [0.78, 0.90] | 0.79 [0.74, 0.85] | 0.82 [0.76, 0.89] |
|  |  | White: Gypsy or Irish<br>Traveller | 0.98 [0.83, 1.16] | 0.93 [0.79, 1.10] | 0.94 [0.80, 1.11] | 0.95 [0.80, 1.12] | 0.95 [0.81, 1.12] |
|  |  | White: Other White | 0.63 [0.61, 0.65] | 0.66 [0.64, 0.68] | 0.73 [0.70, 0.75] | 0.73 [0.70, 0.75] | 0.76 [0.73, 0.79] |
|  |  | Mixed/multiple: White<br>and Black Caribbean | 0.93 [0.88, 0.99] | 0.92 [0.86, 0.97] | 0.92 [0.86, 0.98] | 0.92 [0.86, 0.98] | 0.92 [0.86, 0.98] |
|  |  | Mixed/multiple: White<br>and Black African | 0.73 [0.65, 0.83] | 0.73 [0.65, 0.83] | 0.77 [0.68, 0.87] | 0.76 [0.67, 0.85] | 0.78 [0.69, 0.88] |
|  |  | Mixed/multiple: White<br>and Asian | 0.85 [0.78, 0.91] | 0.86 [0.80, 0.93] | 0.88 [0.82, 0.95] | 0.88 [0.81, 0.95] | 0.89 [0.82, 0.96] |
|  |  | Mixed/multiple: Other<br>Mixed | 0.90 [0.83, 0.97] | 0.91 [0.84, 0.98] | 0.95 [0.88, 1.02] | 0.93 [0.86, 1.01] | 0.95 [0.88, 1.03] |
|  |  | Asian: Indian | 0.69 [0.67, 0.72] | 0.71 [0.69, 0.74] | 0.75 [0.72, 0.78] | 0.74 [0.72, 0.77] | 0.76 [0.73, 0.79] |
|  |  | Asian: Pakistani | 0.70 [0.67, 0.73] | 0.69 [0.66, 0.72] | 0.72 [0.69, 0.75] | 0.73 [0.70, 0.76] | 0.74 [0.71, 0.77] |
|  |  | Asian: Bangladeshi | 0.82 [0.78, 0.87] | 0.82 [0.77, 0.87] | 0.87 [0.82, 0.92] | 0.89 [0.83, 0.94] | 0.91 [0.85, 0.96] |
|  |  | Asian: Chinese | 0.38 [0.34, 0.41] | 0.40 [0.37, 0.44] | 0.44 [0.40, 0.48] | 0.44 [0.40, 0.48] | 0.45 [0.41, 0.50] |
|  |  | Asian: Other Asian | 0.79 [0.76, 0.83] | 0.82 [0.78, 0.85] | 0.89 [0.85, 0.94] | 0.89 [0.85, 0.94] | 0.93 [0.88, 0.98] |
|  |  | Black: African | 0.44 [0.42, 0.46] | 0.43 [0.41, 0.46] | 0.47 [0.45, 0.50] | 0.45 [0.43, 0.48] | 0.48 [0.45, 0.50] |
|  |  | Black: Caribbean | 0.83 [0.79, 0.88] | 0.83 [0.78, 0.87] | 0.85 [0.80, 0.90] | 0.83 [0.78, 0.88] | 0.84 [0.80, 0.89] |
|  |  | Black: Other Black | 0.65 [0.59, 0.72] | 0.64 [0.58, 0.71] | 0.67 [0.60, 0.74] | 0.67 [0.60, 0.74] | 0.68 [0.61, 0.75] |
|  |  | Other: Arab | 0.46 [0.40, 0.52] | 0.45 [0.40, 0.52] | 0.49 [0.43, 0.56] | 0.50 [0.43, 0.57] | 0.52 [0.45, 0.59] |
|  |  | Other: Any other ethnic group | 0.83 [0.77, 0.89] | 0.83 [0.77, 0.90] | 0.90 [0.83, 0.97] | 0.90 [0.83, 0.98] | 0.93 [0.86, 1.01] |
|  | Ethnic group (aggregated) (Reference category: White) | Mixed/Multiple ethnic groups | 0.90 [0.87, 0.94] | 0.90 [0.87, 0.94] | 0.94 [0.90, 0.97] | 0.92 [0.88, 0.95] | 0.93 [0.90, 0.97] |
|  |  | Asian/Asian British | 0.72 [0.71, 0.74] | 0.73 [0.72, 0.75] | 0.83 [0.82, 0.85] | 0.82 [0.80, 0.84] | 0.85 [0.83, 0.87] |
|  |  | Black/African/Caribbean/Black British | 0.60 [0.58, 0.62] | 0.59 [0.57, 0.62] | 0.68 [0.66, 0.71] | 0.62 [0.60, 0.65] | 0.67 [0.65, 0.70] |
|  |  | Other ethnic group | 0.71 [0.67, 0.76] | 0.71 [0.67, 0.76] | 0.85 [0.80, 0.91] | 0.84 [0.78, 0.90] | 0.88 [0.82, 0.94] |
| Secondary analysis | Ethnic group (detailed) (Reference category: White: English/Welsh/Scottish/Northern Irish/British) | White: Irish | 0.81 [0.77, 0.85] | 0.82 [0.79, 0.86] | 0.88 [0.84, 0.93] | 0.82 [0.79, 0.86] | 0.86 [0.82, 0.90] |
|  |  | White: Gypsy or Irish Traveller | 0.99 [0.88, 1.11] | 0.94 [0.84, 1.06] | 0.95 [0.85, 1.07] | 0.95 [0.85, 1.07] | 0.96 [0.85, 1.08] |
|  |  | White: Other White | 0.63 [0.62, 0.64] | 0.66 [0.64, 0.67] | 0.74 [0.72, 0.76] | 0.74 [0.73, 0.76] | 0.78 [0.76, 0.81] |
|  |  | Mixed/multiple: White and Black Caribbean | 1.05 [1.00, 1.09] | 1.02 [0.98, 1.07] | 1.03 [0.99, 1.08] | 1.03 [0.98, 1.07] | 1.03 [0.99, 1.08] |
|  |  | Mixed/multiple: White and Black African | 0.84 [0.77, 0.91] | 0.84 [0.77, 0.91] | 0.89 [0.82, 0.97] | 0.87 [0.80, 0.95] | 0.90 [0.83, 0.98] |
|  |  | Mixed/multiple: White and Asian | 0.87 [0.82, 0.92] | 0.88 [0.84, 0.93] | 0.91 [0.86, 0.96] | 0.90 [0.85, 0.95] | 0.91 [0.86, 0.97] |
|  |  | Mixed/multiple: Other Mixed | 0.94 [0.89, 1.00] | 0.96 [0.91, 1.01] | 1.00 [0.95, 1.06] | 0.99 [0.94, 1.04] | 1.01 [0.96, 1.07] |
|  |  | Asian: Indian | 0.75 [0.73, 0.77] | 0.77 [0.75, 0.79] | 0.82 [0.80, 0.84] | 0.82 [0.80, 0.84] | 0.84 [0.82, 0.87] |
|  |  | Asian: Pakistani | 0.78 [0.76, 0.80] | 0.76 [0.74, 0.78] | 0.81 [0.78, 0.83] | 0.82 [0.79, 0.84] | 0.84 [0.82, 0.87] |
|  |  | Asian: Bangladeshi | 0.84 [0.80, 0.87] | 0.83 [0.79, 0.86] | 0.90 [0.86, 0.94] | 0.92 [0.88, 0.97] | 0.95 [0.91, 1.00] |
|  |  | Asian: Chinese | 0.43 [0.40, 0.45] | 0.46 [0.43, 0.48] | 0.50 [0.47, 0.54] | 0.50 [0.47, 0.54] | 0.53 [0.50, 0.56] |
|  |  | Asian: Other Asian | 0.81 [0.78, 0.83] | 0.83 [0.80, 0.86] | 0.92 [0.89, 0.96] | 0.93 [0.90, 0.96] | 0.98 [0.94, 1.01] |
|  |  | Black: African | 0.62 [0.60, 0.65] | 0.61 [0.59, 0.64] | 0.68 [0.66, 0.71] | 0.65 [0.63, 0.68] | 0.69 [0.67, 0.72] |
|  |  | Black: Caribbean | 1.04 [1.01, 1.08] | 1.03 [0.99, 1.06] | 1.07 [1.03, 1.10] | 1.03 [1.00, 1.07] | 1.05 [1.02, 1.09] |
|  |  | Black: Other Black | 0.89 [0.84, 0.95] | 0.88 [0.82, 0.93] | 0.92 [0.87, 0.98] | 0.92 [0.86, 0.98] | 0.94 [0.88, 1.00] |
|  |  | Other: Arab | 0.53 [0.48, 0.58] | 0.52 [0.48, 0.57] | 0.58 [0.53, 0.63] | 0.59 [0.54, 0.64] | 0.62 [0.56, 0.68] |
|  |  | Other: Any other ethnic group | 0.85 [0.81, 0.90] | 0.85 [0.81, 0.90] | 0.94 [0.89, 0.99] | 0.94 [0.89, 1.00] | 0.98 [0.93, 1.04] |
|  | Ethnic group (aggregated) (Reference category: White) | Mixed/Multiple ethnic groups | 0.98 [0.95, 1.00] | 0.98 [0.95, 1.00] | 1.02 [0.99, 1.05] | 0.99 [0.97, 1.02] | 1.01 [0.99, 1.04] |
|  |  | Asian/Asian British | 0.77 [0.76, 0.78] | 0.78 [0.77, 0.79] | 0.90 [0.89, 0.92] | 0.89 [0.87, 0.90] | 0.93 [0.91, 0.95] |
|  |  | Black/African/Caribbean/Black British | 0.81 [0.80, 0.83] | 0.80 [0.78, 0.82] | 0.92 [0.90, 0.95] | 0.84 [0.82, 0.86] | 0.91 [0.89, 0.93] |
|  |  | Other ethnic group | 0.76 [0.72, 0.79] | 0.75 [0.72, 0.79] | 0.91 [0.87, 0.96] | 0.90 [0.86, 0.94] | 0.95 [0.91, 1.00] |

### Statistical Analysis

To describe the rate of diagnosed endometriosis in NHS hospitals in England, we report crude and age-standardised rates of diagnosis per 100,000 people. Age-standardised rates were calculated as the weighted average of age-specific rates in five-year age bands. The age-specific weights represent the overall age distribution in the observed study population. To identify characteristics associated with endometriosis diagnosis, we used logistic regression models to estimate the odds ratios of being diagnosed with endometriosis in hospital at least once during the study period across different exposure groups. Individuals were followed up between 27 March 2011 and 31 December 2021. Follow-up was censored at time of death, if applicable. Models were first adjusted for age and secondly for age and health (number of pre-study HES APC admissions, excluding mentions of endometriosis). Age was included as a natural spline with boundary knots at the 1st and 99th percentile and five interior knots. The number of knots was chosen using the lowest Akaike information criterion (AIC). General health is both a potential mediator and confounder between socioeconomic factors and endometriosis; hence we first adjusted our models for age, and then further adjusted our models for health to investigate whether the relationship between our characteristics of interest and endometriosis were explained by differences in general health. When looking at general health and disability as exposures, we only adjusted for age. For ethnic group, we fitted models adjusted for main language and country of birth independently and concurrently, to investigate whether differences by ethnicity were being driven by access to healthcare services (for instance, through unfamiliarity with the NHS healthcare system and language barriers). For region, we adjusted for socioeconomic status using IMD decile and highest level of qualification as a proxy. All counts are rounded to the nearest 5 and counts of less than 10 are suppressed for disclosure reasons.

All analyses were conducted in R version 4.4 and Python version 3.10.

### Sensitivity tests

All demographic information was measured at the time of Census 2011. Some characteristics may change over time, which could introduce a bias in our estimates. We ran sensitivity analyses using just two years of follow-up (27 March 2011 to 31 December 2013) to assess the robustness of our findings across the 10-year period. In addition, we also ran sensitivity analyses when education was the exposure including only women aged 25 years and older on Census Day, as education is more likely to be stable after this age.

### Ethics and Data Availability

We obtained ethical approval for this work from the National Statistics Data Ethics Advisory Committee (NSDEC23(18)). All data relating to this work has been published as an ONS dataset. All code is available in a GitHub repository.

### Public and Patient Involvement

Women with endometriosis have reviewed this manuscript and provided feedback prior to submission.

## Results

### Main results

To estimate incidence of diagnosed endometriosis in NHS hospitals in England, we analysed endometriosis diagnoses recorded as primary diagnoses in HES APC. A total of 24,382,270 women were included in our main analysis, with 120,515 having a primary diagnosis of endometriosis [Supplementary Table 1]. The crude rates of endometriosis diagnosis per 100,000 people were highest among women aged 25 to 29 (1,217.48) and 30 to 34 (1,210.61) years at the time of Census 2021 [Table 2]. The average age at diagnosis was 35 years (IQR: 27-43 years) [Supplementary Table 5], and 10.7% of hospital admissions with endometriosis recorded as the primary diagnosis were classified as emergencies [Supplementary Table 6].

**Table 5:**
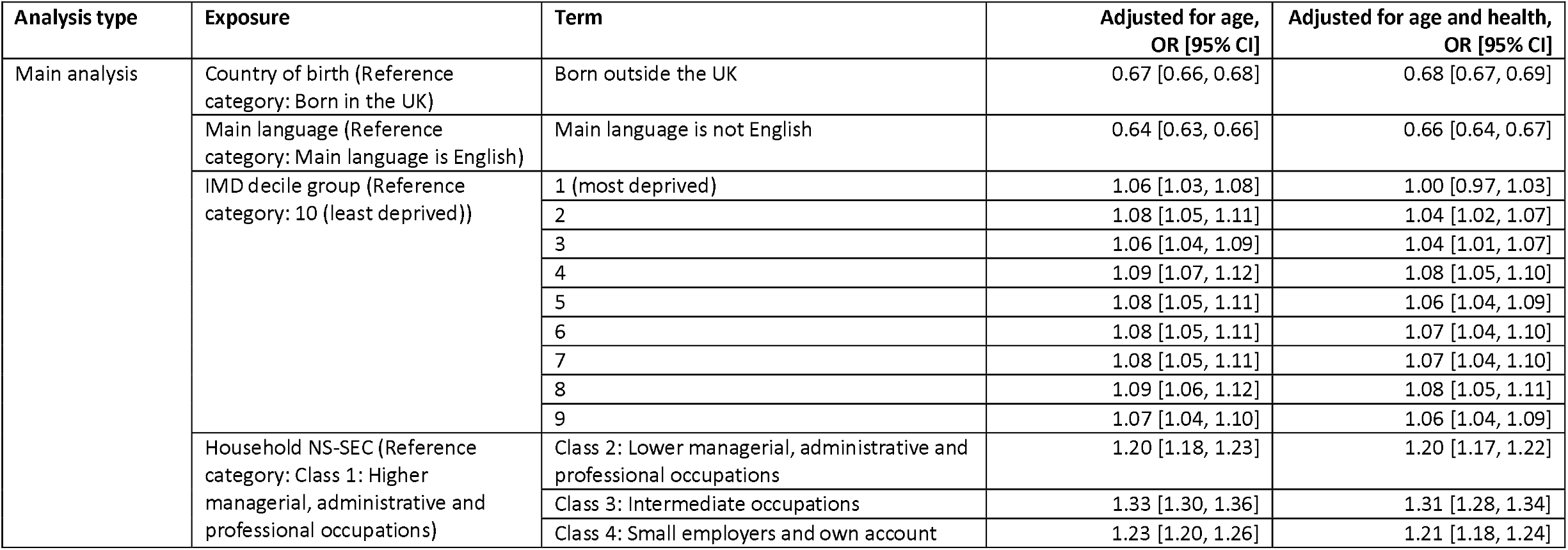
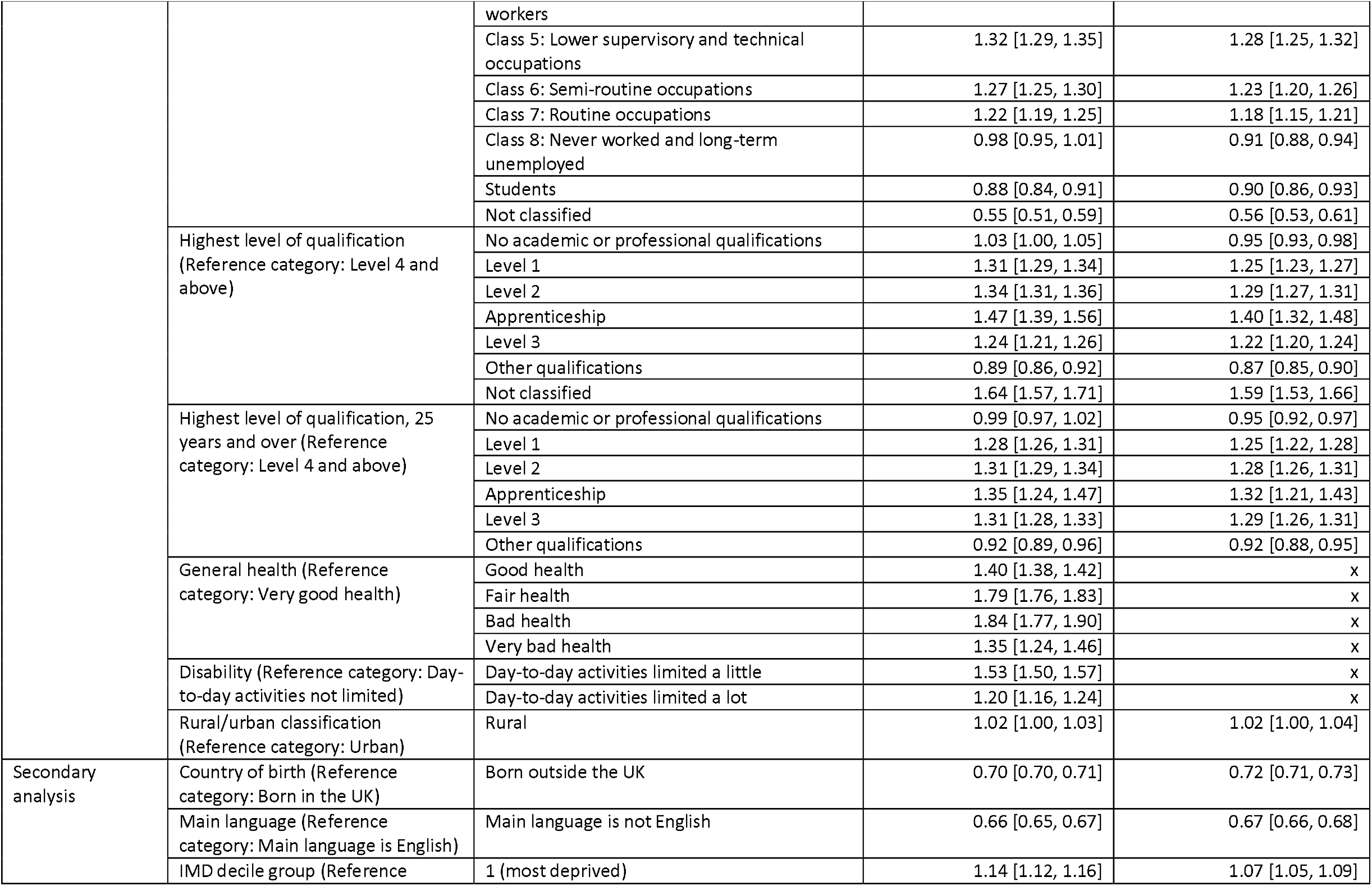
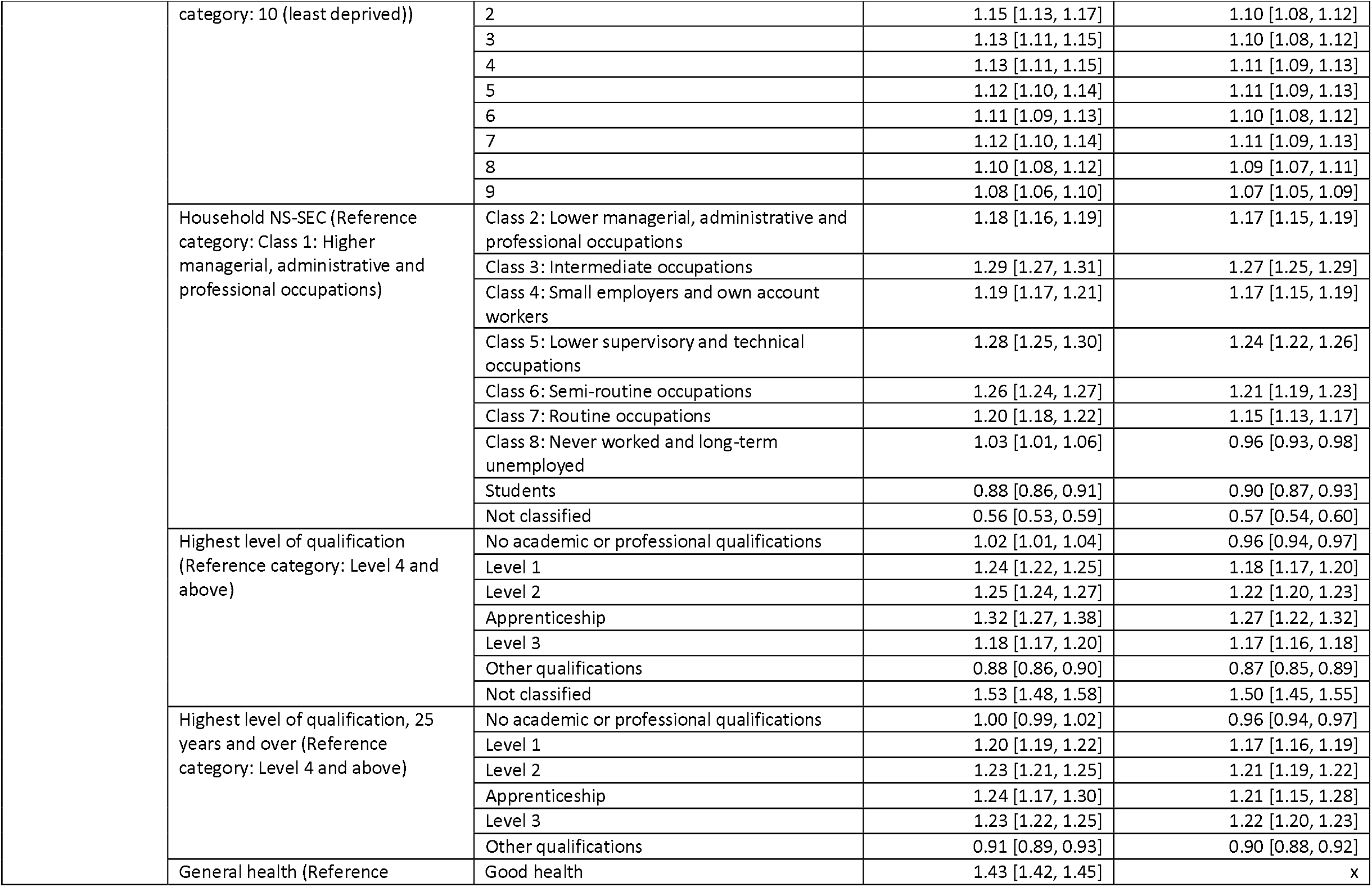
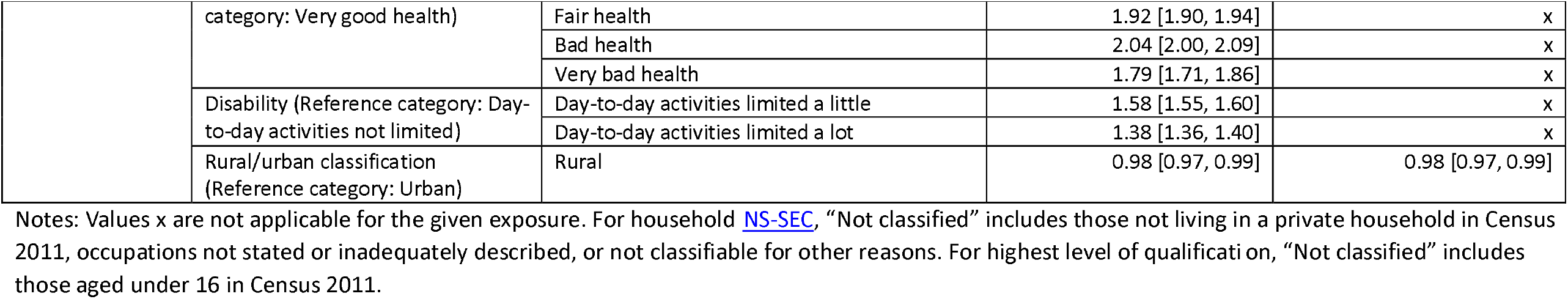
Odds ratios for endometriosis diagnosis and 95% confidence intervals by country of birth, main language, IMD decile group, household NS-SEC, highest level of qualification, general health, disability and rural/urban classification.

Age-standardised rates of diagnosed endometriosis were highest in the White (518.66 per 100,000 persons; 95% confidence interval (CI): 515.53-521.80) and Mixed/Multiple (480.88 per 100,000 persons; 95%CI: 460.51-501.24) ethnic groups [Table 2]. However, the Mixed/Multiple ethnic groups had a significantly lower odds of diagnosis compared with the White group in both the age-adjusted model (Odds Ratio (OR): 0.90; 95%CI: 0.87-0.94) and the age- and health-adjusted model (OR: 0.90; 95%CI: 0.87-0.94) [Table 4]. Compared with the White ethnic group, the largest differences in odds of diagnosis were observed for the Black/African/Caribbean/Black British (OR: 0.59; 95%CI: 0.57-0.62), the Other ethnic group (OR:0.71; 95%CI:0.67-0.76), and the Asian/Asian British group (OR: 0.73; 95%CI:0.72-0.75) ethnic groups. When further adjusting for country of birth and main language, the odds of diagnosis for the White ethnic group remained significantly higher than all other ethnic groups.

Using the more granular breakdown for ethnic group, compared with the White British (White English/Welsh/Scottish/Northern Irish/British) group, the Chinese (Asian/Asian British) (OR: 0.40; 95%CI: 0.37-0.44), African (Black/African/Caribbean/Black British) (OR: 0.43; 95%CI: 0.41-0.46) and Arab (Other ethnic group) (OR: 0.45; 95%CI: 0.40-0.52) ethnic groups had the lowest odds of diagnosis in the age- and health-adjusted model.

Odds of diagnosis were lower for women born outside the UK (compared with those born in the UK; OR: 0.68; 95%CI: 0.67-0.69), and for women whose main language was not English (compared with those with English as their first language; OR: 0.66; 95%CI:0.64-0.67) [Table 5].

Odds of a diagnosis were lowest for women living in the most deprived and least deprived areas of the country [Table 5]. Analysis of household NS-SEC showed the odds were lowest for women in households with the highest and lowest socio-economic classifications. When looking at education level, the odds ratios were lowest for women with Other (foreign or vocational) qualifications (OR:0.87; 95%CI:0.85-0.90) or no qualifications (OR:0.95; 95%CI:0.93-0.98), compared with Level 4 and above qualifications (e.g., degree level).

Odds of diagnosis were highest for women self-reporting to be in bad health (OR: 1.84; 95%CI: 1.77-1.90) or fair health (OR: 1.79; 95%CI: 1.76-1.83), compared with those in very good health. Disabled women reporting to be limited a little (OR: 1.53; 95%CI: 1.50-1.57) or limited a lot (OR: 1.20; 95%CI: 1.16-1.24) in their day-to-day activities had significantly higher odds of diagnosis, compared with non-disabled women.

Compared with London, the odds of a diagnosis were higher for all other regions of the UK [Supplementary Table 9]. Adjusting for IMD and education level had minimal impact on these results overall, except for the North East where the 95% CI included the null after adjustment (OR:1.02;95%CI:0.99-1.06). Broadly, we did not see clear differences in age-standardised rates of diagnosed endometriosis by UTLA [Figure 3, Supplementary Table 10].

### Secondary results

To estimate the underlying rate of diagnosed endometriosis in NHS hospitals in England, we analysed endometriosis diagnoses recorded as either primary or secondary diagnoses in HES APC. 24,560,795 women were included in our secondary analysis, of which 262,065 had a primary or secondary diagnosis of endometriosis in hospital. This is equivalent to approximately 2% of women aged 15 to 49 years [Table 1, Figure 1]. Crude rates of diagnosed endometriosis diagnosis in hospital per 100,000 people were highest among women aged 30 to 34 (2,307.61) and 35 to 39 (2,492.44) years at the time of the 2011 Census [Table 2]. The average age at time of diagnosis was 39.8 years (IQR: 30-47 years), five years older than in the main analysis [Supplementary Table 5].

**Figure 1:**
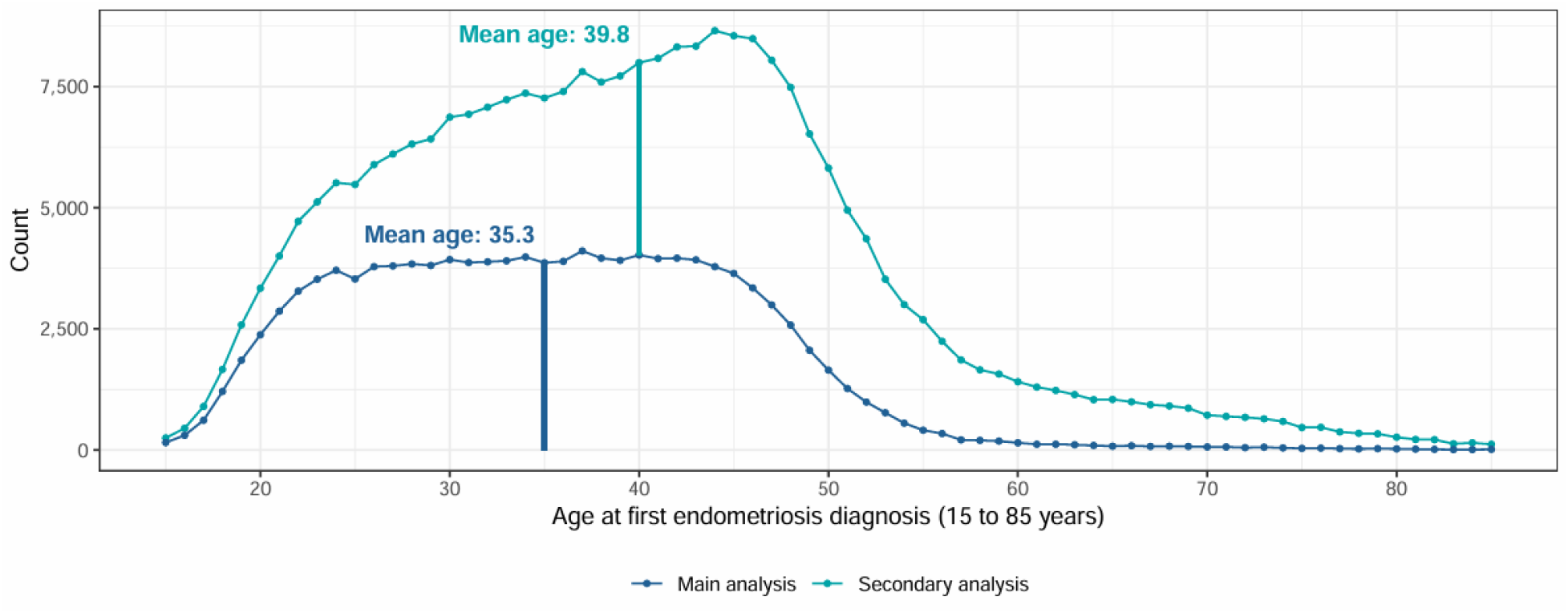
Age distribution of study population at time of first endometriosis diagnosis in study period. The number of diagnoses at each year of age for first endometriosis diagnosis in hospital in our main analysis population (blue) and secondary analysis population (teal). The main analysis population includes primary endometriosis diagnoses only, and the secondary analysis population includes both primary and secondary endometriosis diagnoses. The vertical bar marks the mean age at time of diagnosis for each cohort.

**Figure 2:**
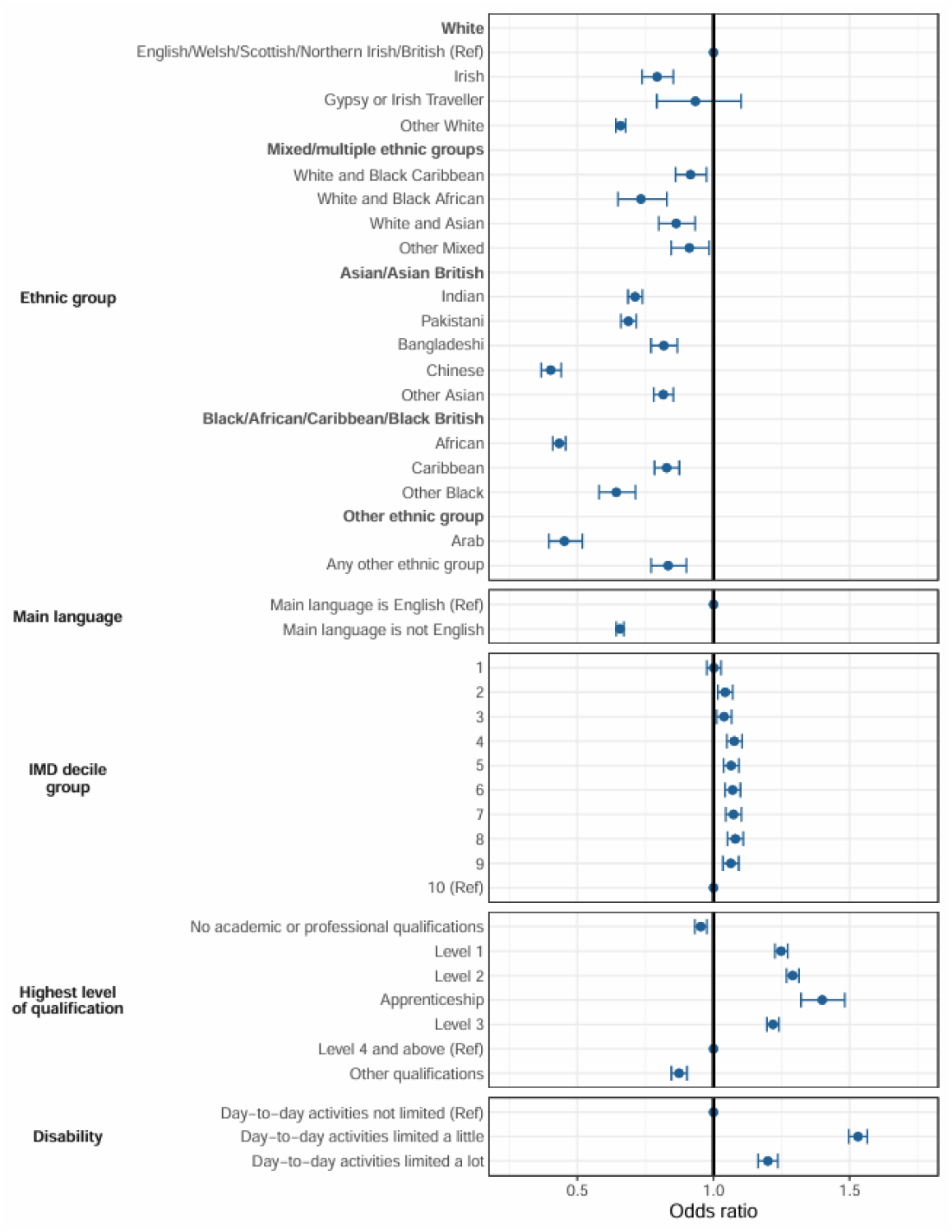
Odds of receiving a primary endometriosis diagnosis by sociodemographic characteristics. The odds ratio of receiving a primary endometriosis diagnosis by ethnic group, main language, IMD decile group, highest level of qualification and disability. The bold vertical line indicates an odds ratio of 1. Point estimates are presented with 95% confidence intervals. All models account for age and health (except for the disability model, which is only age-adjusted).

**Figure 3:**
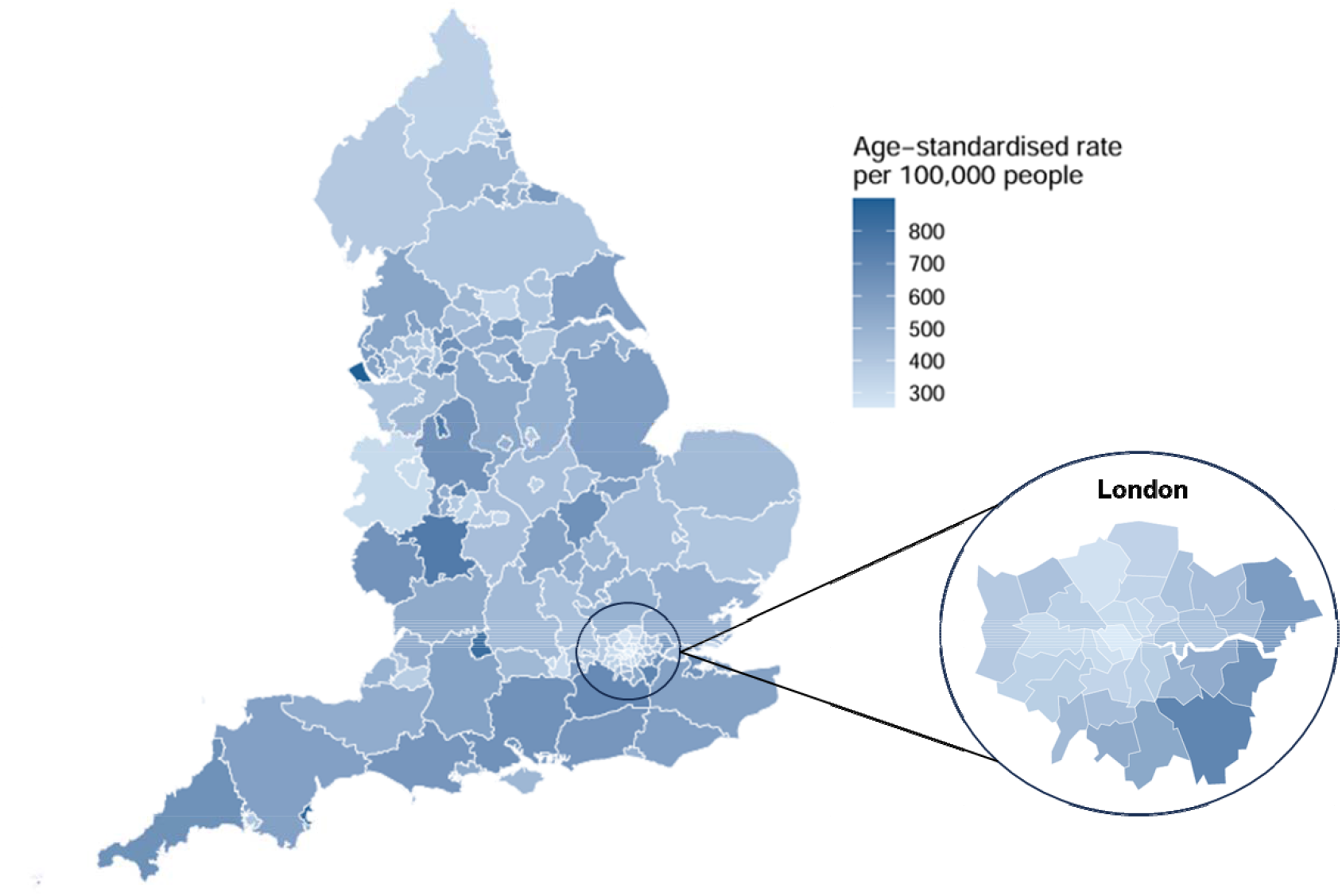
Age-standardised rates of primary endometriosis diagnosis, per 100,000 people by upper tier local authority (UTLA) Age-standardised rates of primary endometriosis diagnosis per 100,000 people shown by UTLA, with darker colours illustrating higher rates. The regions in Greater London have been shown in a second pop-out.

The most frequent primary diagnosis type by ICD-10 code in our secondary cohort was endometriosis (n = 116,835, 44.6%), with the next most prevalent being leiomyoma of uterus (n = 21,845, 8.3%). The top ten primary diagnosis codes included a range of other gynaecological presentations such as abdominal and pelvic pain, non-inflammatory disorders of ovary, fallopian tube and broad ligament, as well as excessive frequent and irregular menstruation [Supplementary Table 11]. 18.7% of admissions were emergencies, with the majority (81.3%) being classified as non-emergency presentations [Supplementary Table 6].

Using the aggregated ethnicity breakdown, the odds ratio comparing diagnosis in the Black/African/Caribbean/Black British ethnic group to the White group was higher in the secondary analysis (OR:0.91; 95%CI:0.89-0.93) compared with the main analysis (OR:0.67; 95%CI:0.65-0.70) in the fully adjusted model [Table 4]. Looking at the more granular breakdown for ethnic group, compared with the White British (White English/Welsh/Scottish/Northern Irish/British) group, the odds of diagnosis were significantly lower for the Black African (OR:0.69; 95%CI:0.67-0.72) group, showed no evidence of a significant difference for the Other Black (OR:0.94; 95%CI:0.88-1.00) group, and were significantly higher for the Black Caribbean (OR:1.05; 95%CI:1.02-1.09) group.

Furthermore, the odds ratios for women self-reporting to be in good, fair, bad or very bad health, compared with those in very good health, were higher compared with the main analysis [Table 5]. Similarly, the odds ratios of women self-reporting to be limited a little or a lot in their day-to-day activities, compared with those not limited, were higher in the secondary analysis. When looking at region, the odds ratios were generally similar, though Yorkshire and the Humber and the South East showed the most prominent decreases in odds ratios compared with the main analysis [Supplementary Table 9]. In general, for all other sociodemographic characteristics we saw the same pattern of results as in the main analysis [Tables 3-5, Supplementary Table 10].

### Sensitivity results

To assess the robustness of our findings over a 10-year follow-up, we repeated the analysis using just a two-year follow-up. 32,855 women had a primary endometriosis diagnosis in our main sensitivity analysis [Supplementary Table 1], and 71,345 women had a primary or secondary endometriosis diagnosis in our secondary sensitivity analysis. For both groups, the sensitivity results were consistent with the findings from the 10-year follow-up. The odds ratios for women reporting to be in good, fair, bad or very bad health compared with those in very good health were all further from the null in the two-year follow up compared with the 10-year follow up [Table 5 and Supplementary Table 12]. Similarly, the odds ratios for women reporting to be limited a little or a lot in their day-to-day activities compared with those not limited were higher in the two-year follow up. Furthermore, we repeated our analysis of the highest level of educational qualification exposure restricting to women who were aged 25 years and over on Census Day, since education level is more stable from this age onwards. Results followed a similar pattern to the analysis of women of any age [Table 5 and Supplementary Table 12].

## Discussion

O results show that the odds of receiving an endometriosis diagnosis in England vary significantly by sociodemographic characteristics and reflect differences in both the likelihood of having endometriosis as well as the likelihood of receiving a diagnosis in an NHS hospital.

The average age at first endometriosis diagnosis in hospital, where endometriosis was the primary diagnosis, was 35 years. Given that symptoms of this disease can start in puberty, these findings support previous work suggesting that women are facing barriers and delays in receiving an endometriosis diagnosis (7,8). In an Australian cohort study of over 10,000 women, 1 in 9 self-reported suspected or confirmed endometriosis by age 44, with most being diagnosed during their early thirties (21). Since 2020, NHS gynaecology waiting lists in England have grown faster than any other elective speciality in percentage terms (22). If left untreated, endometriosis may progress and further negatively impact on quality of life. Future work should explore inequalities in delays in diagnosis across sociodemographic groups.

We estimated the underlying rate of diagnosed endometriosis in NHS hospitals to be 2% of reproductive age women in England based on primary and secondary diagnoses of endometriosis (16). Previous estimates of prevalence of endometriosis range from 2% to 10% in the general female population and up to 50% in women with infertility (17). Endometriosis UK and the World Health Organization (WHO) estimate that 1 in 10 women of reproductive age have endometriosis (1,18), and pooled estimates of European studies suggest a prevalence rate of 1.4% in the general population (19). Our estimate of 2% of reproductive age women is lower than some previous studies. However, our estimate only reflects the number of women who have been diagnosed with endometriosis in an NHS hospital in England between 2011 and 2021. Moreover, our analysis only includes women whose Census 2011 response could be linked to an NHS number; it does not capture all women who have had a diagnosis in hospital. For our main analysis, our sample includes approximately 85% of all women who received an endometriosis diagnosis in hospital during this period [Supplementary Table 1]. It is important to note that prevalence of endometriosis diagnosis in hospital does not reflect the true prevalence of endometriosis in the population. Some women with endometriosis will be undiagnosed. In the US, for instance, six out of ten cases go undiagnosed (20). Some women may be receiving treatment for endometriosis in primary care but have not been referred to hospital for a definitive diagnosis and would therefore not be identified as having an endometriosis diagnosis in this study. Hospital admissions describing symptoms of endometriosis, but not specifically mentioning a diagnosis of endometriosis (ICD-10 codes N80.0-N80.9), may include women with endometriosis who are undiagnosed or misdiagnosed, but would not be defined as having an endometriosis diagnosis in this study.

Previous survey data has shown that over half of women who had symptoms of endometriosis, without a confirmed diagnosis, attended A&E at least once due to their symptoms (21). Although a small proportion of our cases were identified as emergency admissions (10.7%) it is still an important finding that warrants additional exploration. The cost of repeated presentations at A&E centres, as well as emergency treatment costs, in contrast to elective care, should be considered when making recommendations for diagnostic care for women with endometriosis (22).

Finding significantly lower odds of being diagnosed with endometriosis for Chinese, Black African or Arab ethnic groups compared with the White English/Welsh/Scottish/Northern Irish/British group is broadly consistent with existing literature from elsewhere in the world, and smaller UK-based studies (23). Many of these findings need to be interpreted with caution due to limited quality literature exploring this topic and selection bias of White women in research studies. Historically, endometriosis has been thought to be a disease most prevalent in White women (20), but our results demonstrate the odds of diagnosis are as high in the Mixed/multiple ethnic groups. Differences in odds of endometriosis diagnosis between ethnic groups are likely to be explained by social and structural issues around access to healthcare and systemic racism (23).

In our study, women living in the most deprived and least deprived areas had lower odds of diagnosis, compared with those living in other areas. This might be explained by those living in the most deprived areas facing barriers to seeking treatment (24) including longer NHS waiting times (25), and those living in the least deprived areas being more likely to use private healthcare (26). Following a similar trend, we found lower odds of diagnosis for women in households with the highest and lowest socio-economic classifications. Of all educational groups, women in the Other qualifications group had the lowest odds of diagnosis. This group includes women with foreign qualifications, and therefore may include women who received an endometriosis diagnosis outside England not captured by the HES data.

Women self-reporting to be in worse than very good health or disabled had higher odds of diagnosis compared with those in very good health or not disabled, respectively. The odds ratios were higher in the secondary analysis including both primary and secondary diagnoses of endometriosis compared with the main analysis including primary diagnoses only. However, the results of the secondary analysis are likely to also reflect differences in underlying health, because secondary diagnoses require women to have been hospitalised for another reason.

Our sensitivity analyses, using two years of follow-up, showed similar findings to the 10-year follow up. One difference was the odds for women reporting to be limited in their day-to-day activities, compared with those not limited, which were higher in the two-year follow up. These findings indicate that symptoms of endometriosis could be contributing to self-reported health and disability status, highlighting how debilitating endometriosis symptoms can be on overall health.

A strength of our study is the use of population level data for England, making it the most comprehensive study into the characteristics of women with an endometriosis diagnosis in England to date. Furthermore, the linkage to Census data provides detailed information on socioeconomic characteristics. A key limitation of our work is we only capture women who have an endometriosis diagnosis in an NHS hospital in England which, according to the National Institute for Health and Care Excellence (NICE) guidelines, will capture women who have been referred to gynaecology if initial symptom management in primary care is not effective (9). A laparoscopy should be used to diagnose endometriosis even if an ultrasound or MRI has come back normal. Subsequent research should explore diagnosis of endometriosis using primary care data, in addition to secondary care data, to get a more complete understanding of the burden of disease, as well as capturing symptoms related to diagnosis since many women remain undiagnosed.

A limitation of this study is that death could have occurred for a woman with endometriosis prior to being diagnosed with endometriosis in hospital. However, mortality rates in the 25 to 54 age group are low compared with other age groups (27).

Clinically, endometriosis is typically classified into four stages, from minimal to severe, taking into account location and depth of disease in relation to other pelvic structures (28). We are limited to assessing endometriosis diagnosis as a binary outcome based on our data but implore researchers to evaluate sociodemographic characteristics associated with disease severity in subsequent work. Importantly, treatment and hospital diagnosis are based on severity of symptoms, not clinical stage, which often does not correspond to severity of symptoms and cannot be determined without laparoscopy. Many chronic conditions that affect health and disability will only be captured in primary care data, which we did not have access to for this work, so rely on an indicator from secondary care. Finally, using regional information on place of residence from Census (i.e., at the start of the study period) and HES (i.e., at time of diagnosis) is limited as we are not able to evaluate regional differences in diagnosis or procedures. Across the UK, there are 79 specialist endometriosis centres accredited by the British Society of Gynaecology and Endoscopy (29), and subsequent work should look at differences here.

To conclude, our study provides the most comprehensive analysis of the characteristics of women with an endometriosis diagnosis in an NHS hospital in England to date. We utilised a decade’s worth of endometriosis diagnoses taking place in NHS hospitals and linked to Census data to provide a granular understanding of the groups who have the highest odds of receiving an endometriosis diagnosis. This work provides important information to gynaecologists, clinicians and other allied health professionals, as well as policy makers to illustrate the underlying rates of diagnosed endometriosis in NHS hospitals and the groups most affected.

## Data Availability

All data relating to this work has been published as an ONS dataset.

https://www.ons.gov.uk/peoplepopulationandcommunity/healthandsocialcare/healthinequalities/datasets/characteristicsofwomenwithanendometriosisdiagnosisinengland

## Contributions of authors

ILW, VN and DA conceptualised and designed the study. ILW, EC and HB prepared the study data. HB performed the statistical analysis, which was quality checked by ILW. All authors contributed to interpretation of the findings. ILW and HB wrote the original draft. All authors contributed to review and editing of the manuscript and approved the final manuscript. ILW is the guarantor. The corresponding author attests that all listed authors meet authorship criteria and that no others meeting the criteria have been omitted.

## Acknowledgements

We would like to thank Endometriosis UK and the Department of Health and Social Care (DHSC) for supporting our research and public engagement. We would also like to thank Emily Williams for supporting with a literature review early in the study development.

## Funding

This study received funding from His Majesties Treasury’s Labour Market Evaluation Fund (30). GCS is supported by the Medical Research Council (MR/Z504634).

## Conflicts of interest

The authors have no conflicts to declare.

## Notes

### Competing Interest Statement

The authors have declared no competing interest.

### Funding Statement

This study received funding from His Majesties Treasurys Labour Market Evaluation Fund.

### Author Declarations

Ethical approval was obtained from the National Statistician's Data Ethics Advisory Committee.

### Summary of Updates

Revised manuscript following peer-review. The primary and secondary cohorts have been re-ordered for clarity.

