## Supplement for "Characteristics of women diagnosed with endometriosis in England: 2011-2021"

### Supplementary Tables

**Supplementary Table 1: Sample flow**

| **Sample** | **Count, Individuals** | **Count, Primary endometriosis diagnoses (Main analysis)** | **Count, Primary and secondary endometriosis diagnoses (Secondary analysis)** |
| --- | --- | --- | --- |
| All people enumerated in Census 2011 | 56,963,120 | x | x |
| Usual resident in England in Census 2011 | 56,075,910 | x | x |
| Living in England in Census 2011 | 53,012,455 | x | x |
| Linked to the NHS 2011-2013 Patient Registers | 47,454,185 | x | x |
| Alive on Census Day (27 March 2011) | 47,454,035 | x | x |
| Episode-level Hospital Episode Statistics (HES) Admitted Patient Care (APC) episodes which started and ended between 27 March 2011 and 31 December 2021 | 216,385,210 | 201,535 | 550,010 |
| Person-level Hospital Episode Statistics (HES) Admitted Patient Care (APC) episodes which started and ended between 27 March 2011 and 31 December 2021 | 42,146,010 | 151,485 | 308,610 |
| Census 2011 linked to person-level Hospital Episode Statistics (HES) Admitted Patient Care (APC) episodes | 47,454,035 | 128,465 | 262,560 |
| Self-reported as female in Census 2011 | 24,560,795 | 128,215 | 262,065 |
| Main analysis: No primary or secondary endometriosis diagnosis between 1 April 2009 and 26 March 2011, and either no endometriosis diagnosis between 27 March 2011 and 31 December 2021 or a primary endometriosis diagnosis between 27 March 2011 and 31 December 2021 | 24,382,270 | 120,515 | x |
| Main sensitivity analysis: No primary or secondary endometriosis diagnosis between 1 April 2009 and 26 March 2011, and either no endometriosis diagnosis between 27 March 2011 and 31 December 2013 or a primary endometriosis diagnosis between 27 March 2011 and 31 December 2013 | 24,478,655 | 32,855 | x |
| Secondary analysis: Either no endometriosis diagnosis between 27 March 2011 and 31 December 2021 or a primary or secondary endometriosis diagnosis between 27 March 2011 and 31 December 2021 | 24,560,795 | x | 262,065 |
| Secondary sensitivity analysis: Either no endometriosis diagnosis between 27 March 2011 and 31 December 2013 or a primary or secondary endometriosis diagnosis between 27 March 2011 and 31 December 2013 | 24,560,795 | x | 71,345 |

Note: Values x are not applicable for the given sample.

**Supplementary Table 2: List of ICD-10 codes used to define endometriosis**

| **ICD-10 code** | **Description** |
| --- | --- |
| N80 | Endometriosis |
| N80.0 | Endometriosis of uterus |
| N80.1 | Endometriosis of ovary |
| N80.2 | Endometriosis of fallopian tube |
| N80.3 | Endometriosis of pelvic peritoneum |
| N80.4 | Endometriosis of rectovaginal septum and vagina |
| N80.5 | Endometriosis of intestine |
| N80.6 | Endometriosis in cutaneous scar |
| N80.8 | Other endometriosis |
| N80.9 | Endometriosis, unspecified |

**Supplementary Table 3: Population characteristics**

| **Characteristic** | **Data source** | **Classification** |
| --- | --- | --- |
| Age on Census Day (five-year bands) | Census 2011 | 0 to 9 years  10 to 14 years  15 to 19 years  20 to 24 years  25 to 29 years  30 to 34 years  35 to 39 years*  40 to 44 years  45 to 49 years  50 to 54 years  55 to 59 years  60 to 64 years  65 to 69 years  70 to 74 years  75 to 79 years  80 years and over |
| General health | Census 2011 | Very good health*  Good health  Fair health  Bad health  Very bad health |
| Disability status | Census 2011 | Day-to-day activities not limited*  Day-to-day activities limited a little  Day-to-day activities limited a lot |
| Ethnicity (detailed) | Census 2011 | White: English/Welsh/Scottish/Northern Irish/British*  White: Irish  White: Gypsy or Irish Traveller  White: Other White  Mixed/multiple: White and Black Caribbean  Mixed/multiple: White and Black African  Mixed/multiple: White and Asian  Mixed/multiple: Other Mixed  Asian: Indian  Asian: Pakistani  Asian: Bangladeshi  Asian: Chinese  Asian: Other Asian  Black: African  Black: Caribbean  Black: Other Black  Other: Arab  Other: Any other ethnic group |
| Ethnicity (aggregated) | Census 2011 | White*  Mixed/multiple ethnic groups  Asian/Asian British  Black/African/Caribbean/Black British  Other ethnic group |
| Country of birth | Census 2011 | Born in the UK*  Born outside the UK |
| Household NS-SEC | Census 2011 | Class 1: Higher managerial, administrative and professional occupations*  Class 2: Lower managerial, administrative and professional occupations  Class 3: Intermediate occupations  Class 4: Small employers and own account workers  Class 5: Lower supervisory and technical occupations  Class 6: Semi-routine occupations  Class 7: Routine occupations  Class 8: Never worked and long-term unemployed  Students  Not classified |
| Highest qualification | Census 2011 | No academic or professional qualifications  Level 1  Level 2  Apprenticeship  Level 3  Level 4 and above*  Other qualifications  Not classified |
| Highest qualification,  25 years and over | Census 2011 | No academic or professional qualifications  Level 1  Level 2  Apprenticeship  Level 3  Level 4 and above*  Other qualifications  Aged 0 to 24 years on Census Day |
| Rural/urban classification | Census 2011 | Urban*  Rural |
| Main language | Census 2011 | Main language is English*  Main language is not English |
| IMD decile group | Census 2011 | 1 (most deprived)  2  3  4  5  6  7  8  9  10 (least deprived)* |
| Region | Census 2011 | North East  North West  Yorkshire and the Humber  East Midlands  West Midlands  East of England  London*  South East  South West  Outside England or unknown |
| Upper tier local authority (UTLA) | Census 2011 | Barking and Dagenham  Barnet  Barnsley  Bath and North East Somerset  Bedford  Bexley  Birmingham  Blackburn with Darwen  Blackpool  Bolton  Bournemouth, Christchurch and Poole  Bracknell Forest  Bradford  Brent  Brighton and Hove  Bristol, City of  Bromley  Buckinghamshire  Bury  Calderdale  Cambridgeshire  Camden  Central Bedfordshire  Cheshire East  Cheshire West and Chester  City of London and Hackney*  Cornwall and Isles of Scilly  County Durham  Coventry  Croydon  Cumbria  Darlington  Derby  Derbyshire  Devon  Doncaster  Dorset  Dudley  Ealing  East Riding of Yorkshire  East Sussex  Enfield  Essex  Gateshead  Gloucestershire  Greenwich  Halton  Hammersmith and Fulham  Hampshire  Haringey  Harrow  Hartlepool  Havering  Herefordshire, County of  Hertfordshire  Hillingdon  Hounslow  Isle of Wight  Islington  Kensington and Chelsea  Kent  Kingston upon Hull, City of  Kingston upon Thames  Kirklees  Knowsley  Lambeth  Lancashire  Leeds  Leicester  Leicestershire  Lewisham  Lincolnshire  Liverpool  Luton  Manchester  Medway  Merton  Middlesbrough  Milton Keynes  Newcastle upon Tyne  Newham  Norfolk  North East Lincolnshire  North Lincolnshire  North Northamptonshire  North Somerset  North Tyneside  North Yorkshire  Northumberland  Nottingham  Nottinghamshire  Oldham  Oxfordshire  Peterborough  Plymouth  Portsmouth  Reading  Redbridge  Redcar and Cleveland  Richmond upon Thames  Rochdale  Rotherham  Rutland  Salford  Sandwell  Sefton  Sheffield  Shropshire  Slough  Solihull  Somerset  South Gloucestershire  South Tyneside  Southampton  Southend-on-Sea  Southwark  St. Helens  Staffordshire  Stockport  Stockton-on-Tees  Stoke-on-Trent  Suffolk  Sunderland  Surrey  Sutton  Swindon  Tameside  Telford and Wrekin  Thurrock  Torbay  Tower Hamlets  Trafford  Wakefield  Walsall  Waltham Forest  Wandsworth  Warrington  Warwickshire  West Berkshire  West Northamptonshire  West Sussex  Westminster  Wigan  Wiltshire  Windsor and Maidenhead  Wirral  Wokingham  Wolverhampton  Worcestershire  York |
| Number of HES APC episodes pre-study, excluding endometriosis diagnoses | HES APC | 0 episodes  1 to 3 episodes  4 to 6 episodes  7 to 9 episodes  10 to 14 episodes  15 or more episodes |

Note: Classifications marked with an asterisk (*) were used as the reference category in the modelling. All characteristics except “Number of HES APC episodes pre-study, excluding endometriosis diagnoses” were self-reported from Census 2011. The 80+ group has been aggregated for disclosure control reasons.

**Supplementary Table 4: Method of hospital admission in HES APC**

| **Method of hospital admission in HES APC** | **Description** |
| --- | --- |
| Emergency admission | 21 = Emergency Care Department or acute or emergency dental service  22 = General Practitioner: after a request for immediate admission has been made direct to a Hospital Provider, i.e. not through a Bed bureau, by a GP or deputy  23 = Bed bureau  24 = Consultant Clinic, of this or another Health Care Provider  25 = Admission via Mental Health Crisis Resolution Team (available from 2013/14)  2A = Emergency Care Department of another provider where the patient had not been admitted (available from 2013/14)  2B = Transfer of an admitted patient from another Hospital Provider in an emergency (available from 2013/14)  2C = Baby born at home as intended (available from 2013/14)  2D = Other emergency admission (available from 2013/14)  28 = Other means, examples are:  - admitted from the Accident and Emergency Department of another provider where they had not been admitted  - transfer of an admitted PATIENT from another Hospital Provider in an emergency  - baby born at home as intended (retired on 2022-23) |
| Non-emergency admission | 11 = Waiting list. A patient admitted electively from a waiting list having been given no date of admission at a time a decision was made to admit  12 = Booked. A Patient admitted having been given a date at the time the decision to admit was made, determined mainly on the grounds of resource availability  13 = Planned. A Patient admitted, having been given a date or approximate date at the time that the decision to admit was made. This is usually part of a planned sequence of clinical care determined mainly on social or clinical criteria (e.g. check cystoscopy)". A planned admission is one where the date of admission is determined by the needs of the treatment, rather than by the availability of resources.  31 = Admitted ante-partum  32 = Admitted post-partum  82 = The birth of a baby in this Health Care Provider  83 = Baby born outside the Health Care Provider except when born at home as intended.  81 = Transfer of any admitted patient from other Hospital Provider other than in an emergency |
| Not applicable or unknown method of admission | 98 = Not applicable  99 = Method of admission not known |

Source: [NHS Digital Hospital Episode Statistics Data Dictionary](https://digital.nhs.uk/data-and-information/data-tools-and-services/data-services/hospital-episode-statistics/hospital-episode-statistics-data-dictionary)

**Supplementary Table 5: Age at first endometriosis diagnosis during the study period**

| **Age at first endometriosis diagnosis during the study period** | **Count,**  **Main analysis** | **Count,**  **Secondary analysis** |
| --- | --- | --- |
| Total | 120,515 | 262,065 |
| Mean | 35.3 | 39.8 |
| 25th percentile | 27 | 30 |
| 50th percentile (Median) | 35 | 39 |
| 75th percentile | 43 | 47 |
| 0 to 14 years | 30 | 55 |
| 15 to 19 years | 1,065 | 1,550 |
| 20 to 24 years | 3,860 | 6,015 |
| 25 to 29 years | 4,905 | 8,605 |
| 30 to 34 years | 5,375 | 10,185 |
| 35 to 39 years | 5,745 | 11,220 |
| 40 to 44 years | 5,910 | 12,795 |
| 45 to 49 years | 4,105 | 10,750 |
| 50 to 54 years | 1,210 | 4,505 |
| 55 to 59 years | 265 | 1,885 |
| 60 to 64 years | 145 | 1,325 |
| 65 to 69 years | 100 | 1,105 |
| 70 to 74 years | 60 | 660 |
| 75 to 79 years | 45 | 405 |
| 80 years and over | 35 | 285 |

**Supplementary Table 6: Method of hospital admission at first endometriosis diagnosis during the study period**

| **Method of hospital admission** | **Percentage, Main analysis** | **Percentage, Secondary analysis** |
| --- | --- | --- |
| Emergency admissions of endometriosis | 10.7 | 18.7 |
| Non-emergency admissions of endometriosis | 89.3 | 81.3 |
| Not applicable or not known | <0.1 | <0.1 |

**Supplementary Table 7: Characteristics of the study population, number of HES APC episodes pre-study and upper tier local authority (UTLA)**

| **Category** | **Subcategory** | **Main analysis** | | **Secondary analysis** | |
| --- | --- | --- | --- | --- | --- |
|  |  | **Count (%), with an endometriosis diagnosis** | **Count (%), no endometriosis diagnosis** | **Count (%), with an endometriosis diagnosis** | **Count (%), no endometriosis diagnosis** |
| Number of HES APC episodes pre-study, excluding endometriosis diagnoses | 0 episodes | 77,945 (64.68%) | 17,611,970 (72.59%) | 167,025 (63.73%) | 17,630,710 (72.56%) |
|  | 1 to 3 episodes | 36,235 (30.07%) | 5,800,505 (23.91%) | 80,010 (30.53%) | 5,816,005 (23.94%) |
|  | 4 to 6 episodes | 4,460 (3.70%) | 577,980 (2.38%) | 10,350 (3.95%) | 579,825 (2.39%) |
|  | 7 to 9 episodes | 1,110 (0.92%) | 145,445 (0.60%) | 2,655 (1.01%) | 145,940 (0.60%) |
|  | 10 to 14 episodes | 530 (0.44%) | 73,360 (0.30%) | 1,295 (0.49%) | 73,590 (0.30%) |
|  | 15 or more episodes | 235 (0.19%) | 52,495 (0.22%) | 730 (0.28%) | 52,660 (0.22%) |
| Upper tier local authority (UTLA) | Barking and Dagenham | 435 (0.36%) | 82,880 (0.34%) | 920 (0.35%) | 82,990 (0.34%) |
|  | Barnet | 485 (0.40%) | 157,520 (0.65%) | 1,315 (0.50%) | 157,690 (0.65%) |
|  | Barnsley | 560 (0.46%) | 108,910 (0.45%) | 1,300 (0.50%) | 109,115 (0.45%) |
|  | Bath and North East Somerset | 305 (0.25%) | 82,480 (0.34%) | 665 (0.25%) | 82,575 (0.34%) |
|  | Bedford | 350 (0.29%) | 73,950 (0.30%) | 685 (0.26%) | 74,030 (0.30%) |
|  | Bexley | 700 (0.58%) | 108,985 (0.45%) | 1,445 (0.55%) | 109,180 (0.45%) |
|  | Birmingham | 1,880 (1.56%) | 469,875 (1.94%) | 4,930 (1.88%) | 470,470 (1.94%) |
|  | Blackburn with Darwen | 405 (0.34%) | 66,395 (0.27%) | 850 (0.32%) | 66,560 (0.27%) |
|  | Blackpool | 360 (0.30%) | 66,040 (0.27%) | 800 (0.31%) | 66,110 (0.27%) |
|  | Bolton | 530 (0.44%) | 126,790 (0.52%) | 1,270 (0.48%) | 126,970 (0.52%) |
|  | Bournemouth, Christchurch and Poole | 895 (0.74%) | 175,260 (0.72%) | 1,700 (0.65%) | 175,580 (0.72%) |
|  | Bracknell Forest | 325 (0.27%) | 52,770 (0.22%) | 600 (0.23%) | 52,870 (0.22%) |
|  | Bradford | 1,155 (0.96%) | 232,455 (0.96%) | 2,715 (1.04%) | 232,830 (0.96%) |
|  | Brent | 550 (0.46%) | 127,000 (0.52%) | 1,310 (0.50%) | 127,180 (0.52%) |
|  | Brighton and Hove | 695 (0.58%) | 119,930 (0.49%) | 1,160 (0.44%) | 120,045 (0.49%) |
|  | Bristol, City of | 895 (0.74%) | 190,820 (0.79%) | 2,090 (0.80%) | 191,215 (0.79%) |
|  | Bromley | 990 (0.82%) | 146,595 (0.60%) | 2,170 (0.83%) | 146,885 (0.60%) |
|  | Buckinghamshire | 950 (0.79%) | 235,445 (0.97%) | 2,150 (0.82%) | 235,765 (0.97%) |
|  | Bury | 350 (0.29%) | 86,425 (0.36%) | 785 (0.30%) | 86,500 (0.36%) |
|  | Calderdale | 405 (0.34%) | 94,520 (0.39%) | 805 (0.31%) | 94,615 (0.39%) |
|  | Cambridgeshire | 1,265 (1.05%) | 283,540 (1.17%) | 2,600 (0.99%) | 283,985 (1.17%) |
|  | Camden | 350 (0.29%) | 88,390 (0.36%) | 1,025 (0.39%) | 88,485 (0.36%) |
|  | Central Bedfordshire | 580 (0.48%) | 120,180 (0.50%) | 1,275 (0.49%) | 120,350 (0.50%) |
|  | Cheshire East | 750 (0.62%) | 175,670 (0.72%) | 1,680 (0.64%) | 175,875 (0.72%) |
|  | Cheshire West and Chester | 625 (0.52%) | 157,700 (0.65%) | 1,525 (0.58%) | 157,940 (0.65%) |
|  | City of London and Hackney | 480 (0.40%) | 102,855 (0.42%) | 1,125 (0.43%) | 102,985 (0.42%) |
|  | Cornwall and Isles of Scilly | 1,380 (1.15%) | 250,540 (1.03%) | 2,940 (1.12%) | 250,855 (1.03%) |
|  | County Durham | 1,040 (0.86%) | 242,005 (1.00%) | 2,300 (0.88%) | 242,380 (1.00%) |
|  | Coventry | 510 (0.42%) | 141,990 (0.59%) | 1,310 (0.50%) | 142,205 (0.59%) |
|  | Croydon | 940 (0.78%) | 160,200 (0.66%) | 1,845 (0.70%) | 160,395 (0.66%) |
|  | Cumbria | 810 (0.67%) | 234,850 (0.97%) | 1,860 (0.71%) | 235,140 (0.97%) |
|  | Darlington | 250 (0.21%) | 49,260 (0.20%) | 505 (0.19%) | 49,340 (0.20%) |
|  | Derby | 595 (0.49%) | 111,535 (0.46%) | 1,395 (0.53%) | 111,760 (0.46%) |
|  | Derbyshire | 1,805 (1.50%) | 363,400 (1.50%) | 4,260 (1.63%) | 364,130 (1.50%) |
|  | Devon | 1,740 (1.44%) | 358,045 (1.48%) | 3,930 (1.50%) | 358,720 (1.48%) |
|  | Doncaster | 515 (0.43%) | 137,725 (0.57%) | 1,250 (0.48%) | 137,950 (0.57%) |
|  | Dorset | 830 (0.69%) | 174,570 (0.72%) | 1,830 (0.70%) | 174,875 (0.72%) |
|  | Dudley | 735 (0.61%) | 146,635 (0.60%) | 1,420 (0.54%) | 146,795 (0.60%) |
|  | Ealing | 555 (0.46%) | 147,345 (0.61%) | 1,325 (0.51%) | 147,590 (0.61%) |
|  | East Riding of Yorkshire | 770 (0.64%) | 158,810 (0.65%) | 1,490 (0.57%) | 159,000 (0.65%) |
|  | East Sussex | 1,185 (0.98%) | 248,350 (1.02%) | 2,380 (0.91%) | 248,680 (1.02%) |
|  | Enfield | 525 (0.44%) | 138,605 (0.57%) | 1,305 (0.50%) | 138,790 (0.57%) |
|  | Essex | 3,125 (2.59%) | 658,515 (2.71%) | 6,890 (2.63%) | 659,535 (2.71%) |
|  | Gateshead | 320 (0.27%) | 92,460 (0.38%) | 845 (0.32%) | 92,565 (0.38%) |
|  | Gloucestershire | 1,365 (1.13%) | 281,195 (1.16%) | 2,835 (1.08%) | 281,680 (1.16%) |
|  | Greenwich | 690 (0.57%) | 109,980 (0.45%) | 1,355 (0.52%) | 110,160 (0.45%) |
|  | Halton | 250 (0.21%) | 57,990 (0.24%) | 805 (0.31%) | 58,090 (0.24%) |
|  | Hammersmith and Fulham | 320 (0.27%) | 75,135 (0.31%) | 735 (0.28%) | 75,235 (0.31%) |
|  | Hampshire | 3,680 (3.05%) | 621,790 (2.56%) | 6,755 (2.58%) | 622,745 (2.56%) |
|  | Haringey | 455 (0.38%) | 104,380 (0.43%) | 1,040 (0.40%) | 104,530 (0.43%) |
|  | Harrow | 490 (0.41%) | 106,875 (0.44%) | 1,095 (0.42%) | 107,030 (0.44%) |
|  | Hartlepool | 165 (0.14%) | 43,085 (0.18%) | 460 (0.18%) | 43,160 (0.18%) |
|  | Havering | 650 (0.54%) | 112,490 (0.46%) | 1,390 (0.53%) | 112,660 (0.46%) |
|  | Herefordshire, County of | 455 (0.38%) | 84,700 (0.35%) | 915 (0.35%) | 84,845 (0.35%) |
|  | Hertfordshire | 2,560 (2.12%) | 521,240 (2.15%) | 5,600 (2.14%) | 521,995 (2.15%) |
|  | Hillingdon | 525 (0.44%) | 122,340 (0.50%) | 1,185 (0.45%) | 122,505 (0.50%) |
|  | Hounslow | 475 (0.39%) | 110,670 (0.46%) | 1,015 (0.39%) | 110,815 (0.46%) |
|  | Isle of Wight | 245 (0.20%) | 65,125 (0.27%) | 705 (0.27%) | 65,215 (0.27%) |
|  | Islington | 360 (0.30%) | 87,890 (0.36%) | 1,085 (0.41%) | 88,025 (0.36%) |
|  | Kensington and Chelsea | 195 (0.16%) | 54,560 (0.22%) | 460 (0.18%) | 54,625 (0.22%) |
|  | Kent | 3,655 (3.03%) | 681,010 (2.81%) | 7,490 (2.86%) | 682,085 (2.81%) |
|  | Kingston upon Hull, City of | 840 (0.70%) | 115,170 (0.47%) | 1,385 (0.53%) | 115,340 (0.47%) |
|  | Kingston upon Thames | 375 (0.31%) | 72,370 (0.30%) | 740 (0.28%) | 72,495 (0.30%) |
|  | Kirklees | 990 (0.82%) | 191,795 (0.79%) | 1,970 (0.75%) | 192,000 (0.79%) |
|  | Knowsley | 445 (0.37%) | 68,470 (0.28%) | 1,190 (0.45%) | 68,605 (0.28%) |
|  | Lambeth | 595 (0.49%) | 123,625 (0.51%) | 1,330 (0.51%) | 123,770 (0.51%) |
|  | Lancashire | 2,870 (2.38%) | 544,920 (2.25%) | 6,445 (2.46%) | 545,935 (2.25%) |
|  | Leeds | 1,320 (1.10%) | 345,075 (1.42%) | 2,830 (1.08%) | 345,540 (1.42%) |
|  | Leicester | 620 (0.51%) | 145,595 (0.60%) | 1,495 (0.57%) | 145,815 (0.60%) |
|  | Leicestershire | 1,270 (1.05%) | 305,565 (1.26%) | 3,200 (1.22%) | 306,060 (1.26%) |
|  | Lewisham | 700 (0.58%) | 117,670 (0.49%) | 1,495 (0.57%) | 117,860 (0.49%) |
|  | Lincolnshire | 1,795 (1.49%) | 337,900 (1.39%) | 3,725 (1.42%) | 338,540 (1.39%) |
|  | Liverpool | 1,210 (1.00%) | 200,500 (0.83%) | 3,045 (1.16%) | 200,845 (0.83%) |
|  | Luton | 490 (0.41%) | 89,265 (0.37%) | 1,060 (0.40%) | 89,365 (0.37%) |
|  | Manchester | 1,105 (0.92%) | 206,460 (0.85%) | 2,310 (0.88%) | 206,730 (0.85%) |
|  | Medway | 790 (0.66%) | 119,735 (0.49%) | 2,010 (0.77%) | 119,955 (0.49%) |
|  | Merton | 465 (0.39%) | 89,985 (0.37%) | 895 (0.34%) | 90,080 (0.37%) |
|  | Middlesbrough | 335 (0.28%) | 63,265 (0.26%) | 590 (0.23%) | 63,355 (0.26%) |
|  | Milton Keynes | 645 (0.54%) | 112,765 (0.46%) | 1,420 (0.54%) | 112,895 (0.46%) |
|  | Newcastle upon Tyne | 505 (0.42%) | 122,395 (0.50%) | 1,220 (0.47%) | 122,510 (0.50%) |
|  | Newham | 635 (0.53%) | 119,640 (0.49%) | 1,395 (0.53%) | 119,810 (0.49%) |
|  | Norfolk | 1,625 (1.35%) | 403,250 (1.66%) | 3,865 (1.47%) | 403,710 (1.66%) |
|  | North East Lincolnshire | 360 (0.30%) | 73,225 (0.30%) | 695 (0.27%) | 73,340 (0.30%) |
|  | North Lincolnshire | 370 (0.31%) | 76,865 (0.32%) | 665 (0.25%) | 76,955 (0.32%) |
|  | North Northamptonshire | 940 (0.78%) | 148,735 (0.61%) | 1,845 (0.70%) | 148,930 (0.61%) |
|  | North Somerset | 425 (0.35%) | 97,550 (0.40%) | 1,050 (0.40%) | 97,785 (0.40%) |
|  | North Tyneside | 350 (0.29%) | 96,240 (0.40%) | 850 (0.32%) | 96,350 (0.40%) |
|  | North Yorkshire | 980 (0.81%) | 279,495 (1.15%) | 2,185 (0.83%) | 279,860 (1.15%) |
|  | Northumberland | 455 (0.38%) | 149,555 (0.62%) | 1,100 (0.42%) | 149,735 (0.62%) |
|  | Nottingham | 650 (0.54%) | 130,230 (0.54%) | 1,665 (0.64%) | 130,425 (0.54%) |
|  | Nottinghamshire | 1,740 (1.44%) | 368,245 (1.52%) | 4,285 (1.63%) | 369,130 (1.52%) |
|  | Oldham | 655 (0.54%) | 102,150 (0.42%) | 1,315 (0.50%) | 102,400 (0.42%) |
|  | Oxfordshire | 1,415 (1.17%) | 301,550 (1.24%) | 3,065 (1.17%) | 301,905 (1.24%) |
|  | Peterborough | 520 (0.43%) | 83,060 (0.34%) | 945 (0.36%) | 83,190 (0.34%) |
|  | Plymouth | 515 (0.43%) | 117,350 (0.48%) | 1,265 (0.48%) | 117,490 (0.48%) |
|  | Portsmouth | 585 (0.49%) | 88,585 (0.37%) | 1,050 (0.40%) | 88,685 (0.36%) |
|  | Reading | 280 (0.23%) | 70,580 (0.29%) | 620 (0.24%) | 70,710 (0.29%) |
|  | Redbridge | 600 (0.50%) | 123,345 (0.51%) | 1,320 (0.50%) | 123,510 (0.51%) |
|  | Redcar and Cleveland | 350 (0.29%) | 64,230 (0.26%) | 645 (0.25%) | 64,320 (0.26%) |
|  | Richmond upon Thames | 330 (0.27%) | 86,285 (0.36%) | 695 (0.27%) | 86,400 (0.36%) |
|  | Rochdale | 615 (0.51%) | 97,135 (0.40%) | 1,165 (0.44%) | 97,290 (0.40%) |
|  | Rotherham | 740 (0.61%) | 120,825 (0.50%) | 1,575 (0.60%) | 121,035 (0.50%) |
|  | Rutland | 60 (0.05%) | 16,995 (0.07%) | 135 (0.05%) | 17,015 (0.07%) |
|  | Salford | 455 (0.38%) | 104,525 (0.43%) | 1,060 (0.40%) | 104,645 (0.43%) |
|  | Sandwell | 600 (0.50%) | 140,660 (0.58%) | 1,300 (0.50%) | 140,810 (0.58%) |
|  | Sefton | 680 (0.56%) | 128,635 (0.53%) | 1,710 (0.65%) | 128,855 (0.53%) |
|  | Sheffield | 1,200 (1.00%) | 242,790 (1.00%) | 2,280 (0.87%) | 243,070 (1.00%) |
|  | Shropshire | 380 (0.32%) | 140,500 (0.58%) | 1,185 (0.45%) | 140,725 (0.58%) |
|  | Slough | 355 (0.29%) | 60,925 (0.25%) | 680 (0.26%) | 60,995 (0.25%) |
|  | Solihull | 345 (0.29%) | 98,685 (0.41%) | 815 (0.31%) | 98,785 (0.41%) |
|  | Somerset | 1,140 (0.95%) | 252,655 (1.04%) | 2,360 (0.90%) | 253,030 (1.04%) |
|  | South Gloucestershire | 630 (0.52%) | 123,315 (0.51%) | 1,520 (0.58%) | 123,575 (0.51%) |
|  | South Tyneside | 415 (0.34%) | 69,960 (0.29%) | 885 (0.34%) | 70,110 (0.29%) |
|  | Southampton | 790 (0.66%) | 103,960 (0.43%) | 1,550 (0.59%) | 104,110 (0.43%) |
|  | Southend-on-Sea | 250 (0.21%) | 79,755 (0.33%) | 645 (0.25%) | 79,850 (0.33%) |
|  | Southwark | 560 (0.46%) | 116,420 (0.48%) | 1,290 (0.49%) | 116,545 (0.48%) |
|  | St. Helens | 355 (0.29%) | 82,340 (0.34%) | 1,055 (0.40%) | 82,505 (0.34%) |
|  | Staffordshire | 2,315 (1.92%) | 394,545 (1.63%) | 4,825 (1.84%) | 395,170 (1.63%) |
|  | Stockport | 860 (0.71%) | 134,600 (0.55%) | 1,725 (0.66%) | 134,855 (0.55%) |
|  | Stockton-on-Tees | 420 (0.35%) | 89,085 (0.37%) | 1,020 (0.39%) | 89,215 (0.37%) |
|  | Stoke-on-Trent | 885 (0.73%) | 112,915 (0.47%) | 1,695 (0.65%) | 113,105 (0.47%) |
|  | Suffolk | 1,215 (1.01%) | 337,000 (1.39%) | 2,875 (1.10%) | 337,400 (1.39%) |
|  | Sunderland | 585 (0.49%) | 130,340 (0.54%) | 1,330 (0.51%) | 130,545 (0.54%) |
|  | Surrey | 3,460 (2.87%) | 527,210 (2.17%) | 6,195 (2.36%) | 528,065 (2.17%) |
|  | Sutton | 495 (0.41%) | 89,620 (0.37%) | 1,000 (0.38%) | 89,725 (0.37%) |
|  | Swindon | 830 (0.69%) | 95,790 (0.39%) | 1,580 (0.60%) | 96,035 (0.40%) |
|  | Tameside | 615 (0.51%) | 103,565 (0.43%) | 1,360 (0.52%) | 103,765 (0.43%) |
|  | Telford and Wrekin | 240 (0.20%) | 76,145 (0.31%) | 775 (0.30%) | 76,300 (0.31%) |
|  | Thurrock | 340 (0.28%) | 71,665 (0.30%) | 780 (0.30%) | 71,805 (0.30%) |
|  | Torbay | 450 (0.37%) | 62,140 (0.26%) | 870 (0.33%) | 62,270 (0.26%) |
|  | Tower Hamlets | 620 (0.51%) | 102,250 (0.42%) | 1,295 (0.49%) | 102,380 (0.42%) |
|  | Trafford | 560 (0.46%) | 104,425 (0.43%) | 1,155 (0.44%) | 104,640 (0.43%) |
|  | Wakefield | 900 (0.75%) | 153,160 (0.63%) | 1,980 (0.76%) | 153,415 (0.63%) |
|  | Walsall | 855 (0.71%) | 122,540 (0.51%) | 1,640 (0.63%) | 122,735 (0.51%) |
|  | Waltham Forest | 545 (0.45%) | 109,395 (0.45%) | 1,230 (0.47%) | 109,550 (0.45%) |
|  | Wandsworth | 665 (0.55%) | 136,600 (0.56%) | 1,390 (0.53%) | 136,815 (0.56%) |
|  | Warrington | 385 (0.32%) | 94,620 (0.39%) | 1,125 (0.43%) | 94,785 (0.39%) |
|  | Warwickshire | 1,065 (0.88%) | 255,820 (1.05%) | 2,655 (1.01%) | 256,215 (1.05%) |
|  | West Berkshire | 330 (0.27%) | 72,890 (0.30%) | 680 (0.26%) | 73,030 (0.30%) |
|  | West Northamptonshire | 990 (0.82%) | 174,350 (0.72%) | 2,220 (0.85%) | 174,590 (0.72%) |
|  | West Sussex | 2,110 (1.75%) | 384,005 (1.58%) | 4,125 (1.57%) | 384,640 (1.58%) |
|  | Westminster | 260 (0.22%) | 80,115 (0.33%) | 685 (0.26%) | 80,205 (0.33%) |
|  | Wigan | 680 (0.56%) | 147,145 (0.61%) | 1,485 (0.57%) | 147,345 (0.61%) |
|  | Wiltshire | 1,105 (0.92%) | 217,680 (0.90%) | 2,315 (0.88%) | 218,025 (0.90%) |
|  | Windsor and Maidenhead | 270 (0.22%) | 66,605 (0.27%) | 550 (0.21%) | 66,685 (0.27%) |
|  | Wirral | 1,230 (1.02%) | 150,225 (0.62%) | 2,350 (0.90%) | 150,425 (0.62%) |
|  | Wokingham | 230 (0.19%) | 73,480 (0.30%) | 540 (0.21%) | 73,600 (0.30%) |
|  | Wolverhampton | 585 (0.49%) | 110,920 (0.46%) | 1,440 (0.55%) | 111,135 (0.46%) |
|  | Worcestershire | 1,825 (1.51%) | 267,505 (1.10%) | 3,425 (1.31%) | 267,960 (1.10%) |
|  | York | 410 (0.34%) | 91,965 (0.38%) | 800 (0.31%) | 92,085 (0.38%) |

**Supplementary Table 8: Age-standardised rates of endometriosis diagnosis per 100,000 people by upper tier local authority (UTLA)**

| **Subcategory** | **Main analysis,**  **Age-standardised rate [95% CI]** | **Secondary analysis,**  **Age-standardised rate [95% CI]** |
| --- | --- | --- |
| Barking and Dagenham | 461.17 [417.53, 504.81] | 987.22 [922.59, 1,051.84] |
| Barnet | 283.17 [257.77, 308.57] | 780.10 [737.50, 822.71] |
| Barnsley | 524.06 [480.53, 567.59] | 1,192.61 [1,127.59, 1,257.63] |
| Bath and North East Somerset | 374.99 [332.37, 417.61] | 815.55 [752.78, 878.33] |
| Bedford | 466.67 [417.67, 515.66] | 911.66 [843.29, 980.03] |
| Bexley | 641.05 [593.46, 688.65] | 1,312.49 [1,244.77, 1,380.21] |
| Birmingham | 359.79 [343.39, 376.20] | 992.81 [964.78, 1,020.84] |
| Blackburn with Darwen | 577.70 [521.33, 634.08] | 1,237.64 [1,154.17, 1,321.10] |
| Blackpool | 575.18 [515.66, 634.70] | 1,256.85 [1,169.63, 1,344.07] |
| Bolton | 417.00 [381.51, 452.48] | 989.95 [935.44, 1,044.46] |
| Bournemouth, Christchurch and Poole | 532.87 [497.86, 567.88] | 1,004.41 [956.58, 1,052.25] |
| Bracknell Forest | 579.28 [515.70, 642.87] | 1,060.62 [974.98, 1,146.26] |
| Bradford | 476.10 [448.53, 503.67] | 1,144.25 [1,101.04, 1,187.47] |
| Brent | 362.35 [331.48, 393.21] | 899.57 [849.87, 949.27] |
| Brighton and Hove | 496.51 [459.21, 533.81] | 832.90 [784.34, 881.46] |
| Bristol, City of | 397.27 [370.62, 423.91] | 977.70 [934.79, 1,020.62] |
| Bromley | 701.37 [657.35, 745.39] | 1,489.30 [1,426.18, 1,552.42] |
| Buckinghamshire | 434.28 [406.40, 462.17] | 953.80 [913.05, 994.54] |
| Bury | 416.11 [372.50, 459.71] | 917.43 [853.23, 981.64] |
| Calderdale | 443.23 [399.89, 486.57] | 867.12 [807.08, 927.16] |
| Cambridgeshire | 440.55 [416.28, 464.83] | 899.29 [864.71, 933.86] |
| Camden | 307.20 [273.23, 341.17] | 986.14 [922.38, 1,049.89] |
| Central Bedfordshire | 501.30 [460.20, 542.39] | 1,072.66 [1,013.42, 1,131.90] |
| Cheshire East | 476.31 [441.90, 510.72] | 1,026.22 [976.62, 1,075.82] |
| Cheshire West and Chester | 421.34 [388.25, 454.43] | 996.54 [946.33, 1,046.74] |
| City of London and Hackney | 348.01 [315.00, 381.02] | 868.07 [813.52, 922.63] |
| Cornwall and Isles of Scilly | 649.89 [615.43, 684.36] | 1,319.20 [1,271.05, 1,367.34] |
| County Durham | 451.09 [423.62, 478.57] | 974.61 [934.62, 1,014.60] |
| Coventry | 339.96 [310.31, 369.60] | 892.61 [843.95, 941.28] |
| Croydon | 541.96 [507.17, 576.74] | 1,068.55 [1,019.59, 1,117.51] |
| Cumbria | 392.52 [365.29, 419.76] | 856.82 [817.46, 896.18] |
| Darlington | 531.15 [465.25, 597.05] | 1,051.80 [960.04, 1,143.56] |
| Derby | 508.26 [467.30, 549.22] | 1,209.14 [1,145.48, 1,272.81] |
| Derbyshire | 539.26 [514.25, 564.28] | 1,211.00 [1,174.38, 1,247.63] |
| Devon | 577.93 [550.57, 605.30] | 1,221.42 [1,182.71, 1,260.12] |
| Doncaster | 391.13 [357.30, 424.97] | 932.86 [881.08, 984.64] |
| Dorset | 622.95 [579.86, 666.04] | 1,259.67 [1,200.38, 1,318.97] |
| Dudley | 526.01 [487.92, 564.09] | 997.79 [945.76, 1,049.81] |
| Ealing | 317.10 [290.20, 344.00] | 787.60 [744.18, 831.02] |
| East Riding of Yorkshire | 576.87 [535.57, 618.17] | 1,044.92 [990.84, 1,098.99] |
| East Sussex | 586.29 [552.60, 619.98] | 1,122.68 [1,076.97, 1,168.38] |
| Enfield | 346.70 [316.96, 376.44] | 870.36 [822.89, 917.83] |
| Essex | 505.48 [487.70, 523.26] | 1,090.14 [1,064.31, 1,115.97] |
| Gateshead | 351.09 [312.53, 389.64] | 915.80 [854.03, 977.57] |
| Gloucestershire | 528.68 [500.53, 556.83] | 1,067.94 [1,028.45, 1,107.43] |
| Greenwich | 522.14 [482.70, 561.58] | 1,060.56 [1,002.85, 1,118.27] |
| Halton | 429.62 [376.34, 482.91] | 1,363.49 [1,269.31, 1,457.66] |
| Hammersmith and Fulham | 316.10 [278.80, 353.39] | 784.87 [723.51, 846.24] |
| Hampshire | 641.58 [620.73, 662.43] | 1,147.95 [1,120.40, 1,175.49] |
| Haringey | 341.65 [309.47, 373.84] | 832.59 [780.42, 884.76] |
| Harrow | 427.80 [389.65, 465.96] | 965.82 [908.33, 1,023.32] |
| Hartlepool | 406.11 [343.93, 468.30] | 1,097.35 [996.44, 1,198.27] |
| Havering | 598.80 [552.79, 644.81] | 1,259.25 [1,192.94, 1,325.57] |
| Herefordshire, County of | 637.21 [578.08, 696.34] | 1,240.88 [1,159.65, 1,322.11] |
| Hertfordshire | 493.94 [474.71, 513.16] | 1,061.55 [1,033.63, 1,089.46] |
| Hillingdon | 389.51 [356.03, 422.99] | 898.63 [847.21, 950.05] |
| Hounslow | 354.54 [322.02, 387.07] | 801.39 [750.96, 851.82] |
| Isle of Wight | 473.98 [414.08, 533.88] | 1,229.66 [1,136.99, 1,322.34] |
| Islington | 312.39 [277.85, 346.93] | 1,023.26 [957.71, 1,088.82] |
| Kensington and Chelsea | 288.87 [246.70, 331.05] | 718.40 [650.38, 786.42] |
| Kent | 567.41 [548.97, 585.85] | 1,142.36 [1,116.42, 1,168.31] |
| Kingston upon Hull, City of | 670.17 [624.44, 715.91] | 1,131.70 [1,071.61, 1,191.80] |
| Kingston upon Thames | 455.30 [408.89, 501.72] | 915.82 [849.30, 982.34] |
| Kirklees | 510.06 [478.29, 541.83] | 1,013.37 [968.61, 1,058.13] |
| Knowsley | 644.57 [584.38, 704.77] | 1,691.20 [1,594.73, 1,787.68] |
| Lambeth | 357.10 [326.64, 387.56] | 850.98 [801.62, 900.33] |
| Lancashire | 552.39 [532.12, 572.66] | 1,214.46 [1,184.72, 1,244.20] |
| Leeds | 345.13 [326.32, 363.94] | 759.41 [731.12, 787.69] |
| Leicester | 367.01 [337.56, 396.46] | 938.51 [889.80, 987.21] |
| Leicestershire | 439.70 [415.41, 463.99] | 1,069.92 [1,032.65, 1,107.18] |
| Lewisham | 491.10 [454.13, 528.07] | 1,061.91 [1,006.87, 1,116.95] |
| Lincolnshire | 589.73 [562.32, 617.14] | 1,179.34 [1,141.21, 1,217.46] |
| Liverpool | 564.71 [532.43, 597.00] | 1,454.18 [1,401.99, 1,506.37] |
| Luton | 489.33 [445.64, 533.02] | 1,094.27 [1,027.64, 1,160.89] |
| Manchester | 418.21 [392.48, 443.95] | 938.48 [898.32, 978.63] |
| Medway | 642.53 [597.70, 687.37] | 1,617.80 [1,547.03, 1,688.56] |
| Merton | 444.94 [403.38, 486.50] | 862.19 [804.01, 920.37] |
| Middlesbrough | 521.02 [465.04, 577.00] | 916.26 [841.84, 990.68] |
| Milton Keynes | 524.90 [484.06, 565.74] | 1,168.70 [1,107.42, 1,229.99] |
| Newcastle upon Tyne | 381.86 [347.74, 415.98] | 973.04 [917.45, 1,028.64] |
| Newham | 418.75 [385.12, 452.38] | 1,003.42 [947.97, 1,058.87] |
| Norfolk | 452.43 [430.39, 474.47] | 1,038.28 [1,005.38, 1,071.17] |
| North East Lincolnshire | 509.42 [456.50, 562.35] | 979.71 [906.65, 1,052.78] |
| North Lincolnshire | 525.44 [471.84, 579.03] | 911.59 [841.91, 981.27] |
| North Northamptonshire | 642.87 [601.68, 684.06] | 1,238.46 [1,181.76, 1,295.15] |
| North Somerset | 493.82 [446.61, 541.04] | 1,164.82 [1,093.75, 1,235.89] |
| North Tyneside | 374.25 [334.91, 413.60] | 888.33 [828.45, 948.21] |
| North Yorkshire | 414.92 [388.60, 441.24] | 874.88 [837.58, 912.19] |
| Northumberland | 356.37 [323.36, 389.37] | 812.49 [763.81, 861.18] |
| Nottingham | 415.52 [382.18, 448.85] | 1,169.36 [1,110.84, 1,227.88] |
| Nottinghamshire | 504.62 [480.84, 528.40] | 1,188.85 [1,153.09, 1,224.61] |
| Oldham | 642.40 [593.21, 691.59] | 1,282.46 [1,213.07, 1,351.85] |
| Oxfordshire | 459.46 [435.53, 483.39] | 989.99 [954.94, 1,025.05] |
| Peterborough | 583.42 [533.08, 633.77] | 1,074.06 [1,005.34, 1,142.77] |
| Plymouth | 418.49 [382.02, 454.97] | 1,056.28 [997.76, 1,114.79] |
| Portsmouth | 590.55 [542.10, 639.00] | 1,073.63 [1,007.80, 1,139.46] |
| Reading | 321.20 [282.88, 359.53] | 752.02 [690.99, 813.06] |
| Redbridge | 437.38 [402.06, 472.70] | 982.39 [928.81, 1,035.98] |
| Redcar and Cleveland | 605.17 [541.46, 668.88] | 1,079.27 [995.53, 1,163.00] |
| Richmond upon Thames | 367.28 [326.52, 408.04] | 752.59 [695.16, 810.01] |
| Rochdale | 621.16 [572.03, 670.29] | 1,175.48 [1,107.90, 1,243.06] |
| Rotherham | 633.58 [587.84, 679.31] | 1,324.41 [1,258.91, 1,389.90] |
| Rutland | 447.54 [338.65, 579.56] | 956.04 [790.72, 1,121.35] |
| Salford | 397.44 [360.71, 434.17] | 938.05 [881.08, 995.02] |
| Sandwell | 404.72 [372.32, 437.12] | 886.34 [838.07, 934.61] |
| Sefton | 599.58 [554.22, 644.93] | 1,429.49 [1,361.15, 1,497.83] |
| Sheffield | 467.52 [440.81, 494.23] | 899.04 [861.82, 936.26] |
| Shropshire | 314.82 [282.86, 346.79] | 908.81 [856.20, 961.42] |
| Slough | 467.92 [418.40, 517.43] | 928.69 [856.96, 1,000.43] |
| Solihull | 388.44 [347.13, 429.76] | 892.40 [830.53, 954.26] |
| Somerset | 526.39 [495.53, 557.25] | 1,052.00 [1,009.09, 1,094.92] |
| South Gloucestershire | 520.88 [480.08, 561.69] | 1,224.81 [1,163.02, 1,286.61] |
| South Tyneside | 629.77 [569.14, 690.40] | 1,305.79 [1,219.43, 1,392.15] |
| Southampton | 650.52 [603.54, 697.49] | 1,356.68 [1,286.71, 1,426.66] |
| Southend-on-Sea | 324.95 [284.79, 365.11] | 818.37 [755.12, 881.63] |
| Southwark | 368.12 [336.35, 399.89] | 891.73 [840.48, 942.98] |
| St. Helens | 450.87 [403.83, 497.92] | 1,298.18 [1,219.65, 1,376.71] |
| Staffordshire | 637.31 [611.24, 663.38] | 1,279.71 [1,243.39, 1,316.04] |
| Stockport | 680.55 [634.89, 726.20] | 1,328.60 [1,265.72, 1,391.48] |
| Stockton-on-Tees | 483.34 [437.18, 529.49] | 1,144.39 [1,074.04, 1,214.74] |
| Stoke-on-Trent | 759.81 [709.55, 810.07] | 1,466.43 [1,396.41, 1,536.44] |
| Suffolk | 404.17 [381.36, 426.98] | 920.22 [886.40, 954.03] |
| Sunderland | 461.47 [424.03, 498.90] | 1,028.51 [973.15, 1,083.87] |
| Surrey | 682.93 [660.01, 705.84] | 1,202.05 [1,171.90, 1,232.19] |
| Sutton | 528.41 [481.62, 575.20] | 1,057.60 [991.73, 1,123.48] |
| Swindon | 823.19 [767.13, 879.24] | 1,540.25 [1,464.09, 1,616.41] |
| Tameside | 588.88 [542.40, 635.36] | 1,282.67 [1,214.45, 1,350.89] |
| Telford and Wrekin | 313.32 [273.44, 353.19] | 996.55 [926.27, 1,066.83] |
| Thurrock | 443.13 [396.00, 490.26] | 1,017.87 [946.22, 1,089.53] |
| Torbay | 874.48 [793.41, 955.55] | 1,615.92 [1,507.66, 1,724.18] |
| Tower Hamlets | 416.70 [380.20, 453.20] | 991.08 [929.37, 1,052.80] |
| Trafford | 544.87 [499.40, 590.35] | 1,103.45 [1,039.49, 1,167.41] |
| Wakefield | 598.49 [559.39, 637.59] | 1,287.02 [1,230.22, 1,343.81] |
| Walsall | 709.55 [661.92, 757.18] | 1,354.20 [1,288.58, 1,419.83] |
| Waltham Forest | 422.22 [386.33, 458.11] | 970.41 [915.33, 1,025.48] |
| Wandsworth | 340.27 [311.74, 368.80] | 755.29 [711.22, 799.37] |
| Warrington | 414.57 [373.09, 456.05] | 1,172.92 [1,104.16, 1,241.67] |
| Warwickshire | 438.49 [412.12, 464.87] | 1,064.19 [1,023.58, 1,104.80] |
| West Berkshire | 463.44 [412.98, 513.89] | 929.02 [858.35, 999.69] |
| West Northamptonshire | 561.80 [526.72, 596.87] | 1,244.99 [1,193.15, 1,296.84] |
| West Sussex | 622.31 [595.62, 649.00] | 1,178.83 [1,142.65, 1,215.02] |
| Westminster | 251.68 [219.71, 283.66] | 681.57 [628.04, 735.11] |
| Wigan | 469.05 [433.71, 504.39] | 1,009.89 [958.48, 1,061.30] |
| Wiltshire | 565.34 [531.55, 599.13] | 1,134.49 [1,087.60, 1,181.38] |
| Windsor and Maidenhead | 420.72 [370.00, 471.44] | 831.39 [760.91, 901.87] |
| Wirral | 897.81 [847.54, 948.08] | 1,666.85 [1,599.20, 1,734.50] |
| Wokingham | 326.83 [283.81, 369.86] | 742.98 [679.24, 806.73] |
| Wolverhampton | 516.62 [474.71, 558.53] | 1,277.74 [1,211.71, 1,343.76] |
| Worcestershire | 753.65 [718.87, 788.42] | 1,372.27 [1,326.02, 1,418.53] |
| York | 427.17 [385.40, 468.94] | 833.40 [774.95, 891.84] |

**Supplementary Table 9: Odds ratios for endometriosis diagnosis and 95% confidence intervals by region**

| **Analysis type** | **Term** | **Adjusted for age,**  **OR [95% CI]** | **Adjusted for age and health,**  **OR [95% CI]** | **Adjusted for age, health and IMD decile,**  **OR [95% CI]** | **Adjusted for age, health, IMD decile and highest level of qualification,**  **OR [95% CI]** |
| --- | --- | --- | --- | --- | --- |
| Main analysis | North East | 1.09 [1.06, 1.12] | 1.05 [1.02, 1.09] | 1.07 [1.04, 1.10] | 1.02 [0.99, 1.06] |
|  | North West | 1.29 [1.27, 1.32] | 1.25 [1.22, 1.28] | 1.27 [1.24, 1.30] | 1.22 [1.20, 1.25] |
|  | Yorkshire and the Humber | 1.18 [1.15, 1.20] | 1.14 [1.12, 1.17] | 1.16 [1.13, 1.19] | 1.12 [1.09, 1.15] |
|  | East Midlands | 1.25 [1.22, 1.28] | 1.22 [1.19, 1.25] | 1.24 [1.21, 1.27] | 1.19 [1.16, 1.22] |
|  | West Midlands | 1.24 [1.21, 1.27] | 1.21 [1.18, 1.24] | 1.23 [1.20, 1.26] | 1.19 [1.16, 1.21] |
|  | East of England | 1.16 [1.13, 1.19] | 1.14 [1.12, 1.17] | 1.16 [1.13, 1.18] | 1.11 [1.08, 1.13] |
|  | South East | 1.40 [1.37, 1.43] | 1.39 [1.36, 1.42] | 1.41 [1.39, 1.44] | 1.36 [1.34, 1.39] |
|  | South West | 1.35 [1.32, 1.39] | 1.32 [1.29, 1.35] | 1.34 [1.30, 1.37] | 1.28 [1.25, 1.31] |
| Secondary analysis | North East | 1.06 [1.04, 1.08] | 1.02 [1.00, 1.05] | 1.04 [1.01, 1.06] | 1.00 [0.98, 1.02] |
|  | North West | 1.27 [1.25, 1.29] | 1.23 [1.21, 1.24] | 1.24 [1.22, 1.26] | 1.20 [1.19, 1.22] |
|  | Yorkshire and the Humber | 1.06 [1.04, 1.08] | 1.03 [1.02, 1.05] | 1.05 [1.03, 1.07] | 1.02 [1.00, 1.03] |
|  | East Midlands | 1.25 [1.23, 1.27] | 1.22 [1.20, 1.24] | 1.24 [1.22, 1.26] | 1.20 [1.18, 1.22] |
|  | West Midlands | 1.20 [1.18, 1.22] | 1.17 [1.16, 1.19] | 1.19 [1.17, 1.21] | 1.15 [1.14, 1.17] |
|  | East of England | 1.10 [1.08, 1.11] | 1.08 [1.07, 1.10] | 1.10 [1.08, 1.12] | 1.07 [1.05, 1.08] |
|  | South East | 1.19 [1.17, 1.21] | 1.19 [1.17, 1.20] | 1.21 [1.20, 1.23] | 1.18 [1.16, 1.20] |
|  | South West | 1.24 [1.22, 1.26] | 1.21 [1.20, 1.23] | 1.23 [1.21, 1.25] | 1.19 [1.17, 1.21] |

Notes: Reference category: London

**Supplementary Table 10: Odds ratios for endometriosis diagnosis and 95% confidence intervals by upper tier local authority (UTLA)**

| **Analysis type** | **Term** | **Adjusted for age,**  **OR [95% CI]** | **Adjusted for age and health,**  **OR [95% CI]** |
| --- | --- | --- | --- |
| Main analysis | Barking and Dagenham | 1.34 [1.18, 1.53] | 1.31 [1.15, 1.49] |
|  | Barnet | 0.83 [0.73, 0.94] | 0.82 [0.72, 0.93] |
|  | Barnsley | 1.55 [1.37, 1.75] | 1.47 [1.30, 1.66] |
|  | Bath and North East Somerset | 1.08 [0.94, 1.25] | 1.08 [0.94, 1.25] |
|  | Bedford | 1.37 [1.19, 1.57] | 1.35 [1.17, 1.55] |
|  | Bexley | 1.89 [1.68, 2.12] | 1.85 [1.64, 2.08] |
|  | Birmingham | 1.05 [0.95, 1.16] | 1.02 [0.92, 1.13] |
|  | Blackburn with Darwen | 1.69 [1.48, 1.93] | 1.62 [1.42, 1.85] |
|  | Blackpool | 1.70 [1.48, 1.95] | 1.62 [1.41, 1.86] |
|  | Bolton | 1.22 [1.08, 1.38] | 1.18 [1.04, 1.33] |
|  | Bournemouth, Christchurch and Poole | 1.55 [1.39, 1.74] | 1.51 [1.35, 1.69] |
|  | Bracknell Forest | 1.69 [1.47, 1.95] | 1.68 [1.46, 1.94] |
|  | Bradford | 1.39 [1.25, 1.54] | 1.31 [1.18, 1.46] |
|  | Brent | 1.05 [0.93, 1.19] | 1.06 [0.93, 1.19] |
|  | Brighton and Hove | 1.43 [1.27, 1.61] | 1.44 [1.28, 1.62] |
|  | Bristol, City of | 1.15 [1.03, 1.28] | 1.13 [1.01, 1.26] |
|  | Bromley | 2.04 [1.83, 2.28] | 1.98 [1.77, 2.20] |
|  | Buckinghamshire | 1.26 [1.13, 1.41] | 1.26 [1.12, 1.40] |
|  | Bury | 1.22 [1.06, 1.40] | 1.18 [1.03, 1.35] |
|  | Calderdale | 1.29 [1.13, 1.47] | 1.24 [1.09, 1.42] |
|  | Cambridgeshire | 1.29 [1.16, 1.43] | 1.28 [1.15, 1.42] |
|  | Camden | 0.86 [0.75, 0.98] | 0.88 [0.76, 1.01] |
|  | Central Bedfordshire | 1.46 [1.29, 1.65] | 1.43 [1.26, 1.61] |
|  | Cheshire East | 1.39 [1.24, 1.56] | 1.35 [1.21, 1.52] |
|  | Cheshire West and Chester | 1.23 [1.09, 1.38] | 1.18 [1.05, 1.33] |
|  | Cornwall and Isles of Scilly | 1.90 [1.71, 2.11] | 1.82 [1.64, 2.02] |
|  | County Durham | 1.31 [1.18, 1.46] | 1.26 [1.13, 1.41] |
|  | Coventry | 0.97 [0.86, 1.10] | 0.95 [0.83, 1.07] |
|  | Croydon | 1.58 [1.42, 1.77] | 1.54 [1.38, 1.72] |
|  | Cumbria | 1.15 [1.03, 1.29] | 1.13 [1.00, 1.26] |
|  | Darlington | 1.56 [1.34, 1.82] | 1.50 [1.29, 1.75] |
|  | Derby | 1.47 [1.31, 1.66] | 1.41 [1.25, 1.59] |
|  | Derbyshire | 1.58 [1.43, 1.75] | 1.53 [1.38, 1.69] |
|  | Devon | 1.69 [1.53, 1.87] | 1.65 [1.49, 1.82] |
|  | Doncaster | 1.14 [1.01, 1.29] | 1.11 [0.98, 1.26] |
|  | Dorset | 1.81 [1.62, 2.03] | 1.76 [1.57, 1.97] |
|  | Dudley | 1.54 [1.37, 1.73] | 1.47 [1.31, 1.65] |
|  | Ealing | 0.93 [0.82, 1.05] | 0.92 [0.82, 1.04] |
|  | East Riding of Yorkshire | 1.68 [1.50, 1.89] | 1.63 [1.46, 1.83] |
|  | East Sussex | 1.70 [1.53, 1.90] | 1.67 [1.50, 1.86] |
|  | Enfield | 1.01 [0.90, 1.15] | 1.00 [0.88, 1.13] |
|  | Essex | 1.49 [1.35, 1.64] | 1.46 [1.32, 1.61] |
|  | Gateshead | 1.03 [0.89, 1.19] | 1.00 [0.87, 1.16] |
|  | Gloucestershire | 1.54 [1.39, 1.71] | 1.50 [1.36, 1.67] |
|  | Greenwich | 1.53 [1.36, 1.72] | 1.51 [1.34, 1.69] |
|  | Halton | 1.26 [1.08, 1.47] | 1.20 [1.03, 1.40] |
|  | Hammersmith and Fulham | 0.91 [0.79, 1.04] | 0.92 [0.80, 1.06] |
|  | Hampshire | 1.88 [1.71, 2.07] | 1.85 [1.69, 2.04] |
|  | Haringey | 1.01 [0.89, 1.15] | 1.02 [0.89, 1.16] |
|  | Harrow | 1.26 [1.11, 1.43] | 1.26 [1.11, 1.43] |
|  | Hartlepool | 1.17 [0.98, 1.39] | 1.11 [0.93, 1.33] |
|  | Havering | 1.77 [1.57, 1.99] | 1.75 [1.56, 1.98] |
|  | Herefordshire, County of | 1.87 [1.64, 2.13] | 1.83 [1.61, 2.08] |
|  | Hertfordshire | 1.44 [1.30, 1.59] | 1.42 [1.28, 1.56] |
|  | Hillingdon | 1.14 [1.00, 1.29] | 1.12 [0.99, 1.27] |
|  | Hounslow | 1.05 [0.92, 1.19] | 1.05 [0.92, 1.19] |
|  | Isle of Wight | 1.39 [1.19, 1.62] | 1.38 [1.18, 1.61] |
|  | Islington | 0.85 [0.74, 0.97] | 0.86 [0.75, 0.99] |
|  | Kensington and Chelsea | 0.86 [0.73, 1.02] | 0.88 [0.74, 1.04] |
|  | Kent | 1.66 [1.51, 1.83] | 1.64 [1.49, 1.81] |
|  | Kingston upon Hull, City of | 1.96 [1.75, 2.19] | 1.86 [1.67, 2.09] |
|  | Kingston upon Thames | 1.33 [1.17, 1.53] | 1.33 [1.17, 1.53] |
|  | Kirklees | 1.49 [1.34, 1.67] | 1.45 [1.30, 1.61] |
|  | Knowsley | 1.89 [1.66, 2.15] | 1.80 [1.58, 2.04] |
|  | Lambeth | 1.01 [0.89, 1.14] | 1.02 [0.91, 1.15] |
|  | Lancashire | 1.62 [1.47, 1.78] | 1.56 [1.41, 1.72] |
|  | Leeds | 1.00 [0.90, 1.11] | 0.98 [0.88, 1.09] |
|  | Leicester | 1.05 [0.93, 1.18] | 1.01 [0.90, 1.14] |
|  | Leicestershire | 1.28 [1.16, 1.43] | 1.26 [1.13, 1.40] |
|  | Lewisham | 1.41 [1.26, 1.59] | 1.38 [1.23, 1.55] |
|  | Lincolnshire | 1.74 [1.57, 1.92] | 1.68 [1.52, 1.86] |
|  | Liverpool | 1.61 [1.45, 1.79] | 1.56 [1.40, 1.74] |
|  | Luton | 1.43 [1.26, 1.62] | 1.39 [1.23, 1.58] |
|  | Manchester | 1.20 [1.08, 1.34] | 1.18 [1.06, 1.31] |
|  | Medway | 1.89 [1.68, 2.12] | 1.86 [1.66, 2.09] |
|  | Merton | 1.29 [1.13, 1.46] | 1.28 [1.13, 1.46] |
|  | Middlesbrough | 1.53 [1.33, 1.76] | 1.45 [1.26, 1.67] |
|  | Milton Keynes | 1.53 [1.36, 1.73] | 1.50 [1.33, 1.69] |
|  | Newcastle upon Tyne | 1.06 [0.93, 1.20] | 1.04 [0.92, 1.18] |
|  | Newham | 1.20 [1.07, 1.35] | 1.17 [1.04, 1.32] |
|  | Norfolk | 1.32 [1.19, 1.46] | 1.29 [1.16, 1.43] |
|  | North East Lincolnshire | 1.51 [1.31, 1.73] | 1.48 [1.29, 1.70] |
|  | North Lincolnshire | 1.54 [1.35, 1.77] | 1.51 [1.32, 1.73] |
|  | North Northamptonshire | 1.89 [1.69, 2.11] | 1.83 [1.64, 2.05] |
|  | North Somerset | 1.47 [1.29, 1.68] | 1.41 [1.24, 1.61] |
|  | North Tyneside | 1.10 [0.95, 1.26] | 1.05 [0.92, 1.21] |
|  | North Yorkshire | 1.20 [1.08, 1.34] | 1.18 [1.06, 1.32] |
|  | Northumberland | 1.04 [0.92, 1.18] | 1.00 [0.88, 1.14] |
|  | Nottingham | 1.16 [1.03, 1.31] | 1.14 [1.02, 1.29] |
|  | Nottinghamshire | 1.48 [1.34, 1.64] | 1.43 [1.30, 1.59] |
|  | Oldham | 1.89 [1.68, 2.12] | 1.81 [1.60, 2.03] |
|  | Oxfordshire | 1.34 [1.21, 1.49] | 1.34 [1.20, 1.48] |
|  | Peterborough | 1.71 [1.51, 1.94] | 1.67 [1.47, 1.89] |
|  | Plymouth | 1.23 [1.08, 1.39] | 1.19 [1.05, 1.35] |
|  | Portsmouth | 1.71 [1.51, 1.93] | 1.67 [1.48, 1.89] |
|  | Reading | 0.94 [0.81, 1.09] | 0.94 [0.81, 1.09] |
|  | Redbridge | 1.27 [1.13, 1.43] | 1.27 [1.12, 1.43] |
|  | Redcar and Cleveland | 1.74 [1.52, 2.00] | 1.67 [1.45, 1.91] |
|  | Richmond upon Thames | 1.06 [0.92, 1.22] | 1.06 [0.92, 1.22] |
|  | Rochdale | 1.83 [1.62, 2.06] | 1.75 [1.55, 1.97] |
|  | Rotherham | 1.87 [1.67, 2.10] | 1.76 [1.57, 1.97] |
|  | Rutland | 1.24 [0.95, 1.62] | 1.23 [0.94, 1.60] |
|  | Salford | 1.16 [1.02, 1.32] | 1.10 [0.97, 1.25] |
|  | Sandwell | 1.18 [1.05, 1.34] | 1.14 [1.01, 1.29] |
|  | Sefton | 1.76 [1.57, 1.98] | 1.71 [1.52, 1.92] |
|  | Sheffield | 1.35 [1.22, 1.50] | 1.31 [1.18, 1.46] |
|  | Shropshire | 0.93 [0.82, 1.07] | 0.92 [0.81, 1.06] |
|  | Slough | 1.41 [1.23, 1.62] | 1.39 [1.21, 1.59] |
|  | Solihull | 1.12 [0.98, 1.29] | 1.10 [0.95, 1.26] |
|  | Somerset | 1.54 [1.38, 1.71] | 1.49 [1.34, 1.66] |
|  | South Gloucestershire | 1.53 [1.36, 1.72] | 1.50 [1.33, 1.68] |
|  | South Tyneside | 1.85 [1.62, 2.11] | 1.78 [1.56, 2.03] |
|  | Southampton | 1.84 [1.64, 2.06] | 1.81 [1.62, 2.03] |
|  | Southend-on-Sea | 0.95 [0.81, 1.10] | 0.93 [0.79, 1.08] |
|  | Southwark | 1.04 [0.92, 1.17] | 1.04 [0.92, 1.18] |
|  | St. Helens | 1.32 [1.15, 1.52] | 1.26 [1.10, 1.45] |
|  | Staffordshire | 1.87 [1.69, 2.06] | 1.81 [1.64, 2.00] |
|  | Stockport | 1.99 [1.78, 2.23] | 1.91 [1.71, 2.14] |
|  | Stockton-on-Tees | 1.41 [1.23, 1.61] | 1.35 [1.18, 1.54] |
|  | Stoke-on-Trent | 2.24 [2.00, 2.50] | 2.14 [1.91, 2.39] |
|  | Suffolk | 1.18 [1.06, 1.31] | 1.16 [1.04, 1.29] |
|  | Sunderland | 1.35 [1.19, 1.52] | 1.29 [1.14, 1.46] |
|  | Surrey | 2.02 [1.83, 2.22] | 2.00 [1.82, 2.20] |
|  | Sutton | 1.54 [1.36, 1.75] | 1.51 [1.33, 1.71] |
|  | Swindon | 2.41 [2.16, 2.70] | 2.36 [2.11, 2.65] |
|  | Tameside | 1.73 [1.54, 1.95] | 1.64 [1.46, 1.85] |
|  | Telford and Wrekin | 0.91 [0.78, 1.06] | 0.89 [0.76, 1.04] |
|  | Thurrock | 1.29 [1.13, 1.49] | 1.26 [1.10, 1.45] |
|  | Torbay | 2.58 [2.27, 2.94] | 2.49 [2.19, 2.83] |
|  | Tower Hamlets | 1.19 [1.06, 1.34] | 1.21 [1.08, 1.37] |
|  | Trafford | 1.60 [1.42, 1.81] | 1.55 [1.37, 1.76] |
|  | Wakefield | 1.76 [1.58, 1.97] | 1.71 [1.53, 1.92] |
|  | Walsall | 2.09 [1.87, 2.34] | 2.02 [1.81, 2.27] |
|  | Waltham Forest | 1.21 [1.07, 1.37] | 1.20 [1.06, 1.35] |
|  | Wandsworth | 1.00 [0.89, 1.13] | 1.02 [0.91, 1.15] |
|  | Warrington | 1.21 [1.06, 1.39] | 1.17 [1.03, 1.34] |
|  | Warwickshire | 1.29 [1.16, 1.43] | 1.26 [1.13, 1.41] |
|  | West Berkshire | 1.37 [1.19, 1.57] | 1.36 [1.18, 1.56] |
|  | West Northamptonshire | 1.65 [1.48, 1.84] | 1.61 [1.44, 1.79] |
|  | West Sussex | 1.82 [1.65, 2.01] | 1.79 [1.62, 1.98] |
|  | Westminster | 0.73 [0.63, 0.85] | 0.75 [0.65, 0.88] |
|  | Wigan | 1.37 [1.22, 1.54] | 1.32 [1.17, 1.48] |
|  | Wiltshire | 1.66 [1.49, 1.85] | 1.63 [1.46, 1.81] |
|  | Windsor and Maidenhead | 1.24 [1.07, 1.44] | 1.23 [1.06, 1.43] |
|  | Wirral | 2.65 [2.38, 2.95] | 2.53 [2.28, 2.82] |
|  | Wokingham | 0.94 [0.80, 1.10] | 0.94 [0.80, 1.10] |
|  | Wolverhampton | 1.50 [1.33, 1.70] | 1.46 [1.29, 1.65] |
|  | Worcestershire | 2.20 [1.99, 2.44] | 2.17 [1.96, 2.40] |
|  | York | 1.22 [1.07, 1.39] | 1.22 [1.07, 1.39] |
| Secondary analysis | Barking and Dagenham | 1.19 [1.09, 1.30] | 1.16 [1.06, 1.26] |
|  | Barnet | 0.91 [0.84, 0.99] | 0.91 [0.84, 0.98] |
|  | Barnsley | 1.43 [1.31, 1.54] | 1.36 [1.25, 1.47] |
|  | Bath and North East Somerset | 0.96 [0.87, 1.05] | 0.96 [0.87, 1.06] |
|  | Bedford | 1.08 [0.98, 1.19] | 1.07 [0.97, 1.18] |
|  | Bexley | 1.57 [1.45, 1.70] | 1.54 [1.42, 1.66] |
|  | Birmingham | 1.16 [1.08, 1.24] | 1.13 [1.05, 1.20] |
|  | Blackburn with Darwen | 1.46 [1.34, 1.60] | 1.40 [1.28, 1.53] |
|  | Blackpool | 1.50 [1.37, 1.65] | 1.43 [1.31, 1.57] |
|  | Bolton | 1.17 [1.08, 1.27] | 1.14 [1.05, 1.23] |
|  | Bournemouth, Christchurch and Poole | 1.19 [1.10, 1.28] | 1.16 [1.07, 1.25] |
|  | Bracknell Forest | 1.25 [1.13, 1.38] | 1.25 [1.13, 1.38] |
|  | Bradford | 1.35 [1.26, 1.45] | 1.28 [1.19, 1.37] |
|  | Brent | 1.05 [0.97, 1.14] | 1.06 [0.97, 1.14] |
|  | Brighton and Hove | 0.99 [0.91, 1.07] | 1.00 [0.92, 1.08] |
|  | Bristol, City of | 1.14 [1.06, 1.22] | 1.11 [1.04, 1.20] |
|  | Bromley | 1.76 [1.64, 1.89] | 1.70 [1.58, 1.83] |
|  | Buckinghamshire | 1.11 [1.03, 1.20] | 1.11 [1.04, 1.20] |
|  | Bury | 1.09 [0.99, 1.19] | 1.05 [0.96, 1.16] |
|  | Calderdale | 1.02 [0.93, 1.12] | 0.99 [0.90, 1.08] |
|  | Cambridgeshire | 1.06 [0.99, 1.14] | 1.06 [0.99, 1.14] |
|  | Camden | 1.08 [0.99, 1.18] | 1.11 [1.02, 1.21] |
|  | Central Bedfordshire | 1.26 [1.16, 1.36] | 1.24 [1.14, 1.34] |
|  | Cheshire East | 1.20 [1.12, 1.30] | 1.18 [1.09, 1.27] |
|  | Cheshire West and Chester | 1.18 [1.09, 1.27] | 1.14 [1.05, 1.23] |
|  | Cornwall and Isles of Scilly | 1.56 [1.46, 1.67] | 1.50 [1.40, 1.61] |
|  | County Durham | 1.15 [1.07, 1.24] | 1.11 [1.03, 1.19] |
|  | Coventry | 1.03 [0.96, 1.12] | 1.01 [0.93, 1.09] |
|  | Croydon | 1.27 [1.17, 1.36] | 1.23 [1.15, 1.33] |
|  | Cumbria | 1.02 [0.95, 1.10] | 1.00 [0.93, 1.08] |
|  | Darlington | 1.25 [1.12, 1.39] | 1.21 [1.09, 1.34] |
|  | Derby | 1.42 [1.31, 1.54] | 1.36 [1.25, 1.47] |
|  | Derbyshire | 1.46 [1.36, 1.56] | 1.41 [1.32, 1.50] |
|  | Devon | 1.47 [1.38, 1.57] | 1.44 [1.35, 1.54] |
|  | Doncaster | 1.11 [1.02, 1.20] | 1.08 [0.99, 1.17] |
|  | Dorset | 1.50 [1.39, 1.62] | 1.46 [1.35, 1.57] |
|  | Dudley | 1.18 [1.09, 1.28] | 1.13 [1.04, 1.22] |
|  | Ealing | 0.92 [0.85, 0.99] | 0.91 [0.84, 0.99] |
|  | East Riding of Yorkshire | 1.24 [1.15, 1.34] | 1.21 [1.12, 1.30] |
|  | East Sussex | 1.31 [1.22, 1.41] | 1.29 [1.20, 1.39] |
|  | Enfield | 1.03 [0.95, 1.12] | 1.01 [0.93, 1.10] |
|  | Essex | 1.29 [1.21, 1.37] | 1.27 [1.19, 1.35] |
|  | Gateshead | 1.09 [0.99, 1.19] | 1.07 [0.97, 1.17] |
|  | Gloucestershire | 1.25 [1.17, 1.34] | 1.23 [1.15, 1.32] |
|  | Greenwich | 1.25 [1.16, 1.36] | 1.23 [1.14, 1.33] |
|  | Halton | 1.63 [1.49, 1.79] | 1.55 [1.41, 1.70] |
|  | Hammersmith and Fulham | 0.88 [0.80, 0.97] | 0.90 [0.82, 0.99] |
|  | Hampshire | 1.35 [1.27, 1.44] | 1.34 [1.25, 1.42] |
|  | Haringey | 0.97 [0.89, 1.05] | 0.97 [0.89, 1.06] |
|  | Harrow | 1.15 [1.06, 1.25] | 1.16 [1.06, 1.26] |
|  | Hartlepool | 1.29 [1.16, 1.44] | 1.23 [1.10, 1.38] |
|  | Havering | 1.50 [1.39, 1.62] | 1.49 [1.38, 1.61] |
|  | Herefordshire, County of | 1.44 [1.32, 1.58] | 1.43 [1.31, 1.56] |
|  | Hertfordshire | 1.25 [1.18, 1.34] | 1.24 [1.16, 1.32] |
|  | Hillingdon | 1.06 [0.98, 1.15] | 1.04 [0.96, 1.13] |
|  | Hounslow | 0.94 [0.86, 1.02] | 0.94 [0.86, 1.02] |
|  | Isle of Wight | 1.52 [1.38, 1.67] | 1.52 [1.39, 1.68] |
|  | Islington | 1.09 [1.00, 1.19] | 1.11 [1.02, 1.21] |
|  | Kensington and Chelsea | 0.84 [0.75, 0.93] | 0.85 [0.76, 0.95] |
|  | Kent | 1.35 [1.27, 1.44] | 1.34 [1.26, 1.43] |
|  | Kingston upon Hull, City of | 1.34 [1.24, 1.45] | 1.28 [1.18, 1.39] |
|  | Kingston upon Thames | 1.09 [0.99, 1.19] | 1.09 [0.99, 1.19] |
|  | Kirklees | 1.20 [1.12, 1.29] | 1.17 [1.08, 1.26] |
|  | Knowsley | 2.04 [1.87, 2.21] | 1.93 [1.78, 2.10] |
|  | Lambeth | 0.96 [0.89, 1.04] | 0.97 [0.90, 1.05] |
|  | Lancashire | 1.45 [1.36, 1.54] | 1.39 [1.31, 1.49] |
|  | Leeds | 0.89 [0.83, 0.96] | 0.87 [0.82, 0.94] |
|  | Leicester | 1.08 [1.00, 1.17] | 1.04 [0.96, 1.13] |
|  | Leicestershire | 1.28 [1.19, 1.37] | 1.25 [1.17, 1.34] |
|  | Lewisham | 1.25 [1.16, 1.35] | 1.22 [1.13, 1.32] |
|  | Lincolnshire | 1.41 [1.32, 1.51] | 1.37 [1.28, 1.47] |
|  | Liverpool | 1.68 [1.57, 1.80] | 1.63 [1.52, 1.75] |
|  | Luton | 1.29 [1.19, 1.41] | 1.26 [1.16, 1.38] |
|  | Manchester | 1.10 [1.02, 1.18] | 1.07 [0.99, 1.15] |
|  | Medway | 1.94 [1.80, 2.09] | 1.92 [1.78, 2.07] |
|  | Merton | 1.02 [0.93, 1.11] | 1.02 [0.93, 1.11] |
|  | Middlesbrough | 1.09 [0.99, 1.21] | 1.04 [0.94, 1.15] |
|  | Milton Keynes | 1.37 [1.27, 1.49] | 1.35 [1.24, 1.46] |
|  | Newcastle upon Tyne | 1.09 [1.00, 1.18] | 1.07 [0.98, 1.16] |
|  | Newham | 1.14 [1.05, 1.23] | 1.11 [1.03, 1.20] |
|  | Norfolk | 1.23 [1.15, 1.32] | 1.21 [1.13, 1.29] |
|  | North East Lincolnshire | 1.16 [1.06, 1.28] | 1.15 [1.05, 1.27] |
|  | North Lincolnshire | 1.08 [0.98, 1.19] | 1.06 [0.96, 1.17] |
|  | North Northamptonshire | 1.47 [1.36, 1.58] | 1.43 [1.33, 1.54] |
|  | North Somerset | 1.40 [1.29, 1.53] | 1.35 [1.24, 1.47] |
|  | North Tyneside | 1.05 [0.96, 1.15] | 1.01 [0.92, 1.11] |
|  | North Yorkshire | 1.03 [0.95, 1.10] | 1.01 [0.94, 1.09] |
|  | Northumberland | 0.96 [0.88, 1.04] | 0.92 [0.85, 1.00] |
|  | Nottingham | 1.30 [1.21, 1.41] | 1.28 [1.18, 1.38] |
|  | Nottinghamshire | 1.43 [1.34, 1.53] | 1.39 [1.30, 1.48] |
|  | Oldham | 1.53 [1.41, 1.66] | 1.47 [1.35, 1.59] |
|  | Oxfordshire | 1.17 [1.09, 1.26] | 1.17 [1.09, 1.25] |
|  | Peterborough | 1.28 [1.17, 1.39] | 1.25 [1.15, 1.37] |
|  | Plymouth | 1.24 [1.15, 1.35] | 1.21 [1.11, 1.31] |
|  | Portsmouth | 1.28 [1.18, 1.40] | 1.26 [1.16, 1.37] |
|  | Reading | 0.88 [0.80, 0.97] | 0.89 [0.80, 0.98] |
|  | Redbridge | 1.16 [1.07, 1.26] | 1.16 [1.07, 1.25] |
|  | Redcar and Cleveland | 1.26 [1.14, 1.39] | 1.21 [1.10, 1.34] |
|  | Richmond upon Thames | 0.88 [0.80, 0.97] | 0.88 [0.80, 0.97] |
|  | Rochdale | 1.40 [1.29, 1.52] | 1.34 [1.24, 1.46] |
|  | Rotherham | 1.58 [1.46, 1.71] | 1.49 [1.38, 1.61] |
|  | Rutland | 1.08 [0.90, 1.29] | 1.08 [0.90, 1.29] |
|  | Salford | 1.12 [1.03, 1.22] | 1.06 [0.98, 1.16] |
|  | Sandwell | 1.05 [0.97, 1.14] | 1.01 [0.94, 1.10] |
|  | Sefton | 1.73 [1.60, 1.87] | 1.68 [1.55, 1.81] |
|  | Sheffield | 1.06 [0.99, 1.14] | 1.03 [0.96, 1.11] |
|  | Shropshire | 1.12 [1.03, 1.21] | 1.11 [1.02, 1.20] |
|  | Slough | 1.13 [1.03, 1.25] | 1.12 [1.01, 1.23] |
|  | Solihull | 1.04 [0.95, 1.13] | 1.01 [0.93, 1.11] |
|  | Somerset | 1.23 [1.15, 1.33] | 1.20 [1.12, 1.29] |
|  | South Gloucestershire | 1.46 [1.35, 1.58] | 1.43 [1.33, 1.55] |
|  | South Tyneside | 1.55 [1.42, 1.70] | 1.49 [1.37, 1.63] |
|  | Southampton | 1.55 [1.43, 1.67] | 1.53 [1.41, 1.65] |
|  | Southend-on-Sea | 0.97 [0.88, 1.07] | 0.95 [0.86, 1.05] |
|  | Southwark | 1.02 [0.94, 1.10] | 1.02 [0.94, 1.10] |
|  | St. Helens | 1.56 [1.43, 1.69] | 1.49 [1.37, 1.62] |
|  | Staffordshire | 1.52 [1.42, 1.62] | 1.48 [1.39, 1.58] |
|  | Stockport | 1.57 [1.46, 1.69] | 1.51 [1.40, 1.63] |
|  | Stockton-on-Tees | 1.36 [1.24, 1.48] | 1.30 [1.19, 1.42] |
|  | Stoke-on-Trent | 1.75 [1.62, 1.89] | 1.68 [1.56, 1.81] |
|  | Suffolk | 1.09 [1.02, 1.17] | 1.08 [1.00, 1.15] |
|  | Sunderland | 1.22 [1.13, 1.32] | 1.17 [1.08, 1.27] |
|  | Surrey | 1.42 [1.33, 1.51] | 1.41 [1.32, 1.51] |
|  | Sutton | 1.25 [1.15, 1.36] | 1.23 [1.13, 1.34] |
|  | Swindon | 1.84 [1.71, 1.99] | 1.81 [1.67, 1.96] |
|  | Tameside | 1.53 [1.41, 1.66] | 1.45 [1.34, 1.57] |
|  | Telford and Wrekin | 1.19 [1.08, 1.30] | 1.17 [1.06, 1.28] |
|  | Thurrock | 1.21 [1.10, 1.32] | 1.18 [1.08, 1.30] |
|  | Torbay | 1.91 [1.75, 2.09] | 1.85 [1.69, 2.03] |
|  | Tower Hamlets | 1.11 [1.03, 1.20] | 1.13 [1.04, 1.23] |
|  | Trafford | 1.31 [1.20, 1.42] | 1.27 [1.17, 1.38] |
|  | Wakefield | 1.53 [1.42, 1.65] | 1.50 [1.39, 1.61] |
|  | Walsall | 1.62 [1.50, 1.75] | 1.57 [1.46, 1.70] |
|  | Waltham Forest | 1.14 [1.05, 1.24] | 1.13 [1.04, 1.22] |
|  | Wandsworth | 0.89 [0.82, 0.97] | 0.91 [0.84, 0.99] |
|  | Warrington | 1.40 [1.28, 1.52] | 1.36 [1.25, 1.47] |
|  | Warwickshire | 1.26 [1.18, 1.35] | 1.24 [1.16, 1.33] |
|  | West Berkshire | 1.10 [1.00, 1.21] | 1.10 [1.00, 1.21] |
|  | West Northamptonshire | 1.48 [1.38, 1.59] | 1.45 [1.35, 1.56] |
|  | West Sussex | 1.38 [1.29, 1.48] | 1.37 [1.28, 1.46] |
|  | Westminster | 0.81 [0.74, 0.89] | 0.83 [0.75, 0.91] |
|  | Wigan | 1.19 [1.10, 1.29] | 1.15 [1.06, 1.24] |
|  | Wiltshire | 1.35 [1.25, 1.45] | 1.33 [1.23, 1.42] |
|  | Windsor and Maidenhead | 0.98 [0.88, 1.08] | 0.98 [0.88, 1.08] |
|  | Wirral | 1.98 [1.84, 2.13] | 1.90 [1.77, 2.04] |
|  | Wokingham | 0.86 [0.78, 0.96] | 0.87 [0.78, 0.96] |
|  | Wolverhampton | 1.51 [1.40, 1.64] | 1.47 [1.36, 1.59] |
|  | Worcestershire | 1.61 [1.50, 1.72] | 1.59 [1.48, 1.70] |
|  | York | 0.98 [0.89, 1.07] | 0.98 [0.89, 1.07] |

Notes: Reference category City of London and Hackney

**Supplementary Table 11: Secondary analysis – ICD-10 primary diagnosis type**

| **ICD-10 primary diagnosis** | **Count (%)** |
| --- | --- |
| Total | 262,065 (100.0%) |
| N80 Endometriosis | 116,835 (44.6%) |
| D25 Leiomyoma of uterus | 21,845 (8.3%) |
| R10 Abdominal and pelvic pain | 11,135 (4.2%) |
| N83 Noninflammatory disorders of ovary fallopian tube and broad ligament | 10,835 (4.1%) |
| N92 Excessive frequent and irregular menstruation | 8,890 (3.4%) |
| N81 Female genital prolapse | 7,465 (2.8%) |
| D27 Benign neoplasm of ovary | 5,660 (2.2%) |
| N73 Other female pelvic inflammatory diseases | 4,200 (1.6%) |
| C54 Malignant neoplasm of corpus uteri | 4,185 (1.6%) |
| N84 Polyp of female genital tract | 2,745 (1.0%) |
| Other primary diagnosis | 68,275 (26.1%) |

**Supplementary Table 12: Sensitivity analysis – Odds ratios for endometriosis diagnosis and 95% confidence intervals by country of birth, main language, IMD decile group, household NS-SEC, highest level of qualification, general health, disability and rural/urban classification**

| **Analysis type** | **Exposure** | **Term** | **Adjusted for age,**  **OR [95% CI]** | **Adjusted for age and health,**  **OR [95% CI]** |
| --- | --- | --- | --- | --- |
| Main sensitivity analysis | Country of birth (Reference category: Born in the UK) | Born outside the UK | 0.68 [0.65, 0.70] | 0.69 [0.67, 0.71] |
|  | Main language (Reference category: Main language is English) | Main language is not English | 0.64 [0.61, 0.67] | 0.65 [0.63, 0.68] |
|  | IMD decile group (Reference category: 10 (least deprived)) | 1 (most deprived) | 0.99 [0.94, 1.04] | 0.93 [0.89, 0.98] |
|  |  | 2 | 1.02 [0.97, 1.07] | 0.98 [0.93, 1.03] |
|  |  | 3 | 1.06 [1.01, 1.11] | 1.03 [0.98, 1.08] |
|  |  | 4 | 1.05 [1.00, 1.11] | 1.03 [0.98, 1.09] |
|  |  | 5 | 1.07 [1.02, 1.12] | 1.05 [1.00, 1.11] |
|  |  | 6 | 1.04 [0.99, 1.09] | 1.03 [0.98, 1.08] |
|  |  | 7 | 1.08 [1.03, 1.14] | 1.07 [1.02, 1.13] |
|  |  | 8 | 1.08 [1.03, 1.14] | 1.08 [1.02, 1.13] |
|  |  | 9 | 1.06 [1.01, 1.12] | 1.06 [1.01, 1.11] |
|  | Household NS-SEC (Reference category: Class 1: Higher managerial, administrative and professional occupations) | Class 2: Lower managerial, administrative and professional occupations | 1.19 [1.14, 1.23] | 1.18 [1.13, 1.22] |
|  |  | Class 3: Intermediate occupations | 1.29 [1.23, 1.35] | 1.27 [1.21, 1.32] |
|  |  | Class 4: Small employers and own account workers | 1.17 [1.12, 1.23] | 1.15 [1.10, 1.20] |
|  |  | Class 5: Lower supervisory and technical occupations | 1.32 [1.26, 1.38] | 1.27 [1.22, 1.34] |
|  |  | Class 6: Semi-routine occupations | 1.23 [1.18, 1.28] | 1.18 [1.13, 1.23] |
|  |  | Class 7: Routine occupations | 1.15 [1.10, 1.20] | 1.10 [1.05, 1.15] |
|  |  | Class 8: Never worked and long-term unemployed | 0.88 [0.82, 0.94] | 0.81 [0.76, 0.87] |
|  |  | Students | 0.80 [0.73, 0.87] | 0.82 [0.75, 0.90] |
|  |  | Not classified | 0.48 [0.41, 0.56] | 0.49 [0.42, 0.58] |
|  | Highest level of qualification (Reference category: Level 4 and above) | No academic or professional qualifications | 0.92 [0.88, 0.96] | 0.85 [0.81, 0.88] |
|  |  | Level 1 | 1.24 [1.20, 1.28] | 1.18 [1.14, 1.22] |
|  |  | Level 2 | 1.25 [1.21, 1.29] | 1.21 [1.17, 1.25] |
|  |  | Apprenticeship | 1.46 [1.31, 1.62] | 1.39 [1.24, 1.55] |
|  |  | Level 3 | 1.20 [1.16, 1.24] | 1.18 [1.14, 1.22] |
|  |  | Other qualifications | 0.83 [0.78, 0.88] | 0.82 [0.77, 0.87] |
|  |  | Not classified | 0.83 [0.69, 1.00] | 0.79 [0.66, 0.96] |
|  | Highest level of qualification, 25 years and over (Reference category: Level 4 and above) | No academic or professional qualifications | 0.99 [0.97, 1.02] | 0.95 [0.92, 0.97] |
|  |  | Level 1 | 1.28 [1.26, 1.31] | 1.25 [1.22, 1.28] |
|  |  | Level 2 | 1.31 [1.29, 1.34] | 1.28 [1.26, 1.31] |
|  |  | Apprenticeship | 1.35 [1.24, 1.47] | 1.32 [1.21, 1.43] |
|  |  | Level 3 | 1.31 [1.28, 1.33] | 1.29 [1.26, 1.31] |
|  |  | Other qualifications | 0.92 [0.89, 0.96] | 0.92 [0.88, 0.95] |
|  | General health (Reference category: Very good health) | Good health | 1.59 [1.55, 1.63] | x |
|  |  | Fair health | 2.20 [2.13, 2.28] | x |
|  |  | Bad health | 2.30 [2.17, 2.44] | x |
|  |  | Very bad health | 1.63 [1.43, 1.85] | x |
|  | Disability (Reference category: Day-to-day activities not limited) | Day-to-day activities limited a little | 1.72 [1.65, 1.79] | x |
|  |  | Day-to-day activities limited a lot | 1.32 [1.25, 1.39] | x |
|  | Rural/urban classification (Reference category: Urban) | Rural | 1.05 [1.02, 1.08] | 1.05 [1.02, 1.08] |
| Secondary sensitivity analysis | Country of birth (Reference category: Born in the UK) | Born outside the UK | 0.70 [0.68, 0.71] | 0.71 [0.70, 0.73] |
|  | Main language (Reference category: Main language is English) | Main language is not English | 0.65 [0.63, 0.66] | 0.66 [0.64, 0.68] |
|  | IMD decile group (Reference category: 10 (least deprived)) | 1 (most deprived) | 1.13 [1.10, 1.17] | 1.05 [1.02, 1.09] |
|  |  | 2 | 1.13 [1.10, 1.17] | 1.08 [1.04, 1.12] |
|  |  | 3 | 1.14 [1.10, 1.18] | 1.10 [1.06, 1.14] |
|  |  | 4 | 1.13 [1.09, 1.17] | 1.10 [1.06, 1.14] |
|  |  | 5 | 1.15 [1.11, 1.19] | 1.13 [1.09, 1.17] |
|  |  | 6 | 1.10 [1.06, 1.14] | 1.08 [1.05, 1.12] |
|  |  | 7 | 1.13 [1.09, 1.17] | 1.11 [1.08, 1.15] |
|  |  | 8 | 1.10 [1.06, 1.14] | 1.09 [1.05, 1.13] |
|  |  | 9 | 1.09 [1.05, 1.13] | 1.08 [1.04, 1.12] |
|  | Household NS-SEC (Reference category: Class 1: Higher managerial, administrative and professional occupations) | Class 2: Lower managerial, administrative and professional occupations | 1.19 [1.16, 1.22] | 1.18 [1.15, 1.21] |
|  |  | Class 3: Intermediate occupations | 1.29 [1.25, 1.32] | 1.26 [1.22, 1.30] |
|  |  | Class 4: Small employers and own account workers | 1.18 [1.15, 1.22] | 1.16 [1.12, 1.19] |
|  |  | Class 5: Lower supervisory and technical occupations | 1.31 [1.27, 1.35] | 1.26 [1.22, 1.30] |
|  |  | Class 6: Semi-routine occupations | 1.26 [1.23, 1.30] | 1.20 [1.17, 1.24] |
|  |  | Class 7: Routine occupations | 1.20 [1.16, 1.23] | 1.14 [1.10, 1.17] |
|  |  | Class 8: Never worked and long-term unemployed | 0.95 [0.91, 1.00] | 0.86 [0.82, 0.90] |
|  |  | Students | 0.83 [0.78, 0.89] | 0.86 [0.80, 0.91] |
|  |  | Not classified | 0.50 [0.45, 0.57] | 0.51 [0.45, 0.57] |
|  | Highest level of qualification (Reference category: Level 4 and above) | No academic or professional qualifications | 0.99 [0.97, 1.02] | 0.91 [0.89, 0.94] |
|  |  | Level 1 | 1.21 [1.18, 1.23] | 1.15 [1.12, 1.17] |
|  |  | Level 2 | 1.24 [1.22, 1.27] | 1.20 [1.17, 1.23] |
|  |  | Apprenticeship | 1.40 [1.30, 1.51] | 1.33 [1.24, 1.44] |
|  |  | Level 3 | 1.19 [1.16, 1.22] | 1.17 [1.15, 1.20] |
|  |  | Other qualifications | 0.85 [0.82, 0.89] | 0.84 [0.80, 0.87] |
|  |  | Not classified | 0.89 [0.77, 1.04] | 0.86 [0.74, 1.00] |
|  | Highest level of qualification, 25 years and over (Reference category: Level 4 and above) | No academic or professional qualifications | 1.00 [0.99, 1.02] | 0.96 [0.94, 0.97] |
|  |  | Level 1 | 1.20 [1.19, 1.22] | 1.17 [1.16, 1.19] |
|  |  | Level 2 | 1.23 [1.21, 1.25] | 1.21 [1.19, 1.22] |
|  |  | Apprenticeship | 1.24 [1.17, 1.30] | 1.21 [1.15, 1.28] |
|  |  | Level 3 | 1.23 [1.22, 1.25] | 1.22 [1.20, 1.23] |
|  |  | Other qualifications | 0.91 [0.89, 0.93] | 0.90 [0.88, 0.92] |
|  | General health (Reference category: Very good health) | Good health | 1.65 [1.62, 1.68] | x |
|  |  | Fair health | 2.42 [2.36, 2.47] | x |
|  |  | Bad health | 2.62 [2.53, 2.72] | x |
|  |  | Very bad health | 2.23 [2.08, 2.39] | x |
|  | Disability (Reference category: Day-to-day activities not limited) | Day-to-day activities limited a little | 1.81 [1.77, 1.86] | x |
|  |  | Day-to-day activities limited a lot | 1.52 [1.48, 1.57] | x |
|  | Rural/urban classification (Reference category: Urban) | Rural | 0.99 [0.97, 1.01] | 1.00 [0.98, 1.02] |

Notes: Values x are not applicable for the given exposure. For household [NS-SEC](https://www.ons.gov.uk/methodology/classificationsandstandards/otherclassifications/thenationalstatisticssocioeconomicclassificationnssecrebasedonsoc2010), “Not classified” includes those not living in a private household in Census 2011, occupations not stated or inadequately described, or not classifiable for other reasons. For highest level of qualification, “Not classified” includes those aged under 16 in Census 2011.

**Supplementary Table 13: Sensitivity analysis – Odds ratios for endometriosis diagnosis and 95% confidence intervals by age on Census Day (five-year bands)**

| **Analysis type** | **Term** | **Adjusted for health,**  **OR [95% CI]** |
| --- | --- | --- |
| Main sensitivity analysis | 0 to 9 years | c |
|  | 10 to 14 years | c |
|  | 15 to 19 years | 0.38 [0.36, 0.40] |
|  | 20 to 24 years | 0.78 [0.75, 0.81] |
|  | 25 to 29 years | 0.86 [0.83, 0.89] |
|  | 30 to 34 years | 0.87 [0.84, 0.90] |
|  | 40 to 44 years | 0.89 [0.86, 0.92] |
|  | 45 to 49 years | 0.50 [0.48, 0.52] |
|  | 50 to 54 years | 0.14 [0.13, 0.15] |
|  | 55 to 59 years | 0.04 [0.04, 0.05] |
|  | 60 to 64 years | 0.02 [0.02, 0.03] |
|  | 65 to 69 years | 0.02 [0.01, 0.02] |
|  | 70 to 74 years | 0.02 [0.01, 0.02] |
|  | 75 to 79 years | 0.01 [0.01, 0.01] |
|  | 80 years and over | 0.01 [0.00, 0.01] |
| Secondary sensitivity analysis | 0 to 9 years | c |
|  | 10 to 14 years | c |
|  | 15 to 19 years | 0.29 [0.27, 0.30] |
|  | 20 to 24 years | 0.64 [0.62, 0.66] |
|  | 25 to 29 years | 0.77 [0.75, 0.80] |
|  | 30 to 34 years | 0.85 [0.82, 0.87] |
|  | 40 to 44 years | 1.02 [0.99, 1.04] |
|  | 45 to 49 years | 0.72 [0.70, 0.74] |
|  | 50 to 54 years | 0.31 [0.29, 0.32] |
|  | 55 to 59 years | 0.16 [0.15, 0.16] |
|  | 60 to 64 years | 0.12 [0.11, 0.13] |
|  | 65 to 69 years | 0.10 [0.10, 0.11] |
|  | 70 to 74 years | 0.07 [0.07, 0.08] |
|  | 75 to 79 years | 0.05 [0.04, 0.05] |
|  | 80 years and over | 0.02 [0.02, 0.02] |

Note: Reference category: 35 to 39 years. Values c have been suppressed for the purposes of statistical disclosure control.

**Supplementary Table 14: Sensitivity analysis – Odds ratios for endometriosis diagnosis and 95% confidence intervals by ethnic group**

| **Analysis type** | **Exposure** | **Term** | **Adjusted for age,**  **OR [95% CI]** | **Adjusted for age and health,**  **OR [95% CI]** | **Adjusted for age, health and country of birth,**  **OR [95% CI]** | **Adjusted for age, health and main language,**  **OR [95% CI]** | **Adjusted for age, health, country of birth and main language,**  **OR [95% CI]** |
| --- | --- | --- | --- | --- | --- | --- | --- |
| Main sensitivity analysis | Ethnic group (detailed) (Reference category: White: English/Welsh/Scottish/Northern Irish/British) | White: Irish | 0.82 [0.72, 0.93] | 0.84 [0.74, 0.95] | 0.84 [0.78, 0.90] | 0.79 [0.74, 0.85] | 0.82 [0.76, 0.89] |
|  |  | White: Gypsy or Irish Traveller | 1.03 [0.75, 1.41] | 0.96 [0.70, 1.33] | 0.94 [0.80, 1.11] | 0.95 [0.80, 1.12] | 0.95 [0.81, 1.12] |
|  |  | White: Other White | 0.64 [0.61, 0.67] | 0.67 [0.64, 0.71] | 0.73 [0.70, 0.75] | 0.73 [0.70, 0.75] | 0.76 [0.73, 0.79] |
|  |  | Mixed/multiple: White and Black Caribbean | 0.96 [0.84, 1.09] | 0.93 [0.82, 1.06] | 0.92 [0.86, 0.98] | 0.92 [0.86, 0.98] | 0.92 [0.86, 0.98] |
|  |  | Mixed/multiple: White and Black African | 0.70 [0.54, 0.90] | 0.70 [0.54, 0.90] | 0.77 [0.68, 0.87] | 0.76 [0.67, 0.85] | 0.78 [0.69, 0.88] |
|  |  | Mixed/multiple: White and Asian | 1.03 [0.89, 1.19] | 1.06 [0.91, 1.22] | 0.88 [0.82, 0.95] | 0.88 [0.81, 0.95] | 0.89 [0.82, 0.96] |
|  |  | Mixed/multiple: Other Mixed | 0.95 [0.81, 1.10] | 0.96 [0.83, 1.12] | 0.95 [0.88, 1.02] | 0.93 [0.86, 1.01] | 0.95 [0.88, 1.03] |
|  |  | Asian: Indian | 0.65 [0.61, 0.70] | 0.67 [0.62, 0.72] | 0.75 [0.72, 0.78] | 0.74 [0.72, 0.77] | 0.76 [0.73, 0.79] |
|  |  | Asian: Pakistani | 0.64 [0.59, 0.70] | 0.62 [0.57, 0.68] | 0.72 [0.69, 0.75] | 0.73 [0.70, 0.76] | 0.74 [0.71, 0.77] |
|  |  | Asian: Bangladeshi | 0.76 [0.67, 0.86] | 0.75 [0.66, 0.85] | 0.87 [0.82, 0.92] | 0.89 [0.83, 0.94] | 0.91 [0.85, 0.96] |
|  |  | Asian: Chinese | 0.42 [0.36, 0.50] | 0.46 [0.38, 0.54] | 0.44 [0.40, 0.48] | 0.44 [0.40, 0.48] | 0.45 [0.41, 0.50] |
|  |  | Asian: Other Asian | 0.79 [0.73, 0.86] | 0.82 [0.75, 0.89] | 0.89 [0.85, 0.94] | 0.89 [0.85, 0.94] | 0.93 [0.88, 0.98] |
|  |  | Black: African | 0.44 [0.40, 0.49] | 0.43 [0.39, 0.48] | 0.47 [0.45, 0.50] | 0.45 [0.43, 0.48] | 0.48 [0.45, 0.50] |
|  |  | Black: Caribbean | 0.78 [0.70, 0.86] | 0.77 [0.69, 0.85] | 0.85 [0.80, 0.90] | 0.83 [0.78, 0.88] | 0.84 [0.80, 0.89] |
|  |  | Black: Other Black | 0.64 [0.52, 0.79] | 0.63 [0.51, 0.77] | 0.67 [0.60, 0.74] | 0.67 [0.60, 0.74] | 0.68 [0.61, 0.75] |
|  |  | Other: Arab | 0.41 [0.31, 0.55] | 0.41 [0.31, 0.54] | 0.49 [0.43, 0.56] | 0.50 [0.43, 0.57] | 0.52 [0.45, 0.59] |
|  |  | Other: Any other ethnic group | 0.92 [0.80, 1.06] | 0.92 [0.80, 1.06] | 0.90 [0.83, 0.97] | 0.90 [0.83, 0.98] | 0.93 [0.86, 1.01] |
|  | Ethnic group (aggregated) (Reference category: White) | Mixed/Multiple ethnic groups | 0.97 [0.90, 1.05] | 0.97 [0.90, 1.05] | 0.94 [0.90, 0.97] | 0.92 [0.88, 0.95] | 0.93 [0.90, 0.97] |
|  |  | Asian/Asian British | 0.69 [0.66, 0.72] | 0.70 [0.67, 0.73] | 0.83 [0.82, 0.85] | 0.82 [0.80, 0.84] | 0.85 [0.83, 0.87] |
|  |  | Black/African/Caribbean/Black British | 0.59 [0.55, 0.63] | 0.58 [0.54, 0.62] | 0.68 [0.66, 0.71] | 0.62 [0.60, 0.65] | 0.67 [0.65, 0.70] |
|  |  | Other ethnic group | 0.77 [0.68, 0.87] | 0.76 [0.67, 0.86] | 0.85 [0.80, 0.91] | 0.84 [0.78, 0.90] | 0.88 [0.82, 0.94] |
| Secondary sensitivity analysis | Ethnic group (detailed) (Reference category: White: English/Welsh/Scottish/Northern Irish/British) | White: Irish | 0.82 [0.76, 0.89] | 0.84 [0.77, 0.91] | 0.88 [0.84, 0.93] | 0.82 [0.79, 0.86] | 0.86 [0.82, 0.90] |
|  |  | White: Gypsy or Irish Traveller | 1.06 [0.85, 1.32] | 0.99 [0.79, 1.23] | 0.95 [0.85, 1.07] | 0.95 [0.85, 1.07] | 0.96 [0.85, 1.08] |
|  |  | White: Other White | 0.62 [0.60, 0.65] | 0.66 [0.63, 0.68] | 0.74 [0.72, 0.76] | 0.74 [0.73, 0.76] | 0.78 [0.76, 0.81] |
|  |  | Mixed/multiple: White and Black Caribbean | 1.03 [0.94, 1.13] | 1.00 [0.91, 1.09] | 1.03 [0.99, 1.08] | 1.03 [0.98, 1.07] | 1.03 [0.99, 1.08] |
|  |  | Mixed/multiple: White and Black African | 0.77 [0.65, 0.91] | 0.77 [0.65, 0.91] | 0.89 [0.82, 0.97] | 0.87 [0.80, 0.95] | 0.90 [0.83, 0.98] |
|  |  | Mixed/multiple: White and Asian | 0.92 [0.83, 1.03] | 0.95 [0.85, 1.06] | 0.91 [0.86, 0.96] | 0.90 [0.85, 0.95] | 0.91 [0.86, 0.97] |
|  |  | Mixed/multiple: Other Mixed | 0.96 [0.86, 1.07] | 0.98 [0.88, 1.09] | 1.00 [0.95, 1.06] | 0.99 [0.94, 1.04] | 1.01 [0.96, 1.07] |
|  |  | Asian: Indian | 0.71 [0.67, 0.74] | 0.73 [0.70, 0.77] | 0.82 [0.80, 0.84] | 0.82 [0.80, 0.84] | 0.84 [0.82, 0.87] |
|  |  | Asian: Pakistani | 0.74 [0.70, 0.79] | 0.71 [0.67, 0.75] | 0.81 [0.78, 0.83] | 0.82 [0.79, 0.84] | 0.84 [0.82, 0.87] |
|  |  | Asian: Bangladeshi | 0.78 [0.71, 0.85] | 0.76 [0.70, 0.83] | 0.90 [0.86, 0.94] | 0.92 [0.88, 0.97] | 0.95 [0.91, 1.00] |
|  |  | Asian: Chinese | 0.48 [0.43, 0.53] | 0.52 [0.46, 0.58] | 0.50 [0.47, 0.54] | 0.50 [0.47, 0.54] | 0.53 [0.50, 0.56] |
|  |  | Asian: Other Asian | 0.82 [0.77, 0.87] | 0.84 [0.80, 0.89] | 0.92 [0.89, 0.96] | 0.93 [0.90, 0.96] | 0.98 [0.94, 1.01] |
|  |  | Black: African | 0.60 [0.57, 0.64] | 0.59 [0.55, 0.63] | 0.68 [0.66, 0.71] | 0.65 [0.63, 0.68] | 0.69 [0.67, 0.72] |
|  |  | Black: Caribbean | 1.03 [0.97, 1.10] | 1.01 [0.95, 1.07] | 1.07 [1.03, 1.10] | 1.03 [1.00, 1.07] | 1.05 [1.02, 1.09] |
|  |  | Black: Other Black | 0.94 [0.83, 1.05] | 0.91 [0.81, 1.02] | 0.92 [0.87, 0.98] | 0.92 [0.86, 0.98] | 0.94 [0.88, 1.00] |
|  |  | Other: Arab | 0.53 [0.45, 0.63] | 0.52 [0.44, 0.62] | 0.58 [0.53, 0.63] | 0.59 [0.54, 0.64] | 0.62 [0.56, 0.68] |
|  |  | Other: Any other ethnic group | 0.84 [0.76, 0.93] | 0.84 [0.76, 0.93] | 0.94 [0.89, 0.99] | 0.94 [0.89, 1.00] | 0.98 [0.93, 1.04] |
|  | Ethnic group (aggregated)  (Reference category: White) | Mixed/Multiple ethnic groups | 0.98 [0.93, 1.04] | 0.98 [0.92, 1.03] | 1.02 [0.99, 1.05] | 0.99 [0.97, 1.02] | 1.01 [0.99, 1.04] |
|  |  | Asian/Asian British | 0.75 [0.73, 0.77] | 0.76 [0.73, 0.78] | 0.90 [0.89, 0.92] | 0.89 [0.87, 0.90] | 0.93 [0.91, 0.95] |
|  |  | Black/African/Caribbean/Black British | 0.81 [0.77, 0.84] | 0.78 [0.75, 0.82] | 0.92 [0.90, 0.95] | 0.84 [0.82, 0.86] | 0.91 [0.89, 0.93] |
|  |  | Other ethnic group | 0.76 [0.69, 0.83] | 0.75 [0.69, 0.82] | 0.91 [0.87, 0.96] | 0.90 [0.86, 0.94] | 0.95 [0.91, 1.00] |

**Supplementary Table 15: Sensitivity analysis – Odds ratios for endometriosis diagnosis and 95% confidence intervals by region**

| **Analysis type** | **Term** | **Adjusted for age,**  **OR [95% CI]** | **Adjusted for age and health,**  **OR [95% CI]** | **Adjusted for age, health and IMD decile,**  **OR [95% CI]** | **Adjusted for age, health, IMD decile and highest level of qualification,**  **OR [95% CI]** |
| --- | --- | --- | --- | --- | --- |
| Main sensitivity analysis | North East | 1.03 [0.97, 1.10] | 0.99 [0.94, 1.06] | 1.07 [1.04, 1.10] | 1.02 [0.99, 1.06] |
|  | North West | 1.20 [1.15, 1.25] | 1.16 [1.11, 1.20] | 1.27 [1.24, 1.30] | 1.22 [1.20, 1.25] |
|  | Yorkshire and the Humber | 1.18 [1.13, 1.24] | 1.14 [1.09, 1.20] | 1.16 [1.13, 1.19] | 1.12 [1.09, 1.15] |
|  | East Midlands | 1.22 [1.16, 1.27] | 1.18 [1.13, 1.24] | 1.24 [1.21, 1.27] | 1.19 [1.16, 1.22] |
|  | West Midlands | 1.22 [1.17, 1.28] | 1.19 [1.14, 1.25] | 1.23 [1.20, 1.26] | 1.19 [1.16, 1.21] |
|  | East of England | 1.14 [1.09, 1.19] | 1.12 [1.07, 1.17] | 1.16 [1.13, 1.18] | 1.11 [1.08, 1.13] |
|  | South East | 1.42 [1.37, 1.48] | 1.41 [1.36, 1.47] | 1.41 [1.39, 1.44] | 1.36 [1.34, 1.39] |
|  | South West | 1.42 [1.36, 1.48] | 1.39 [1.33, 1.45] | 1.34 [1.30, 1.37] | 1.28 [1.25, 1.31] |
| Secondary sensitivity analysis | North East | 1.07 [1.03, 1.11] | 1.03 [0.98, 1.07] | 1.04 [1.01, 1.06] | 1.00 [0.98, 1.02] |
|  | North West | 1.28 [1.25, 1.32] | 1.23 [1.20, 1.27] | 1.24 [1.22, 1.26] | 1.20 [1.19, 1.22] |
|  | Yorkshire and the Humber | 1.12 [1.09, 1.16] | 1.09 [1.05, 1.12] | 1.05 [1.03, 1.07] | 1.02 [1.00, 1.03] |
|  | East Midlands | 1.28 [1.24, 1.32] | 1.24 [1.21, 1.28] | 1.24 [1.22, 1.26] | 1.20 [1.18, 1.22] |
|  | West Midlands | 1.27 [1.23, 1.31] | 1.24 [1.20, 1.27] | 1.19 [1.17, 1.21] | 1.15 [1.14, 1.17] |
|  | East of England | 1.10 [1.07, 1.13] | 1.09 [1.06, 1.12] | 1.10 [1.08, 1.12] | 1.07 [1.05, 1.08] |
|  | South East | 1.23 [1.20, 1.27] | 1.23 [1.20, 1.26] | 1.21 [1.20, 1.23] | 1.18 [1.16, 1.20] |
|  | South West | 1.41 [1.37, 1.45] | 1.38 [1.34, 1.42] | 1.23 [1.21, 1.25] | 1.19 [1.17, 1.21] |

Notes: Reference category: London

**Supplementary Table 16: Sensitivity analysis – Odds ratios for endometriosis diagnosis and 95% confidence intervals by upper tier local authority (UTLA)**

| **Analysis type** | **Term** | **Adjusted for age,**  **OR [95% CI]** | **Adjusted for age and health,**  **OR [95% CI]** |
| --- | --- | --- | --- |
| Main sensitivity analysis | Barking and Dagenham | 1.05 [0.81, 1.37] | 1.02 [0.78, 1.33] |
|  | Barnet | 0.83 [0.65, 1.05] | 0.82 [0.64, 1.04] |
|  | Barnsley | 1.61 [1.28, 2.02] | 1.52 [1.21, 1.91] |
|  | Bath and North East Somerset | 1.03 [0.78, 1.36] | 1.03 [0.78, 1.36] |
|  | Bedford | 1.04 [0.78, 1.39] | 1.03 [0.77, 1.36] |
|  | Bexley | 2.08 [1.68, 2.59] | 2.03 [1.64, 2.52] |
|  | Birmingham | 0.96 [0.79, 1.16] | 0.93 [0.77, 1.13] |
|  | Blackburn with Darwen | 1.66 [1.29, 2.14] | 1.57 [1.22, 2.03] |
|  | Blackpool | 1.21 [0.91, 1.62] | 1.15 [0.86, 1.53] |
|  | Bolton | 0.90 [0.70, 1.16] | 0.87 [0.67, 1.12] |
|  | Bournemouth, Christchurch and Poole | 1.54 [1.25, 1.90] | 1.49 [1.21, 1.84] |
|  | Bracknell Forest | 1.42 [1.07, 1.88] | 1.41 [1.06, 1.86] |
|  | Bradford | 1.45 [1.18, 1.77] | 1.36 [1.11, 1.66] |
|  | Brent | 1.14 [0.91, 1.43] | 1.15 [0.91, 1.44] |
|  | Brighton and Hove | 1.48 [1.19, 1.84] | 1.50 [1.20, 1.86] |
|  | Bristol, City of | 1.31 [1.07, 1.61] | 1.29 [1.05, 1.58] |
|  | Bromley | 2.01 [1.64, 2.47] | 1.93 [1.57, 2.37] |
|  | Buckinghamshire | 1.18 [0.95, 1.45] | 1.17 [0.95, 1.45] |
|  | Bury | 1.00 [0.76, 1.32] | 0.97 [0.73, 1.27] |
|  | Calderdale | 1.15 [0.89, 1.49] | 1.10 [0.85, 1.43] |
|  | Cambridgeshire | 1.38 [1.14, 1.69] | 1.37 [1.13, 1.67] |
|  | Camden | 0.85 [0.65, 1.10] | 0.87 [0.67, 1.14] |
|  | Central Bedfordshire | 1.27 [1.01, 1.61] | 1.24 [0.98, 1.57] |
|  | Cheshire East | 1.32 [1.06, 1.64] | 1.28 [1.03, 1.59] |
|  | Cheshire West and Chester | 1.24 [0.99, 1.55] | 1.19 [0.95, 1.49] |
|  | Cornwall and Isles of Scilly | 1.57 [1.28, 1.92] | 1.50 [1.22, 1.83] |
|  | County Durham | 1.30 [1.06, 1.60] | 1.25 [1.01, 1.53] |
|  | Coventry | 1.08 [0.85, 1.36] | 1.05 [0.83, 1.32] |
|  | Croydon | 1.28 [1.03, 1.59] | 1.24 [1.00, 1.54] |
|  | Cumbria | 1.08 [0.87, 1.34] | 1.05 [0.85, 1.31] |
|  | Darlington | 1.58 [1.18, 2.10] | 1.52 [1.14, 2.02] |
|  | Derby | 1.28 [1.01, 1.62] | 1.22 [0.96, 1.54] |
|  | Derbyshire | 1.41 [1.17, 1.72] | 1.36 [1.12, 1.65] |
|  | Devon | 2.05 [1.70, 2.48] | 2.00 [1.66, 2.41] |
|  | Doncaster | 1.05 [0.82, 1.33] | 1.02 [0.80, 1.29] |
|  | Dorset | 1.98 [1.61, 2.44] | 1.91 [1.55, 2.35] |
|  | Dudley | 1.16 [0.92, 1.46] | 1.11 [0.88, 1.39] |
|  | Ealing | 0.98 [0.78, 1.24] | 0.98 [0.78, 1.23] |
|  | East Riding of Yorkshire | 1.57 [1.26, 1.95] | 1.51 [1.22, 1.88] |
|  | East Sussex | 1.54 [1.25, 1.88] | 1.50 [1.23, 1.85] |
|  | Enfield | 0.88 [0.69, 1.13] | 0.86 [0.68, 1.10] |
|  | Essex | 1.55 [1.29, 1.86] | 1.52 [1.27, 1.82] |
|  | Gateshead | 0.92 [0.70, 1.21] | 0.90 [0.68, 1.18] |
|  | Gloucestershire | 1.34 [1.10, 1.64] | 1.31 [1.07, 1.60] |
|  | Greenwich | 1.64 [1.32, 2.04] | 1.61 [1.29, 2.00] |
|  | Halton | 1.34 [1.01, 1.78] | 1.26 [0.95, 1.68] |
|  | Hammersmith and Fulham | 1.08 [0.84, 1.39] | 1.10 [0.86, 1.42] |
|  | Hampshire | 1.94 [1.62, 2.32] | 1.90 [1.59, 2.28] |
|  | Haringey | 0.91 [0.71, 1.17] | 0.92 [0.71, 1.18] |
|  | Harrow | 1.05 [0.82, 1.35] | 1.06 [0.82, 1.36] |
|  | Hartlepool | 0.84 [0.58, 1.23] | 0.80 [0.54, 1.17] |
|  | Havering | 1.83 [1.47, 2.28] | 1.81 [1.45, 2.26] |
|  | Herefordshire, County of | 2.30 [1.82, 2.89] | 2.26 [1.79, 2.85] |
|  | Hertfordshire | 1.23 [1.02, 1.48] | 1.20 [1.00, 1.45] |
|  | Hillingdon | 1.12 [0.89, 1.42] | 1.10 [0.87, 1.40] |
|  | Hounslow | 0.90 [0.70, 1.15] | 0.90 [0.70, 1.15] |
|  | Isle of Wight | 1.60 [1.21, 2.11] | 1.60 [1.22, 2.12] |
|  | Islington | 0.77 [0.59, 1.01] | 0.79 [0.61, 1.03] |
|  | Kensington and Chelsea | 1.00 [0.75, 1.35] | 1.03 [0.76, 1.38] |
|  | Kent | 1.72 [1.44, 2.06] | 1.70 [1.42, 2.03] |
|  | Kingston upon Hull, City of | 1.96 [1.58, 2.43] | 1.85 [1.49, 2.29] |
|  | Kingston upon Thames | 1.29 [0.99, 1.67] | 1.29 [1.00, 1.67] |
|  | Kirklees | 1.48 [1.20, 1.82] | 1.43 [1.16, 1.76] |
|  | Knowsley | 1.38 [1.05, 1.80] | 1.30 [0.99, 1.69] |
|  | Lambeth | 1.03 [0.82, 1.30] | 1.05 [0.84, 1.32] |
|  | Lancashire | 1.50 [1.25, 1.80] | 1.43 [1.19, 1.72] |
|  | Leeds | 1.16 [0.95, 1.41] | 1.13 [0.93, 1.37] |
|  | Leicester | 1.17 [0.94, 1.46] | 1.12 [0.90, 1.40] |
|  | Leicestershire | 1.28 [1.05, 1.56] | 1.25 [1.03, 1.53] |
|  | Lewisham | 1.39 [1.12, 1.74] | 1.35 [1.08, 1.68] |
|  | Lincolnshire | 2.11 [1.75, 2.55] | 2.04 [1.70, 2.47] |
|  | Liverpool | 1.22 [0.99, 1.50] | 1.17 [0.95, 1.45] |
|  | Luton | 1.41 [1.11, 1.79] | 1.37 [1.08, 1.74] |
|  | Manchester | 1.08 [0.88, 1.33] | 1.05 [0.85, 1.29] |
|  | Medway | 2.48 [2.02, 3.05] | 2.44 [1.99, 3.00] |
|  | Merton | 1.03 [0.80, 1.33] | 1.03 [0.80, 1.33] |
|  | Middlesbrough | 1.45 [1.11, 1.90] | 1.36 [1.04, 1.79] |
|  | Milton Keynes | 1.65 [1.32, 2.05] | 1.61 [1.29, 2.00] |
|  | Newcastle upon Tyne | 0.77 [0.59, 1.00] | 0.76 [0.58, 0.98] |
|  | Newham | 1.36 [1.09, 1.69] | 1.32 [1.06, 1.64] |
|  | Norfolk | 1.17 [0.96, 1.43] | 1.14 [0.94, 1.39] |
|  | North East Lincolnshire | 1.29 [0.98, 1.69] | 1.27 [0.97, 1.67] |
|  | North Lincolnshire | 1.11 [0.84, 1.47] | 1.09 [0.82, 1.44] |
|  | North Northamptonshire | 1.68 [1.36, 2.08] | 1.63 [1.31, 2.01] |
|  | North Somerset | 1.55 [1.22, 1.98] | 1.49 [1.17, 1.89] |
|  | North Tyneside | 0.95 [0.73, 1.25] | 0.91 [0.69, 1.19] |
|  | North Yorkshire | 1.15 [0.94, 1.42] | 1.13 [0.92, 1.39] |
|  | Northumberland | 1.02 [0.80, 1.30] | 0.97 [0.76, 1.24] |
|  | Nottingham | 0.77 [0.60, 0.98] | 0.75 [0.58, 0.96] |
|  | Nottinghamshire | 1.45 [1.19, 1.75] | 1.39 [1.15, 1.69] |
|  | Oldham | 1.97 [1.58, 2.46] | 1.88 [1.50, 2.34] |
|  | Oxfordshire | 1.29 [1.06, 1.57] | 1.29 [1.06, 1.57] |
|  | Peterborough | 1.84 [1.46, 2.32] | 1.79 [1.42, 2.26] |
|  | Plymouth | 1.18 [0.93, 1.50] | 1.14 [0.90, 1.45] |
|  | Portsmouth | 1.38 [1.08, 1.75] | 1.34 [1.05, 1.71] |
|  | Reading | 0.84 [0.63, 1.12] | 0.85 [0.63, 1.13] |
|  | Redbridge | 1.13 [0.90, 1.43] | 1.13 [0.89, 1.43] |
|  | Redcar and Cleveland | 1.62 [1.24, 2.12] | 1.55 [1.19, 2.02] |
|  | Richmond upon Thames | 1.01 [0.77, 1.31] | 1.00 [0.77, 1.31] |
|  | Rochdale | 1.79 [1.42, 2.25] | 1.70 [1.35, 2.13] |
|  | Rotherham | 1.83 [1.47, 2.27] | 1.70 [1.37, 2.12] |
|  | Rutland | 1.58 [1.00, 2.51] | 1.56 [0.99, 2.48] |
|  | Salford | 0.95 [0.74, 1.23] | 0.90 [0.70, 1.16] |
|  | Sandwell | 1.06 [0.84, 1.34] | 1.01 [0.80, 1.28] |
|  | Sefton | 1.37 [1.08, 1.73] | 1.32 [1.04, 1.66] |
|  | Sheffield | 1.27 [1.04, 1.56] | 1.23 [1.01, 1.51] |
|  | Shropshire | 1.26 [0.99, 1.59] | 1.25 [0.99, 1.58] |
|  | Slough | 1.05 [0.79, 1.39] | 1.03 [0.77, 1.36] |
|  | Solihull | 0.97 [0.74, 1.27] | 0.94 [0.72, 1.24] |
|  | Somerset | 1.63 [1.34, 2.00] | 1.58 [1.29, 1.93] |
|  | South Gloucestershire | 1.73 [1.39, 2.16] | 1.69 [1.36, 2.11] |
|  | South Tyneside | 2.08 [1.63, 2.65] | 1.99 [1.56, 2.53] |
|  | Southampton | 2.29 [1.86, 2.82] | 2.25 [1.83, 2.77] |
|  | Southend-on-Sea | 1.33 [1.03, 1.73] | 1.30 [1.00, 1.69] |
|  | Southwark | 1.04 [0.82, 1.31] | 1.04 [0.83, 1.31] |
|  | St. Helens | 1.21 [0.92, 1.58] | 1.15 [0.88, 1.50] |
|  | Staffordshire | 1.70 [1.41, 2.05] | 1.65 [1.37, 1.99] |
|  | Stockport | 2.14 [1.73, 2.63] | 2.04 [1.66, 2.51] |
|  | Stockton-on-Tees | 1.28 [0.99, 1.65] | 1.22 [0.94, 1.57] |
|  | Stoke-on-Trent | 1.71 [1.37, 2.14] | 1.62 [1.30, 2.03] |
|  | Suffolk | 1.17 [0.95, 1.43] | 1.14 [0.93, 1.40] |
|  | Sunderland | 1.28 [1.01, 1.61] | 1.22 [0.97, 1.54] |
|  | Surrey | 1.98 [1.65, 2.37] | 1.96 [1.64, 2.35] |
|  | Sutton | 1.20 [0.93, 1.54] | 1.17 [0.91, 1.51] |
|  | Swindon | 2.69 [2.18, 3.31] | 2.63 [2.13, 3.24] |
|  | Tameside | 1.89 [1.51, 2.35] | 1.77 [1.42, 2.21] |
|  | Telford and Wrekin | 1.16 [0.88, 1.53] | 1.13 [0.86, 1.49] |
|  | Thurrock | 1.27 [0.97, 1.66] | 1.24 [0.95, 1.62] |
|  | Torbay | 2.85 [2.25, 3.61] | 2.74 [2.16, 3.47] |
|  | Tower Hamlets | 1.45 [1.16, 1.80] | 1.48 [1.19, 1.84] |
|  | Trafford | 1.98 [1.59, 2.47] | 1.91 [1.53, 2.38] |
|  | Wakefield | 1.85 [1.50, 2.28] | 1.80 [1.46, 2.22] |
|  | Walsall | 2.18 [1.77, 2.69] | 2.10 [1.70, 2.60] |
|  | Waltham Forest | 1.17 [0.93, 1.48] | 1.15 [0.91, 1.46] |
|  | Wandsworth | 1.10 [0.88, 1.36] | 1.13 [0.91, 1.40] |
|  | Warrington | 1.20 [0.93, 1.54] | 1.15 [0.90, 1.49] |
|  | Warwickshire | 1.31 [1.07, 1.60] | 1.28 [1.04, 1.57] |
|  | West Berkshire | 1.27 [0.97, 1.67] | 1.27 [0.97, 1.66] |
|  | West Northamptonshire | 1.14 [0.92, 1.43] | 1.11 [0.89, 1.38] |
|  | West Sussex | 1.84 [1.53, 2.22] | 1.81 [1.50, 2.19] |
|  | Westminster | 0.96 [0.74, 1.25] | 0.99 [0.77, 1.29] |
|  | Wigan | 1.10 [0.87, 1.39] | 1.06 [0.84, 1.33] |
|  | Wiltshire | 1.64 [1.34, 2.01] | 1.61 [1.31, 1.97] |
|  | Windsor and Maidenhead | 0.86 [0.63, 1.18] | 0.86 [0.62, 1.18] |
|  | Wirral | 2.28 [1.86, 2.80] | 2.17 [1.77, 2.66] |
|  | Wokingham | 0.77 [0.56, 1.05] | 0.77 [0.56, 1.06] |
|  | Wolverhampton | 1.66 [1.32, 2.08] | 1.61 [1.28, 2.01] |
|  | Worcestershire | 2.22 [1.83, 2.68] | 2.18 [1.80, 2.64] |
|  | York | 1.22 [0.95, 1.57] | 1.22 [0.95, 1.57] |
| Secondary sensitivity analysis | Barking and Dagenham | 1.11 [0.93, 1.33] | 1.07 [0.89, 1.28] |
|  | Barnet | 0.97 [0.83, 1.14] | 0.96 [0.82, 1.13] |
|  | Barnsley | 1.67 [1.42, 1.95] | 1.57 [1.34, 1.83] |
|  | Bath and North East Somerset | 0.95 [0.78, 1.16] | 0.96 [0.78, 1.16] |
|  | Bedford | 0.94 [0.77, 1.15] | 0.93 [0.76, 1.14] |
|  | Bexley | 1.91 [1.64, 2.23] | 1.87 [1.60, 2.17] |
|  | Birmingham | 1.28 [1.13, 1.46] | 1.24 [1.09, 1.41] |
|  | Blackburn with Darwen | 1.99 [1.69, 2.36] | 1.88 [1.59, 2.23] |
|  | Blackpool | 1.31 [1.08, 1.58] | 1.23 [1.02, 1.49] |
|  | Bolton | 1.24 [1.06, 1.46] | 1.20 [1.02, 1.41] |
|  | Bournemouth, Christchurch and Poole | 1.58 [1.36, 1.82] | 1.52 [1.31, 1.76] |
|  | Bracknell Forest | 1.17 [0.95, 1.44] | 1.17 [0.95, 1.43] |
|  | Bradford | 1.71 [1.49, 1.96] | 1.59 [1.39, 1.83] |
|  | Brent | 1.34 [1.15, 1.57] | 1.35 [1.15, 1.57] |
|  | Brighton and Hove | 1.09 [0.92, 1.28] | 1.10 [0.93, 1.29] |
|  | Bristol, City of | 1.38 [1.19, 1.59] | 1.34 [1.16, 1.55] |
|  | Bromley | 1.87 [1.62, 2.16] | 1.79 [1.55, 2.07] |
|  | Buckinghamshire | 1.06 [0.91, 1.23] | 1.06 [0.92, 1.23] |
|  | Bury | 1.07 [0.89, 1.28] | 1.03 [0.85, 1.24] |
|  | Calderdale | 1.04 [0.86, 1.24] | 1.00 [0.83, 1.20] |
|  | Cambridgeshire | 1.27 [1.11, 1.46] | 1.27 [1.10, 1.46] |
|  | Camden | 1.09 [0.92, 1.30] | 1.13 [0.95, 1.34] |
|  | Central Bedfordshire | 1.29 [1.09, 1.51] | 1.26 [1.07, 1.48] |
|  | Cheshire East | 1.25 [1.08, 1.46] | 1.22 [1.05, 1.42] |
|  | Cheshire West and Chester | 1.29 [1.11, 1.50] | 1.23 [1.06, 1.44] |
|  | Cornwall and Isles of Scilly | 1.62 [1.41, 1.86] | 1.55 [1.35, 1.78] |
|  | County Durham | 1.29 [1.12, 1.49] | 1.23 [1.07, 1.42] |
|  | Coventry | 1.29 [1.11, 1.51] | 1.25 [1.07, 1.46] |
|  | Croydon | 1.16 [0.99, 1.35] | 1.12 [0.96, 1.31] |
|  | Cumbria | 0.96 [0.83, 1.12] | 0.94 [0.81, 1.09] |
|  | Darlington | 1.46 [1.19, 1.78] | 1.41 [1.15, 1.72] |
|  | Derby | 1.47 [1.26, 1.73] | 1.39 [1.19, 1.63] |
|  | Derbyshire | 1.61 [1.41, 1.84] | 1.55 [1.36, 1.77] |
|  | Devon | 1.95 [1.71, 2.22] | 1.90 [1.67, 2.17] |
|  | Doncaster | 1.28 [1.09, 1.50] | 1.24 [1.06, 1.45] |
|  | Dorset | 1.88 [1.63, 2.17] | 1.81 [1.57, 2.09] |
|  | Dudley | 1.12 [0.96, 1.32] | 1.06 [0.91, 1.25] |
|  | Ealing | 0.97 [0.82, 1.13] | 0.96 [0.82, 1.13] |
|  | East Riding of Yorkshire | 1.37 [1.18, 1.60] | 1.32 [1.13, 1.54] |
|  | East Sussex | 1.42 [1.23, 1.64] | 1.40 [1.21, 1.61] |
|  | Enfield | 1.03 [0.87, 1.21] | 1.00 [0.85, 1.18] |
|  | Essex | 1.48 [1.31, 1.68] | 1.46 [1.28, 1.66] |
|  | Gateshead | 1.06 [0.88, 1.27] | 1.03 [0.86, 1.24] |
|  | Gloucestershire | 1.43 [1.24, 1.64] | 1.40 [1.22, 1.61] |
|  | Greenwich | 1.51 [1.30, 1.76] | 1.48 [1.27, 1.72] |
|  | Halton | 1.86 [1.56, 2.22] | 1.74 [1.46, 2.08] |
|  | Hammersmith and Fulham | 0.90 [0.75, 1.09] | 0.92 [0.77, 1.11] |
|  | Hampshire | 1.57 [1.38, 1.78] | 1.55 [1.36, 1.76] |
|  | Haringey | 1.13 [0.95, 1.33] | 1.13 [0.96, 1.34] |
|  | Harrow | 1.17 [0.99, 1.38] | 1.18 [1.00, 1.40] |
|  | Hartlepool | 1.32 [1.06, 1.64] | 1.25 [1.00, 1.55] |
|  | Havering | 1.74 [1.49, 2.03] | 1.73 [1.48, 2.02] |
|  | Herefordshire, County of | 2.06 [1.75, 2.42] | 2.04 [1.74, 2.40] |
|  | Hertfordshire | 1.20 [1.05, 1.36] | 1.18 [1.04, 1.34] |
|  | Hillingdon | 1.26 [1.08, 1.48] | 1.24 [1.06, 1.46] |
|  | Hounslow | 1.02 [0.86, 1.21] | 1.02 [0.86, 1.21] |
|  | Isle of Wight | 1.87 [1.56, 2.24] | 1.90 [1.58, 2.27] |
|  | Islington | 1.06 [0.89, 1.25] | 1.08 [0.91, 1.28] |
|  | Kensington and Chelsea | 0.98 [0.79, 1.20] | 1.00 [0.81, 1.23] |
|  | Kent | 1.50 [1.32, 1.70] | 1.48 [1.31, 1.69] |
|  | Kingston upon Hull, City of | 1.66 [1.43, 1.94] | 1.56 [1.34, 1.82] |
|  | Kingston upon Thames | 1.20 [1.00, 1.44] | 1.20 [1.00, 1.44] |
|  | Kirklees | 1.38 [1.20, 1.60] | 1.34 [1.16, 1.55] |
|  | Knowsley | 2.10 [1.78, 2.48] | 1.97 [1.67, 2.32] |
|  | Lambeth | 0.95 [0.81, 1.12] | 0.97 [0.83, 1.14] |
|  | Lancashire | 1.65 [1.45, 1.88] | 1.57 [1.38, 1.79] |
|  | Leeds | 1.12 [0.98, 1.29] | 1.09 [0.95, 1.26] |
|  | Leicester | 1.27 [1.09, 1.48] | 1.21 [1.04, 1.42] |
|  | Leicestershire | 1.44 [1.26, 1.65] | 1.41 [1.23, 1.62] |
|  | Lewisham | 1.33 [1.14, 1.55] | 1.28 [1.10, 1.50] |
|  | Lincolnshire | 1.88 [1.65, 2.14] | 1.82 [1.60, 2.08] |
|  | Liverpool | 1.90 [1.66, 2.18] | 1.83 [1.59, 2.09] |
|  | Luton | 1.52 [1.29, 1.80] | 1.48 [1.26, 1.75] |
|  | Manchester | 1.13 [0.98, 1.31] | 1.10 [0.95, 1.27] |
|  | Medway | 2.70 [2.35, 3.11] | 2.67 [2.32, 3.08] |
|  | Merton | 1.04 [0.87, 1.24] | 1.04 [0.87, 1.24] |
|  | Middlesbrough | 1.20 [0.98, 1.46] | 1.12 [0.92, 1.37] |
|  | Milton Keynes | 1.73 [1.48, 2.01] | 1.68 [1.45, 1.96] |
|  | Newcastle upon Tyne | 0.91 [0.76, 1.08] | 0.88 [0.74, 1.05] |
|  | Newham | 1.34 [1.15, 1.57] | 1.29 [1.11, 1.51] |
|  | Norfolk | 1.23 [1.07, 1.41] | 1.20 [1.05, 1.37] |
|  | North East Lincolnshire | 1.20 [1.00, 1.46] | 1.19 [0.99, 1.44] |
|  | North Lincolnshire | 0.95 [0.78, 1.17] | 0.94 [0.77, 1.15] |
|  | North Northamptonshire | 1.54 [1.33, 1.79] | 1.49 [1.28, 1.73] |
|  | North Somerset | 1.98 [1.69, 2.31] | 1.89 [1.62, 2.21] |
|  | North Tyneside | 1.19 [1.00, 1.41] | 1.12 [0.94, 1.34] |
|  | North Yorkshire | 1.11 [0.96, 1.29] | 1.10 [0.95, 1.27] |
|  | Northumberland | 1.02 [0.87, 1.21] | 0.97 [0.83, 1.15] |
|  | Nottingham | 1.23 [1.05, 1.43] | 1.20 [1.02, 1.40] |
|  | Nottinghamshire | 1.56 [1.37, 1.78] | 1.51 [1.32, 1.72] |
|  | Oldham | 1.85 [1.58, 2.16] | 1.75 [1.50, 2.05] |
|  | Oxfordshire | 1.17 [1.02, 1.34] | 1.17 [1.02, 1.34] |
|  | Peterborough | 1.62 [1.37, 1.91] | 1.58 [1.34, 1.87] |
|  | Plymouth | 1.64 [1.41, 1.91] | 1.59 [1.36, 1.85] |
|  | Portsmouth | 1.19 [1.00, 1.42] | 1.16 [0.97, 1.38] |
|  | Reading | 0.97 [0.80, 1.18] | 0.99 [0.81, 1.20] |
|  | Redbridge | 1.16 [0.98, 1.36] | 1.15 [0.98, 1.36] |
|  | Redcar and Cleveland | 1.36 [1.12, 1.64] | 1.29 [1.07, 1.57] |
|  | Richmond upon Thames | 0.99 [0.82, 1.19] | 0.99 [0.82, 1.19] |
|  | Rochdale | 1.53 [1.30, 1.80] | 1.45 [1.23, 1.70] |
|  | Rotherham | 1.72 [1.48, 2.01] | 1.60 [1.37, 1.86] |
|  | Rutland | 1.31 [0.94, 1.83] | 1.30 [0.94, 1.82] |
|  | Salford | 1.12 [0.94, 1.33] | 1.05 [0.88, 1.25] |
|  | Sandwell | 1.13 [0.96, 1.33] | 1.08 [0.92, 1.27] |
|  | Sefton | 2.01 [1.74, 2.34] | 1.93 [1.67, 2.24] |
|  | Sheffield | 1.09 [0.95, 1.26] | 1.06 [0.91, 1.22] |
|  | Shropshire | 1.48 [1.26, 1.72] | 1.47 [1.26, 1.71] |
|  | Slough | 1.11 [0.91, 1.35] | 1.09 [0.89, 1.32] |
|  | Solihull | 0.97 [0.81, 1.17] | 0.95 [0.79, 1.14] |
|  | Somerset | 1.45 [1.26, 1.66] | 1.40 [1.22, 1.61] |
|  | South Gloucestershire | 1.82 [1.56, 2.11] | 1.78 [1.53, 2.07] |
|  | South Tyneside | 2.07 [1.75, 2.44] | 1.97 [1.67, 2.33] |
|  | Southampton | 1.95 [1.68, 2.27] | 1.92 [1.65, 2.23] |
|  | Southend-on-Sea | 1.28 [1.07, 1.53] | 1.25 [1.04, 1.50] |
|  | Southwark | 0.95 [0.81, 1.12] | 0.95 [0.81, 1.12] |
|  | St. Helens | 1.67 [1.41, 1.97] | 1.58 [1.34, 1.87] |
|  | Staffordshire | 1.68 [1.48, 1.92] | 1.63 [1.43, 1.86] |
|  | Stockport | 1.93 [1.66, 2.23] | 1.84 [1.59, 2.13] |
|  | Stockton-on-Tees | 1.49 [1.26, 1.77] | 1.42 [1.20, 1.68] |
|  | Stoke-on-Trent | 1.85 [1.59, 2.16] | 1.75 [1.51, 2.04] |
|  | Suffolk | 1.17 [1.02, 1.34] | 1.15 [1.00, 1.32] |
|  | Sunderland | 1.36 [1.16, 1.59] | 1.30 [1.11, 1.52] |
|  | Surrey | 1.57 [1.38, 1.79] | 1.57 [1.38, 1.78] |
|  | Sutton | 1.15 [0.96, 1.37] | 1.13 [0.94, 1.34] |
|  | Swindon | 2.34 [2.02, 2.71] | 2.29 [1.98, 2.66] |
|  | Tameside | 1.68 [1.43, 1.96] | 1.57 [1.34, 1.84] |
|  | Telford and Wrekin | 1.54 [1.30, 1.83] | 1.51 [1.27, 1.79] |
|  | Thurrock | 1.38 [1.15, 1.65] | 1.35 [1.12, 1.61] |
|  | Torbay | 2.54 [2.15, 3.00] | 2.45 [2.07, 2.89] |
|  | Tower Hamlets | 1.31 [1.12, 1.53] | 1.34 [1.14, 1.57] |
|  | Trafford | 1.51 [1.28, 1.77] | 1.45 [1.23, 1.70] |
|  | Wakefield | 1.69 [1.46, 1.95] | 1.64 [1.42, 1.90] |
|  | Walsall | 1.92 [1.66, 2.23] | 1.86 [1.60, 2.16] |
|  | Waltham Forest | 1.21 [1.03, 1.43] | 1.19 [1.01, 1.40] |
|  | Wandsworth | 1.07 [0.92, 1.25] | 1.11 [0.95, 1.29] |
|  | Warrington | 1.53 [1.30, 1.80] | 1.47 [1.25, 1.73] |
|  | Warwickshire | 1.41 [1.23, 1.63] | 1.39 [1.21, 1.60] |
|  | West Berkshire | 1.24 [1.03, 1.49] | 1.24 [1.03, 1.49] |
|  | West Northamptonshire | 1.20 [1.03, 1.39] | 1.16 [1.00, 1.35] |
|  | West Sussex | 1.55 [1.36, 1.77] | 1.53 [1.34, 1.75] |
|  | Westminster | 0.94 [0.79, 1.14] | 0.97 [0.81, 1.17] |
|  | Wigan | 1.20 [1.03, 1.40] | 1.15 [0.98, 1.35] |
|  | Wiltshire | 1.59 [1.38, 1.84] | 1.56 [1.36, 1.80] |
|  | Windsor and Maidenhead | 0.92 [0.75, 1.14] | 0.92 [0.75, 1.14] |
|  | Wirral | 1.92 [1.66, 2.22] | 1.83 [1.58, 2.11] |
|  | Wokingham | 0.96 [0.79, 1.17] | 0.97 [0.80, 1.19] |
|  | Wolverhampton | 1.92 [1.65, 2.24] | 1.86 [1.60, 2.17] |
|  | Worcestershire | 1.77 [1.54, 2.02] | 1.74 [1.52, 2.00] |
|  | York | 1.05 [0.88, 1.26] | 1.05 [0.87, 1.26] |

Notes: Reference category: City of London and Hackney

**Supplementary Table 17: Monthly counts of first endometriosis diagnoses since Census Day (27 March 2011)**

| **Month** | **Main analysis,**  **Count** | **Secondary analysis,**  **Count** |
| --- | --- | --- |
| April 2011 | 1,030 | 2,375 |
| May 2011 | 1,235 | 2,840 |
| June 2011 | 1,255 | 2,910 |
| July 2011 | 1,200 | 2,625 |
| August 2011 | 1,150 | 2,560 |
| September 2011 | 1,210 | 2,675 |
| October 2011 | 1,245 | 2,585 |
| November 2011 | 1,295 | 2,795 |
| December 2011 | 1,095 | 2,265 |
| January 2012 | 1,220 | 2,695 |
| February 2012 | 1,185 | 2,500 |
| March 2012 | 1,240 | 2,535 |
| April 2012 | 1,070 | 2,155 |
| May 2012 | 1,185 | 2,510 |
| June 2012 | 1,090 | 2,260 |
| July 2012 | 1,155 | 2,375 |
| August 2012 | 1,085 | 2,250 |
| September 2012 | 1,120 | 2,185 |
| October 2012 | 1,200 | 2,530 |
| November 2012 | 1,195 | 2,465 |
| December 2012 | 915 | 1,835 |
| January 2013 | 1,235 | 2,520 |
| February 2013 | 1,125 | 2,180 |
| March 2013 | 1,070 | 2,170 |
| April 2013 | 1,175 | 2,300 |
| May 2013 | 1,140 | 2,320 |
| June 2013 | 1,135 | 2,255 |
| July 2013 | 1,120 | 2,390 |
| August 2013 | 985 | 2,070 |
| September 2013 | 1,185 | 2,385 |
| October 2013 | 1,240 | 2,600 |
| November 2013 | 1,145 | 2,300 |
| December 2013 | 1,020 | 1,995 |
| January 2014 | 1,140 | 2,425 |
| February 2014 | 1,030 | 2,105 |
| March 2014 | 1,135 | 2,290 |
| April 2014 | 1,085 | 2,165 |
| May 2014 | 1,045 | 2,120 |
| June 2014 | 1,185 | 2,420 |
| July 2014 | 1,190 | 2,460 |
| August 2014 | 1,000 | 1,990 |
| September 2014 | 1,240 | 2,460 |
| October 2014 | 1,280 | 2,475 |
| November 2014 | 1,170 | 2,340 |
| December 2014 | 1,065 | 2,105 |
| January 2015 | 1,115 | 2,260 |
| February 2015 | 1,190 | 2,245 |
| March 2015 | 1,290 | 2,495 |
| April 2015 | 1,090 | 2,135 |
| May 2015 | 1,050 | 2,105 |
| June 2015 | 1,260 | 2,395 |
| July 2015 | 1,280 | 2,450 |
| August 2015 | 1,045 | 2,115 |
| September 2015 | 1,215 | 2,330 |
| October 2015 | 1,165 | 2,310 |
| November 2015 | 1,225 | 2,415 |
| December 2015 | 1,000 | 2,020 |
| January 2016 | 1,100 | 2,210 |
| February 2016 | 1,225 | 2,380 |
| March 2016 | 1,165 | 2,300 |
| April 2016 | 1,070 | 2,225 |
| May 2016 | 1,150 | 2,260 |
| June 2016 | 1,230 | 2,495 |
| July 2016 | 1,130 | 2,335 |
| August 2016 | 1,130 | 2,250 |
| September 2016 | 1,215 | 2,465 |
| October 2016 | 1,185 | 2,365 |
| November 2016 | 1,285 | 2,525 |
| December 2016 | 980 | 2,010 |
| January 2017 | 1,195 | 2,390 |
| February 2017 | 1,140 | 2,285 |
| March 2017 | 1,335 | 2,660 |
| April 2017 | 1,080 | 2,190 |
| May 2017 | 1,170 | 2,415 |
| June 2017 | 1,300 | 2,655 |
| July 2017 | 1,155 | 2,380 |
| August 2017 | 1,190 | 2,430 |
| September 2017 | 1,200 | 2,460 |
| October 2017 | 1,355 | 2,695 |
| November 2017 | 1,355 | 2,705 |
| December 2017 | 960 | 2,015 |
| January 2018 | 1,180 | 2,415 |
| February 2018 | 1,160 | 2,290 |
| March 2018 | 1,165 | 2,450 |
| April 2018 | 1,155 | 2,430 |
| May 2018 | 1,200 | 2,535 |
| June 2018 | 1,305 | 2,685 |
| July 2018 | 1,270 | 2,715 |
| August 2018 | 1,140 | 2,540 |
| September 2018 | 1,205 | 2,565 |
| October 2018 | 1,325 | 2,755 |
| November 2018 | 1,260 | 2,785 |
| December 2018 | 1,070 | 2,290 |
| January 2019 | 1,315 | 2,735 |
| February 2019 | 1,125 | 2,480 |
| March 2019 | 1,360 | 2,815 |
| April 2019 | 1,275 | 2,680 |
| May 2019 | 1,155 | 2,590 |
| June 2019 | 1,195 | 2,620 |
| July 2019 | 1,390 | 2,995 |
| August 2019 | 1,145 | 2,605 |
| September 2019 | 1,210 | 2,660 |
| October 2019 | 1,340 | 2,925 |
| November 2019 | 1,270 | 2,810 |
| December 2019 | 1,095 | 2,440 |
| January 2020 | 1,350 | 2,860 |
| February 2020 | 1,260 | 2,710 |
| March 2020 | 800 | 1,910 |
| April 2020 | 85 | 560 |
| May 2020 | 200 | 935 |
| June 2020 | 460 | 1,380 |
| July 2020 | 665 | 1,825 |
| August 2020 | 780 | 1,985 |
| September 2020 | 1,065 | 2,470 |
| October 2020 | 1,200 | 2,645 |
| November 2020 | 1,115 | 2,495 |
| December 2020 | 930 | 2,165 |
| January 2021 | 435 | 1,475 |
| February 2021 | 605 | 1,735 |
| March 2021 | 910 | 2,455 |
| April 2021 | 990 | 2,570 |
| May 2021 | 1,170 | 2,875 |
| June 2021 | 1,275 | 3,025 |
| July 2021 | 1,155 | 2,825 |
| August 2021 | 1,010 | 2,495 |
| September 2021 | 1,190 | 2,740 |
| October 2021 | 1,105 | 2,700 |
| November 2021 | 1,240 | 2,930 |
| December 2021 | 965 | 2,400 |
| January 2022 | 960 | 2,485 |
| February 2022 | 1,070 | 2,625 |
| March 2022 | 1,340 | 3,020 |
| April 2022 | 1,105 | 2,695 |
| May 2022 | 1,375 | 3,185 |
| June 2022 | 1,230 | 2,910 |
| July 2022 | 1,080 | 2,705 |
| August 2022 | 1,205 | 2,870 |
| September 2022 | 1,250 | 3,025 |
| October 2022 | 1,230 | 2,980 |
| November 2022 | 1,390 | 3,300 |
| December 2022 | 1,010 | 2,515 |
| January 2023 | 1,205 | 3,045 |
| February 2023 | 1,195 | 2,890 |
| March 2023 | 1,410 | 3,295 |
| April 2023 | 970 | 2,680 |
| May 2023 | 1,355 | 3,285 |
| June 2023 | 1,355 | 3,415 |
